# Temporal patterning of trigeminal nerve stimulation gates hippocampal plasticity across species

**DOI:** 10.64898/2026.08.25.26361320

**Authors:** Liyi Chen, Qiang Sun, Xinxia Guo, Hemmings Wu, Boateng Asamoah, Wentai Ye, Nina Seminck, Haorun Huang, Myles Mc Laughlin

## Abstract

Non-invasive neuromodulation can influence memory, but whether peripheral stimulation can engage hippocampal plasticity through a defined mechanism and translate across species remains unclear. Here we provide, to our knowledge, the first cross-species evidence linking trigeminal nerve stimulation to sustained hippocampal plasticity, direct human hippocampal engagement and associative-memory benefit. In rats, intermittent 200 Hz TNS produced persistent CA1 fEPSP potentiation and prolonged neuronal firing despite substantially lower cumulative charge than continuous 100-Hz stimulation. LC inhibition strongly suppressed these responses. In patients undergoing stereo-EEG monitoring, i200-TNS evoked prominent hippocampal and thalamic responses and increased hippocampal theta–gamma coupling. In a randomized active–sham crossover study, i200-TNS was associated with improved delayed occupation recall and accompanying EEG changes. These results link patterned trigeminal stimulation to hippocampal physiology across species and support its potential for engaging human memory-related networks.

## 1. Introduction

Memory consolidation transforms labile traces into stable, retrievable representations through coordinated synaptic and systems-level plasticity^1,2^. The hippocampus is central to this process: at the synaptic scale, long-term potentiation (LTP) provides a durable increase in synaptic efficacy after high frequency afferent activity^3–5^. At the network scale, brief high frequency “ripple” events are thought to orchestrate replay and redistribution of memory representations across hippocampal neocortical circuits^6,7^. Yet despite decades of mechanistic work, there remain few non-invasive interventions that can reliably engage hippocampal plasticity and modulate ripple dynamics in humans^8^. Existing electrical and magnetic approaches show promise but often yield inconsistent behavioral effects and lack direct physiological validation of LTP-like mechanisms in hippocampus^9–11^.

Peripheral cranial nerve stimulation offers a complementary approach to modulate deep memory circuits via evolutionarily conserved brainstem relays^12^. Trigeminal nerve stimulation (TNS) is especially attractive: trigeminal afferents project to brainstem nuclei that include the locus coeruleus (LC) and dorsal raphe, which then innervate the hippocampus and cortex^13^. These brainstem neurotransmitter pathways can shape hippocampal plasticity and memory^14^. In particular, LC-derived noradrenergic and dopaminergic signalling regulates hippocampal synaptic plasticity^15,16^, while LC-dependent changes in arousal and network state are closely linked to the occurrence of hippocampal sharp-wave ripples^17,18^. Prior work^19–22^ has shown that TNS can influence hippocampal activity and broader cortical dynamics, but whether temporally structured TNS can drive LTP-like synaptic strengthening and targeted modulation of hippocampal ripples remains unknown. Moreover, clinical TNS devices typically deploy continuous pulse trains (e.g., 100 Hz), providing limited control over burst timing, duty cycle, and frequency patterns that are critical for plasticity ^23^.

Here we tested the hypothesis that the temporal pattern of peripheral stimulation, rather than stimulation dose alone, influences its ability to engage hippocampal plasticity in rats. We introduce an intermittent 200 Hz TNS protocol (i200-TNS) modeled after classical LTP inducing bursts in hippocampal pathways, with brief high frequency trains separated by long inter-train intervals^4,24^. The protocol was designed around known features of synaptic induction and neuromodulator dynamics. High-frequency bursts can promote NMDA-dependent plasticity^25,26^, whereas the intervening pauses may favour phasic LC output and transmitter replenishment^27^, potentially optimizing conditions for LTP induction and consolidation^28–30^. We contrast this with a conventional continuous 100 Hz protocol (c100-TNS) that delivers substantially higher cumulative absolute charge but lacks temporal structure thought to gate plasticity.

To establish cross-species validity and mechanistic coherence, we first compared i200-TNS with c100-TNS in rats and measured CA1 fEPSPs, neuronal spiking and ripple dynamics. We then tested i200-TNS in patients undergoing stereo-EEG monitoring to assess hippocampal and thalamic responses and hippocampal network dynamics. Finally, a randomized, sham-controlled study in healthy adults examined associative memory together with scalp EEG.

Our results advance a principle of temporal transduction: properly timed peripheral bursts drive central hippocampal plasticity and optimize ripple dynamics in ways that generalize across species and measurement scales. By linking stimulation timing to canonical synaptic rules and hippocampal biomarkers, i200-TNS provides a mechanistically grounded, noninvasive approach for targeting memory consolidation. The findings further suggest that stimulation timing is an important parameter when targeting hippocampal circuits through peripheral afferents. This provides a basis for testing burst-structured peripheral neuromodulation in future studies.

## 2. Results

### 2.1 Intermittent and continuous TNS produce distinct hippocampal response profiles

Given that the hippocampus is particularly well known for its neuroplasticity, we examined whether TNS influences hippocampal synaptic plasticity. Long-term potentiation (LTP) is a canonical form of synaptic plasticity characterized by a long-lasting increase in synaptic strength following high-frequency stimulation. LTP is typically defined as a sustained increase of more than 20% in the slope of fEPSP relative to baseline, lasting at least 30–60 minutes. In these experiments, a single biphasic pulse was delivered to the Schaffer collateral pathway and the resulting fEPSP was measured in CA1. Classical LTP was induced by 5-minute trains of intermittent high frequency burst stimulation (100-400Hz/s, burst duty cycle: 3.33%) on Schaffer collateral pathway. Fig.1a shows the experimental schematic of fEPSP measurement. fEPSP was measured every 30 seconds before and after TNS, which refers to a 30-minute baseline period and a 90-minute post-stimulation period. Fig.1b (upper panel) shows one rat example of classical LTP induction by applying intermittent 200 Hz intracranial stimulation directly to the Schaffer collaterals for 5 minutes. This induced a robust and sustained increase in fEPSP slope exceeding 20% relative to baseline, consistent with established definitions of LTP.

In this experiment, we compared the effects of i200-TNS and c100-TNS on synaptic plasticity. We measured fEPSPs from the CA1 region in 8 anesthetized rats in each group. Fig.1b (lower panel) shows one example rat with the same intermittent 200 Hz stimulation pattern but applied to the trigeminal nerve (i200-TNS). This non-invasive stimulation also induced a significant increase in fEPSP slope, exceeding 20% of baseline and persisting for the full 90-minute recording period, albeit with a more gradual onset and smaller increment compared to intracranial LTP induction.

Time-resolved group data of i200-TNS are shown on Fig. 1c, faded dots represent normalized fEPSP slope averaged across all rats (n = 8), the error bar shows mean value and 95% confidence interval (CI) of average fEPSP slope in each 10-minute time epoch at rat level. Linear mixed effects (LME) model showed that fEPSP slope was affected by time epoch (F(11, 80) = 10.30, p = 1.93× 10^-11^), LME model and statistical details see supplementary table1. After 5-minutes of i200-TNS, fEPSP increased to 120% and persisted for 90mins (all p < 0.001, post-hoc linear contrast, Bonferroni corrected). Fig.1d shows group comparison of fEPSP slopes 30 minutes before and 30minutes after i200-TNS. Each faded dot refers to the original fEPSP normalized slope at each timepoint over the 30-minute baseline and 30-minute post-stimulation intervals. LME model showed that fEPSP slope was affected by time epoch (baseline 30mins vs. post 30mins, F (1, 951) = 97.55, p = 5.73× 10^-22^). LME model and statistical details see supplementary table2. fEPSP slope was facilitated after 5 mins i200-TNS when compared to baseline 30mins (p = 5.73× 10^-22^, Cohen’s d = 0.64, Linear contrast post-hoc test). Fig. 1e showed c100-TNS fEPSP dynamic change from another 8 rats and with same data visualization structure. However, LME model showed that fEPSP slope was not affected by time epoch (F (11, 84)= 1.02, p = 0.44). LME model and statistical details see supplementary table3. Importantly, no time window after c100-TNS exceeded the 120% threshold commonly used to define LTP (Fig.1e). In group analysis for 30min window before and after c100-TNS, LME model showed that fEPSP slope was not affected by time epoch (baseline 30mins vs. post 30mins, F (1, 939) = 0.43, p = 0.51). LME model and statistical details see supplementary table4.

**Figure 1.**
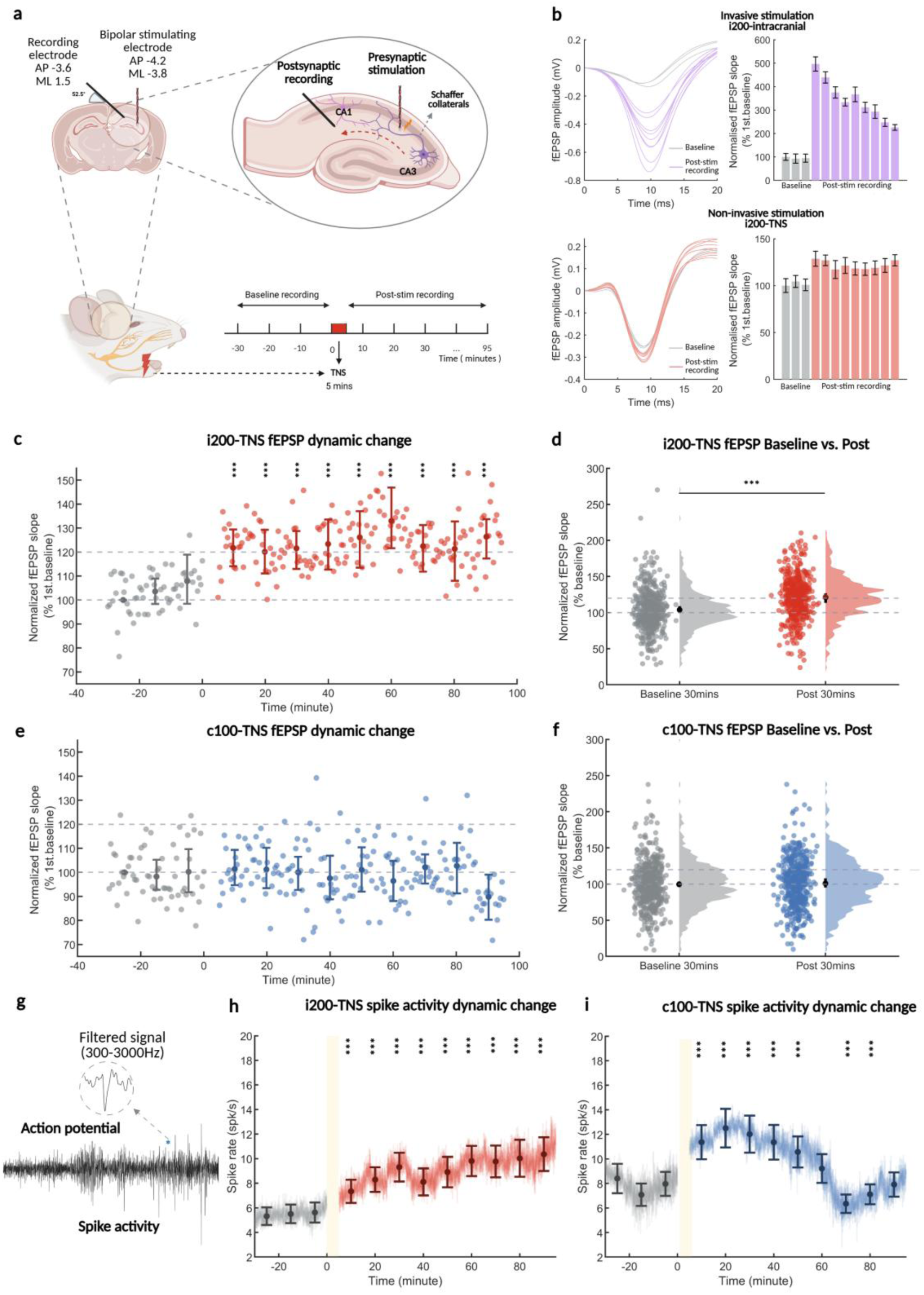
i200-TNS enhances hippocampal synaptic plasticity as measured by field excitatory postsynaptic potentials (fEPSPs). **a.** Schematic of the fEPSP recording setup. A single biphasic stimulation pulse was delivered to the Schaffer collateral pathway, and the inducing postsynaptic response was recorded in the CA1 stratum radiatum. Following a 30-minute baseline recording of fEPSPs (stimulated every 30 seconds), rats received either i200-TNS or c100-TNS for 5 minutes. Post-stimulation fEPSPs were recorded for 90 minutes. **b.** Left panels show averaged fEPSP waveforms before and after stimulation under two conditions. Upper left: intracranial intermittent 200 Hz stimulation applied directly to the Schaffer collaterals (classical LTP induction). Lower left: external intermittent 200 Hz stimulation applied non-invasively to the trigeminal nerve. Right panels show normalized fEPSP slopes grouped into 10-minute bins (each bar represents the average of 20 data points; one fEPSP was recorded every 30 seconds). LTP was operationally defined as a sustained increase of more than 20% in normalized fEPSP slope, maintained for at least 60 minutes after stimulation. **c.** Dynamic changes in fEPSP slope following i200-TNS. Each faded dot represents the average fEPSP slope across 8 rats, measured every 30 seconds. Solid dots indicate group means ± 95% CI, calculated from averaged 20 points at each 10-minute bin across 8 rats. **d.** Group comparison of fEPSP slopes before and after i200-TNS. Each faded dot represents the original fEPSP slope over the 30-minute baseline and 30-minute post-stimulation intervals across 8 rats (60 x 8 data points per interval). Solid dots indicate group means ± 95% CI at rat level. **e.** Dynamic changes in fEPSP slope following c100-TNS, presented as in panel C. **f.** Group comparison of baseline and post-stimulation fEPSP slopes for c100-TNS, presented as in panel D. **g.** Representative signals that show electrophysiological features extracted from hippocampal recordings. Raw action potentials, bandpass-filtered spike signal (300–3000 Hz). **h,i.** Dynamic changes of hippocampal spike rates in the i200-TNS **(h)** and c100-TNS **(i)** groups. Spike rates were averaged across all recorded single units and aligned to stimulation onset (light yellow shading, 0–5 min). Each dot represents mean spike rate across 8 rats at each 10-minute bin. The light trace show the continuous group average across neurons. Data in c, d, e, f, h, i perform post-hoc linear contrast test after linear mixed effect model. p values were Bonferroni-adjusted. Asterisks of c, e, h, i denote time bins with statistically significant difference compared to the first baseline bin from -30mins to -20mins. Significance levels: * p < 0.05, ** p < 0.01, *** p < 0.001.

Next, we investigated whether the synaptic potentiation induced by i200-TNS was accompanied by a corresponding change in hippocampal neuronal output. To address this, we isolated single-unit activity from the same hippocampal recordings and quantified spike-rate dynamics following i200-TNS and c100-TNS. In the i200-TNS group, LME model showed that spike rate was affected by time epoch (F (11, 2257) = 36.00, p = 1.23× 10^-71^). LME model and statistical details see supplementary table5. Fig.1h showed that compared to the first baseline bin (–30 to –20 min), hippocampal spike rate significantly increased after i200-TNS, from post-10 minutes to post-90 mins, which persisted for over 90 mins (all p < 0.001, post-hoc linear contrast, Bonferroni corrected). In c100-TNS group, LME model showed spike rate was affected by time epoch (F (11, 2508) = 38.51, p = 2.88× 10^-77^). LME model and statistical details see supplementary table6. Fig.1i showed a more modest increase in spike rates in c100-TNS group, with significant increase from post-10mins to post-50mins (all p < 0.05), then recovered to the baseline level at post-60mins (p = 1), further decreased at post-70mins (p = 3.07× 10^-4^) and rebound to baseline level at post-80mins (p = 0.09) and post-90mins (p = 1). i200-TNS therefore produced sustained CA1 potentiation despite delivering approximately 15-fold less cumulative charge than c100-TNS (≈0.0008 C versus ≈0.012 C). The increase in single-unit firing was also more persistent after i200-TNS than after c100-TNS.

### 2.2 LC and hippocampal D1/D5 receptor activity mediate i200-TNS–induced fEPSP potentiation

A central prediction of our temporal-patterning hypothesis was that the intermittent burst structure of i200-TNS promotes persistent CA1 potentiation by engaging catecholaminergic pathways that support hippocampal LTP. We therefore investigated whether the observed potentiation required LC activity and local hippocampal D1/D5 receptor signaling. We tested this pathway at two levels by inhibiting the LC with clonidine or blocking D1/D5 receptors locally in CA1. After 30 min of stable baseline recording, rats received local infusion of clonidine into the LC or SCH-23390 into dorsal hippocampal CA1, followed by a 30-min post-infusion blockade period, 5 min of i200-TNS, and 90 min of post-stimulation recording (Fig. 2a,b).

**Figure 2.**
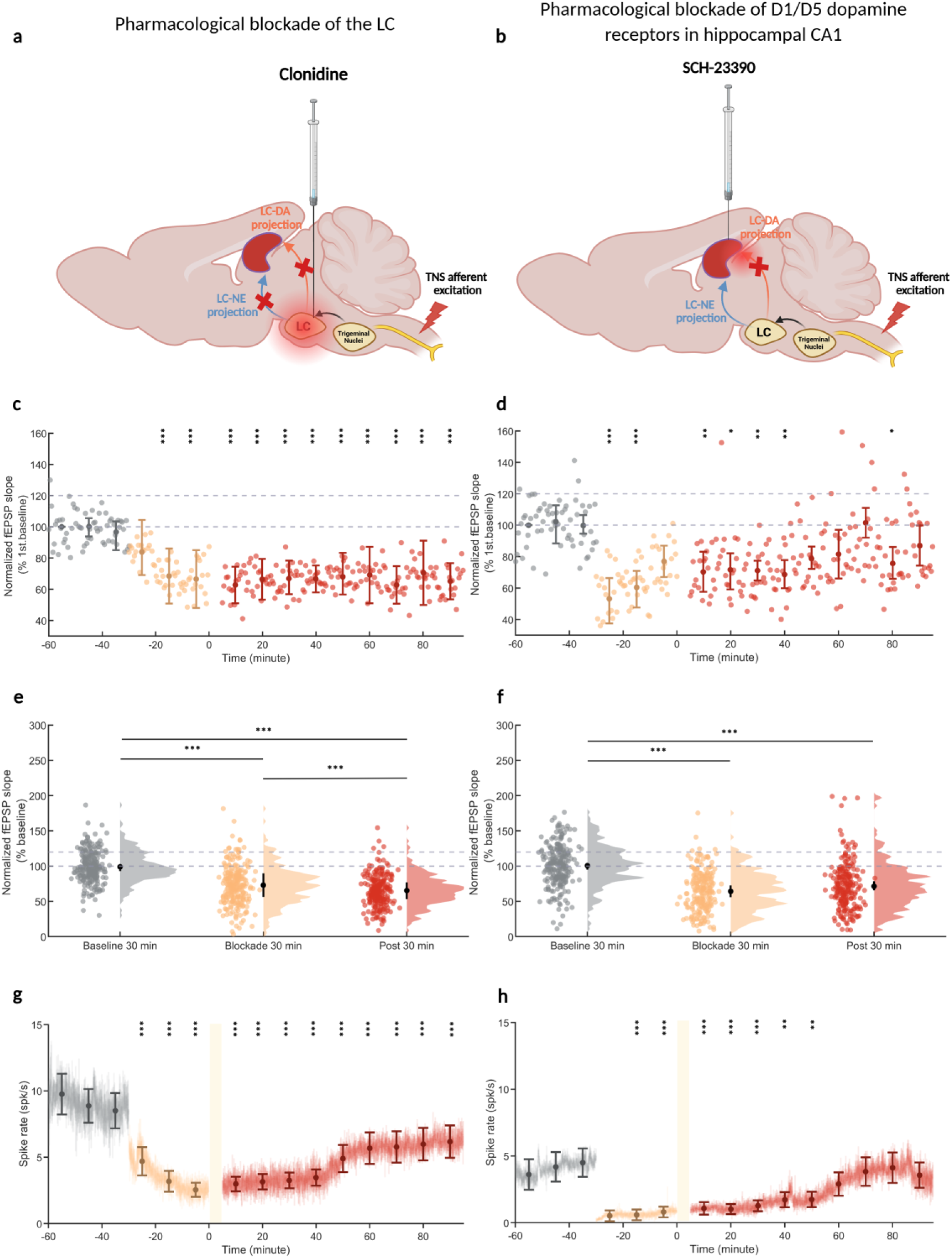
Pharmacological dissection of the catecholaminergic mechanism underlying i200-TNS-induced hippocampal plasticity. **a,b.** Experimental schematics illustrating pharmacological blockade of the proposed catecholaminergic pathway engaged by i200-TNS. **a.** Local infusion of clonidine into the locus coeruleus (LC) to suppress LC activity and reduce LC-derived noradrenergic and dopaminergic influence on the hippocampus. **b.** Local infusion of the D1/D5 receptor antagonist SCH-23390 into dorsal hippocampal CA1 to test whether hippocampal dopamine receptor signaling contributes to i200-TNS-induced plasticity. **c,d.** Time course of normalized fEPSP slope before and after pharmacological manipulation. Gray, baseline recording; orange, 30-min post-infusion blockade period; red, post-stimulation recording following i200-TNS. Intra-LC clonidine markedly suppressed fEPSP slope and prevented the subsequent stimulation-induced potentiation (**c**). Intra-CA1 SCH-23390 also reduced fEPSP slope during the blockade period and altered the subsequent post-stimulation trajectory, but did not fully abolish fEPSP responses (**d**). Solid dots indicate group means ± 95% CI, calculated from averaged 20 points at each 10-min bin across 4 rats. **e,f.** Group comparison of normalized fEPSP slopes during the baseline, blockade and post-stimulation periods for the LC clonidine (**e**) and CA1 SCH-23390 (**f**) experiments. Each faded dot represents the original fEPSP slope values across the 30-min baseline, 30-min post-blockade and 30-min post-stimulation intervals across 4 rats (60 × 4 data points per interval). Solid dots indicate rat-level means ± 95% CI. Clonidine infusion into the LC significantly reduced synaptic responses and blocked the long-lasting potentiation normally induced by i200-TNS. By contrast, SCH-23390 infusion into CA1 significantly reduced fEPSP slope during the blockade period, and the post-stimulation responses were not clearly separable from the blockade epoch, indicating that under the present conditions the effect of local D1/D5 receptor blockade on i200-TNS-induced potentiation could not be resolved unambiguously. **g,h.** Time course of hippocampal spike rate across the same experimental epochs for LC clonidine (**g**) and CA1 SCH-23390 (**h**). Each dot represents mean spike rate across 4 rats at each 10-minute bin; light trace depicts the continuous group average across neurons. Intra-LC clonidine strongly suppressed spontaneous firing and attenuated the stimulation-evoked recovery in spiking, whereas intra-CA1 SCH-23390 produced a transient suppression of firing during the blockade period followed by a gradual post-stimulation increase. Shaded region indicates the 5-min i200-TNS period.

Time-resolved group data for LC blockade is shown in Fig. 2c. Faded dots represent normalized fEPSP slope values across all rats, and solid dots with error bars indicate rat-level mean ± 95% CI for each 10-min time epoch. Following intra-LC clonidine infusion, fEPSP slope was markedly reduced during the blockade period and remained suppressed after i200-TNS. LME model showed that fEPSP slope was affected by time epoch (F(14, 45) = 12.84, p = 2.27× 10^-11^). Compared with the first baseline, fEPSP slope was significantly reduced during the blockade period (10-20mins after infusion) and remained significantly lower throughout the post-stimulation period (all p < 0.001, post hoc linear contrast, Bonferroni corrected, statistical detail see supplementary table 7). Fig. 2e shows group comparison of normalized fEPSP slopes during the 30-min baseline, 30-min blockade, and 30-min post-stimulation intervals. LME model showed that fEPSP slope was affected by time epoch (F(2, 715) = 164.22, p = 2.05× 10^-59^), with significant reductions in both the blockade and post-stimulation periods relative to baseline (all p < 0.001, post hoc linear contrast, Bonferroni corrected, statistical detail see supplementary table 9). These results indicate that LC inhibition abolished the i200-TNS-induced potentiation of hippocampal synaptic responses.

We next examined whether local hippocampal D1/D5 receptor signaling contributes to this effect by infusing SCH-23390 directly into dorsal CA1 before stimulation. Fig. 2d shows the time-resolved change in fEPSP slope following intra-CA1 SCH-23390 infusion. LME model showed that fEPSP slope was affected by time epoch (F(14, 45) = 7.62, p = 7.64× 10^-8^). SCH-23390 reduced fEPSP slope during the 30-min blockade period relative to the first baseline, and post-stimulation responses remained variable over time (statistical detail see supplementary table 8). Group comparison in Fig. 2f likewise showed an effect of time epoch (F(2, 676) = 90.09, p = 2.06× 10^-35^), with a significant reduction during the blockade period compared to baseline (p < 0.001), whereas the post-stimulation period was not significantly different from the blockade period (p = 0.08, Bonferroni corrected, statistical detail see supplementary table 10). Because D1/D5 receptor blockade substantially altered basal synaptic transmission before i200-TNS was delivered, the absence of a clear post-stimulation potentiation cannot be attributed specifically to blockade of the i200-TNS-induced response. These findings are consistent with a contribution of hippocampal D1/D5 signaling to the observed plasticity, but do not establish that D1/D5 receptor activation is required for i200-TNS-induced potentiation.

We next examined hippocampal spike activity under the same pharmacological conditions (Fig. 2g,h). In the LC clonidine group, LME model showed that spike rate was affected by time epoch (F(14, 1485) = 41.20, p = 1.77× 10^-95^), with significant suppression during the blockade and post-stimulation periods relative to the first baseline (all p < 0.001, statistical detail see supplementary table 11). In the SCH-23390 group, LME model showed that spike rate was affected by time epoch (F(14, 1215) = 17.12, p = 4.76× 10^-39^). Spike rate decreased after drug infusion (all p < 0.001, statistical detail see supplementary table 12) and then gradually increased during the post-stimulation period. LC inhibition prevented the sustained synaptic and spiking responses observed after i200-TNS. CA1 D1/D5 receptor blockade also altered both measures, although neuronal firing partially recovered after stimulation. These results support a contribution of local D1/D5 signaling to the hippocampal response. The stronger effect of LC inhibition suggests that other LC-dependent signals, including LC-noradrenergic and LC-dopaminergic signaling, may also contribute.

### 2.3. i200-TNS evokes hippocampal responses and theta–gamma phase–amplitude coupling in human sEEG

To determine whether temporally structured trigeminal nerve stimulation engages human memory-related circuits, we recorded continuous stereo-EEG (sEEG) activity across pre-stimulation, stimulation and post-stimulation periods in nine patients with drug-resistant epilepsy undergoing clinically indicated intracranial monitoring. During a 20-min stimulation period, 1-s bursts of 200-Hz i200-TNS were delivered at 30-s intervals. Electrode contacts were localized using postoperative imaging and grouped according to their anatomical regions of interest (Fig. 3a–c).

**Figure 3.**
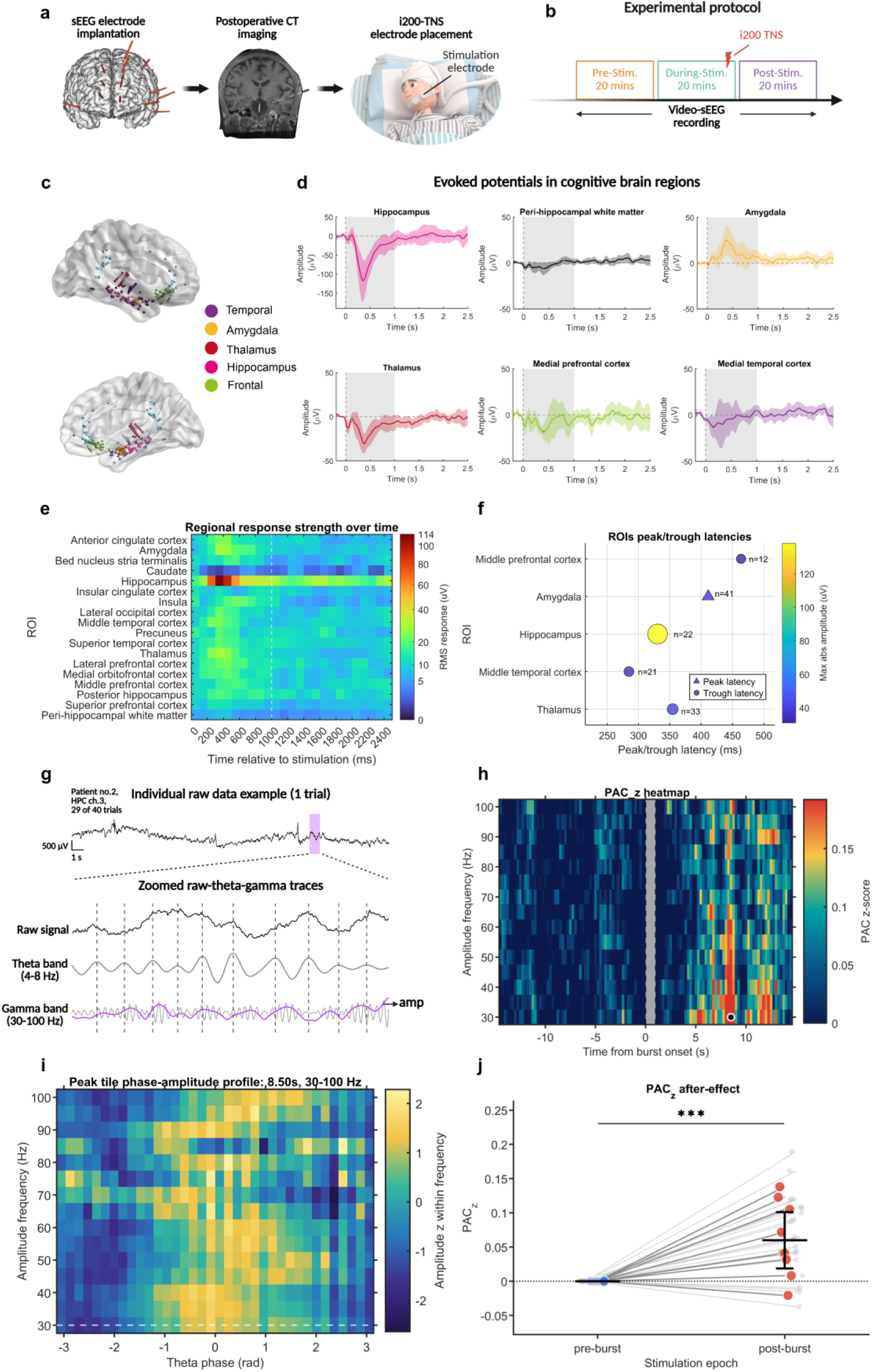
Experimental workflow, TNS-evoked regional sEEG responses and hippocampal theta–gamma phase–amplitude coupling. **a.** Experimental workflow showing sEEG electrode implantation, postoperative CT imaging and placement of the non-invasive i200-TNS electrode. **b.** Experimental protocol comprising 20-min pre-stimulation, stimulation and post-stimulation periods, continuous video-sEEG was recorded throughout. Stimulation consisted of 1-s, 200-Hz TNS bursts. **c.** Three-dimensional reconstruction of implanted sEEG contacts, colour-coded by anatomical region. **d.** Contact-averaged TNS-evoked potentials in selected cortical, limbic and thalamic regions. Lines show the mean response across subjects, and shaded areas indicate approximate 95% confidence intervals. Grey shading marks the 0–1-s stimulation period and the dashed vertical line indicates stimulation onset. Owing to its larger response amplitude, the hippocampal trace is displayed using a separate voltage scale from the other regional traces. The peri-hippocampal white-matter trace is shown as an anatomical control. **e.** Regional response strength over time, quantified as RMS amplitude in successive 100-ms bins from 0 to 2.5 s; the dashed line marks the end of stimulation and colour intensity is displayed on a square-root-transformed scale. **f.** Peak and trough latencies in selected regions. Triangles indicate peak latency, circles indicate trough latency, marker size and colour indicate maximum absolute amplitude, and n denotes the number of sEEG contacts**. g.** Representative single-trial example of theta–gamma coupling from the ipsilateral hippocampus. The upper trace shows the raw burst-centred sEEG signal; the shaded window marks the segment with the highest PAC_z_. Lower traces show the zoomed raw signal, 4–8-Hz theta component, and gamma-band activity with its amplitude envelope. Dashed lines indicate theta peaks. **h.** During-stimulation PAC_z_ heat map for theta phase (4–8 Hz) and gamma amplitude (30–100 Hz). Grey shading marks the 0– 1-s stimulation period, and the white circle indicates the peak PAC_z_ tile. **i.** Phase–amplitude profile from the peak tile in h. Colour indicates amplitude normalized within each gamma-frequency band, plotted as a function of theta phase. **j.** During-stimulation PAC_z_ after-effect comparing pre-burst and post-burst windows. Grey lines indicate paired channel-level observations, colored points indicate subject-level values, and black bars indicate subject-level mean ±95% confidence interval; significance was assessed with a linear mixed-effects model.

i200-TNS evoked spatially differentiated responses across cortical and limbic regions (Fig. 3d– f). The hippocampus exhibited the largest response, characterized by a prominent negative deflection that reached approximately −136.91 μV at around 330.50 ms (subjects-averaged level) after stimulation onset. The thalamus showed a similar negative-going waveform with a smaller amplitude, whereas the amygdala displayed a prominent (polarity-reversed) positive response. Responses in medial prefrontal, medial temporal and other cortical regions were smaller, and peri-hippocampal white-matter contacts did not show a consistent stimulus-locked potential. Root-mean-square (RMS) amplitude, computed in successive 100-ms bins, showed that regional response magnitude was greatest during the 0–1 s stimulation window. Peak- and trough-latency analyses confirmed that the dominant hippocampal and thalamic deflections occurred within the first 500 ms after burst onset.

Since hippocampal theta–gamma coupling organizes faster local activity within slower theta rhythms and has been implicated in associative and episodic memory processing^31–33^, we next examined whether i200-TNS altered theta–gamma phase–amplitude coupling (PAC) around individual stimulation bursts. In a representative hippocampal trial, the raw sEEG signal showed theta-periodic activity accompanied by phase-dependent modulation of gamma amplitude (Fig. 3g). At the group level, PAC_z values increased after burst offset, particularly across the 30–100-Hz amplitude-frequency range, with the largest post-burst tile centered at approximately 8.5 s and 30 Hz (Fig. 3h). The phase–amplitude profile extracted from this tile showed a non-uniform distribution of gamma amplitude across theta phase bins, consistent with theta–gamma coupling (Fig. 3i).

The 0–1-s stimulation interval was excluded from PAC inference because stimulation artefacts and filtering-related distortions can compromise the estimation of phase and amplitude during the burst. Within the stimulation block, PAC_z was therefore compared between pre-burst and post-burst windows. The pre-burst values were close to zero, as expected from trial-wise baseline normalization, whereas post-burst PAC_z was higher across most subject-channel pairs (Fig. 3j). A linear mixed-effects model including period as a fixed effect and random intercepts for subject and subject-channel confirmed a significant post-burst increase in PAC_z (F(1,34.88) = 41.003, p = 2.32 × 10^-7^; 22 subject-channels from 9 patients). These findings indicate that i200-TNS evokes a robust hippocampal response and was followed by a transient increase in hippocampal theta–gamma coupling.

### 2.4 Brain-state-dependent effects of i200-TNS on hippocampal ripple dynamics

To link the rodent physiology with the human sEEG findings, we quantified sharp-wave and ripple dynamics across pre-stimulation, stimulation and post-stimulation periods. In the anesthetized rat experiments, sharp-wave rate, sharp-wave–ripple (SWR) rate and ripple rate were calculated as the number of detected events per minute from 100–250 Hz filtered hippocampal recordings. Ripple duration was defined as the interval between event onset and offset. In the human sEEG recordings, ripple events were identified from hippocampal contacts after 80–120 Hz band-pass filtering, envelope-based event detection and exclusion of events outside the predefined duration range. Ripple rate and mean ripple duration were then compared across three 20-min recording periods in nine patients.

In anesthetized rats, i200-TNS produced a transient increase in sharp-wave, SWR and ripple occurrence during stimulation, consistent with enhanced recruitment of hippocampal population events (Fig. 4a–e). These events occurred in a quiescent, low-arousal brain state, in which hippocampal SWRs are classically associated with replay and memory consolidation. This interpretation is supported by animal studies showing that selective disruption of hippocampal ripples impairs spatial memory, although event rate alone does not establish that memory replay or consolidation has occurred^6,34^.

**Figure 4.**
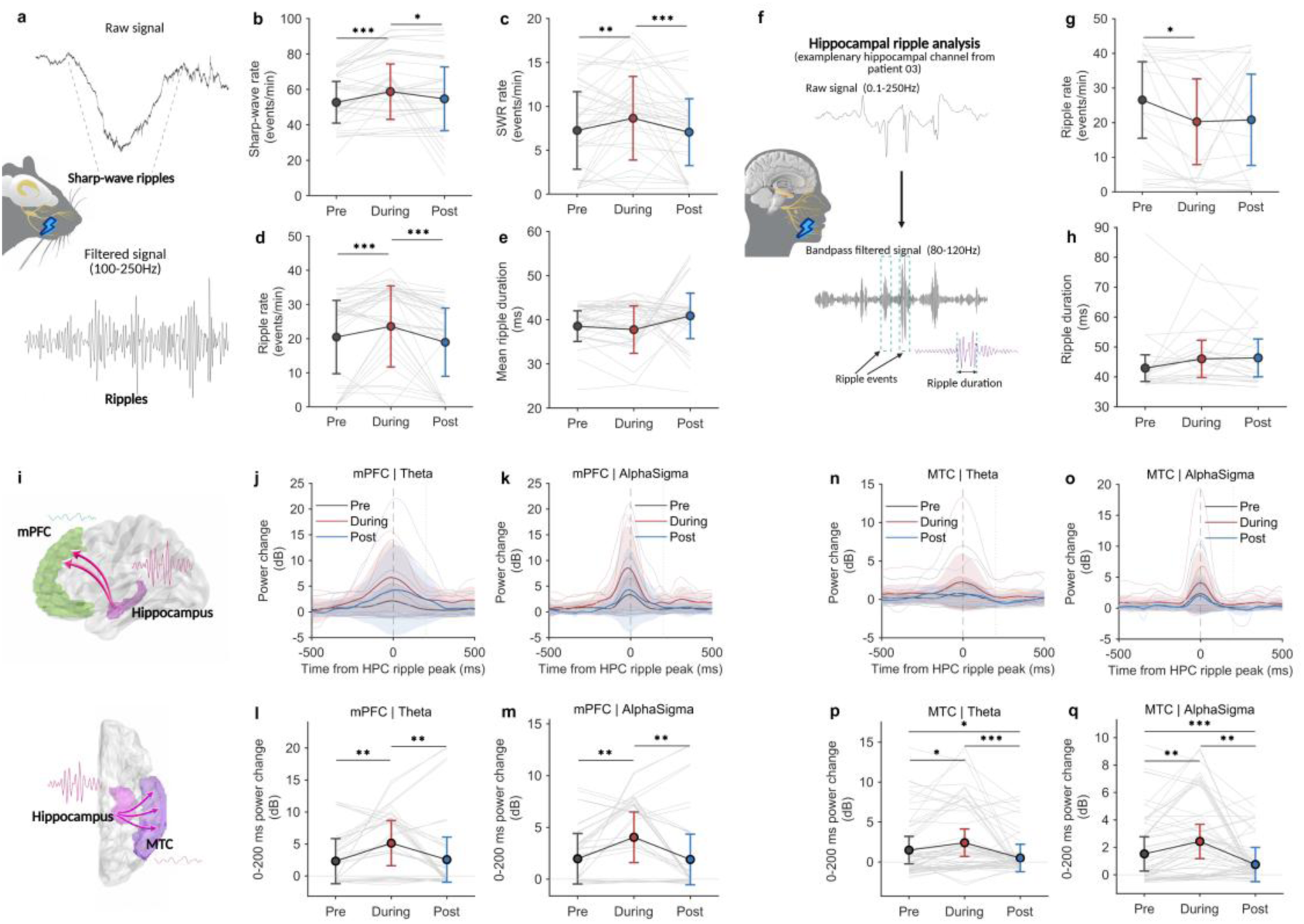
TNS-associated modulation of sharp-wave/ripple activity and ripple-locked cortical power. **a.** Representative in vivo hippocampal electrophysiological recordings from anaesthetized rats, showing the raw signal and corresponding filtered traces used to identify sharp waves and 100–250-Hz ripple events. **b–e.** Paired comparisons across the pre-stimulation, stimulation and post-stimulation periods of sharp-wave rate **(b)**, sharp-wave-ripple (SWR) rate **(c)**, ripple rate **(d)** and mean ripple duration **(e)**. **f.** Example hippocampal ripple analysis from patient 03, showing the raw signal, the 80–120-Hz band-pass-filtered signal, detected ripple events and ripple duration. **g,h.** Summary comparisons of ripple rate and ripple duration, respectively. **i.** Three-dimensional reconstructions showing the anatomical locations of contacts included in the ripple and ripple-coupled power analyses. **j,k,n,o.** Ripple-locked changes in theta and alpha–sigma power in the medial prefrontal cortex (mPFC) and medial temporal cortex (MTC), aligned to the hippocampal (HPC) ripple peak. The lower-row summary plots **l,m,p,q** show mean power changes during the 0–200-ms interval following the HPC ripple peak for the corresponding region–frequency combinations. In b–e,g,h and the summary plots, thin grey lines indicate hippocampal ripple channel level observations, whereas colored symbols and error bars indicate rats/subjects’ level means and 95% confidence intervals. In j,k,n,o, shaded envelopes indicate 95% confidence intervals. Asterisks denote significant pairwise contrasts (*P < 0.05, **P < 0.01 and ***P < 0.001); exact sample sizes, statistical tests and correction procedures are reported in the Methods. Pre, During and Post denote the pre-stimulation, stimulation and post-stimulation periods.

The human recordings showed a different pattern of ripple modulation. During i200-TNS, hippocampal ripple rate was modestly reduced relative to the pre-stimulation period, whereas ripple duration was numerically longer during stimulation (Fig. 4f–h). Thus, the human response was not characterized by a uniform increase in the number of ripple events. Instead, i200-TNS appeared to alter the balance between ripple occurrence and the temporal persistence of individual events. Ripple-locked analyses further showed transient changes in theta- and alpha/sigma-band power in the mPFC and MTC after the hippocampal ripple peak, indicating that i200-TNS modified hippocampal–cortical coupling during the stimulation period (Fig. 4i–q).

The divergent rate effects across preparations are consistent with a brain-state-dependent interpretation. In anesthetized animals, an increase in SWR and ripple occurrence is compatible with enhanced offline hippocampal activity related to memory replay and consolidation. In awake humans, by contrast, reduced ripple occurrence may reflect a shift toward a more aroused and externally engaged state. Recent human intracranial recordings indicate that hippocampal ripples are preferentially expressed during low-arousal states and are reduced during higher-arousal periods^18^. The locus coeruleus–noradrenaline system is a plausible contributor to this state transition because it regulates arousal and hippocampal memory-related network dynamics^16^. However, locus coeruleus activity and peripheral arousal indices were not directly measured in the human experiment. Therefore, the human ripple-rate reduction should be interpreted as being consistent with increased arousal-related neuromodulation, rather than as direct evidence of an LC-mediated mechanism.

### 2.5. i200-TNS enhances consolidation memory in face-name-occupation associative memory

To evaluate the effects of the i200-TNS protocol on associative memory performance in humans, a face–name-occupation associative memory task was used across two sessions in a randomized, crossover, double-blinded design (Fig. 5a and b). Each session included either i200-TNS or sham stimulation applied in both the encoding and consolidation phases, with a washout interval of at least two days between sessions (sham vs i200-TNS). Each session in the same participant used completely different sets face, name and occupation to avoid any practice or carry-over effect. During each session, participants learned associations between unfamiliar faces, names, and occupations. Memory performance was assessed with a short-term recall test 2 minutes after encoding and a long-term recall test after 20 minutes consolidation. Stimulation effects on the encoding and consolidation phases were quantified as changes in recall accuracy between the active and sham conditions, measured before and after each respective phase.

**Figure 5.**
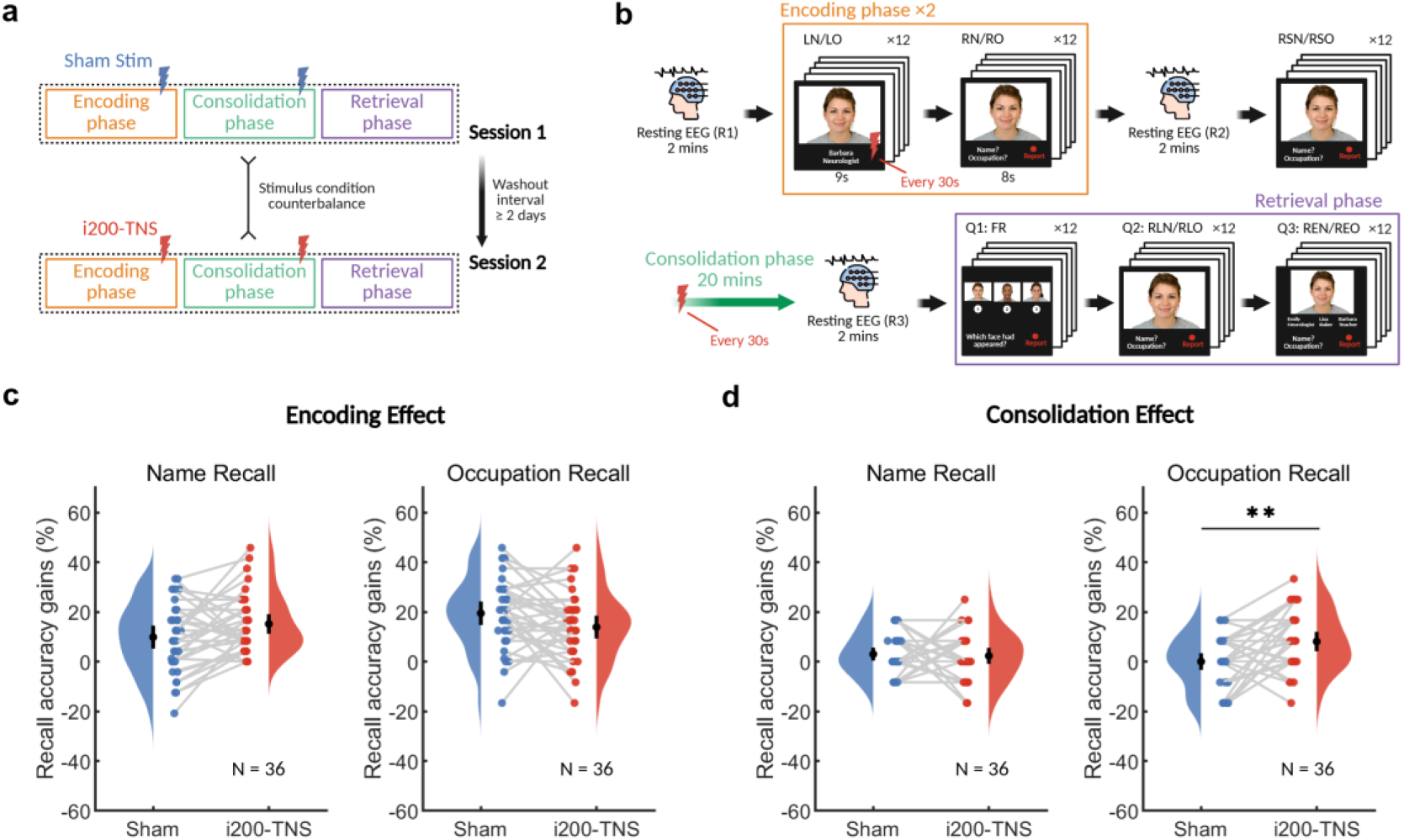
Experimental design and behavioral outcomes of the face–name associative memory task. **a.** Schematic of the experimental protocol. Each participant completed two sessions in a randomized, counterbalanced order, separated by a washout interval of at least two days. Each session consisted of an encoding phase, a consolidation phase, and a retrieval phase. Sham stimulation or intermittent 200-Hz external trigeminal nerve stimulation (i200-TNS) was delivered during the encoding and consolidation phases. EEG was recorded throughout each session. **b.** Structure of the face–name-occupation associative memory task. During encoding, participants learned 12 unfamiliar faces, each paired with a name and an occupation, and were instructed to memorize both associations. Each learning block was followed by a reinforced recall block, in which participants verbally reported the associated name and occupation for each face. After a 2-min resting EEG recording, short-term memory was assessed. Following a 20-min consolidation phase and another resting EEG recording, participants completed the retrieval phase, which included three tasks: face recognition (FR), free recall of the associated name and occupation (RLN/RLO), and cued recognition of the associated name and occupation from three options (REN/REO). **c,d.** Behavioral effects of i200-TNS compared with sham stimulation. Recall accuracy gains are shown for name and occupation recall following stimulation during the encoding phase (c) and consolidation phase (d). Colored dots indicate individual participants, gray lines indicate within-subject paired comparisons, and black dots with error bars indicate group means with 95% confidence intervals. i200-TNS significantly improved occupation recall when delivered during the consolidation phase. Asterisks indicate significance after correction for multiple comparisons: **p < 0.01.

When i200-TNS was applied during the encoding phase, short-term name recall showed a modest increase under i200-TNS compared to sham (Fig. 5c left panel), although the difference did not reach statistical significance (paired t-test, t(35) = -2.2650, p = 0.1192 (Bonferroni corrected), Cohen’s d = -0.3775). Occupation recall also showed a non-significant pattern (Fig. 5c right panel; t(35) = 2.1061, p = 0.1698 (Bonferroni corrected), Cohen’s d = 0.3510). In the long-term recall test, participants who received i200-TNS during both the encoding and consolidation phases showed a significant enhancement in occupation recall relative to the sham condition (Fig. 5d right panel; t(35) = -3.4009, p = 0.0068 (Bonferroni corrected), Cohen’s d = -0.5668). Long-term name recall did not differ between i200-TNS and sham (Fig. 5d left panel; t(35) = 0.3007, p = 1 (Bonferroni corrected), Cohen’s d = 0.0501). Shapiro–Wilk tests confirmed normality for all comparisons (p > 0.05), and paired t-tests were used accordingly. All p-values were adjusted using the Bonferroni correction by multiplying the raw p value by four (comparisons).

### 2.6. i200-TNS modulates EEG dynamics during associative memory processing

During the face–name-occupation associative memory task, we simultaneously recorded 64-channel scalp EEG to examine how i200-TNS modulates cortical activity across different stages of memory processing. Three EEG outcomes were analyzed: (i).stimulation-evoked potentials (EPs) during the consolidation phase, (ii).visual evoked potentials (VEPs) elicited by face presentation during memory retrieval and (iii).resting-state spectral power measured across three resting periods. By relating these EEG measures to behavioral performance, we aimed to identify cortical signatures associated with the memory-enhancing effect of i200-TNS.

During the 20-min stimulation phase, i200-TNS elicited robust evoked potentials across widespread brain regions (Fig. 6a). The response was characterized by an early temporal deflection followed by a sustained negative component, most clearly observed over bilateral temporal electrodes, including T7 and T8. Given the anatomical proximity of these scalp sites to medial temporal structures, such temporal responses may reflect the engagement of memory-related cortical–subcortical networks ^35,36^. Topographical maps further showed a dynamic propagation of the evoked response, suggesting that i200-TNS recruited a distributed neural network during the consolidation period.

**Figure 6.**
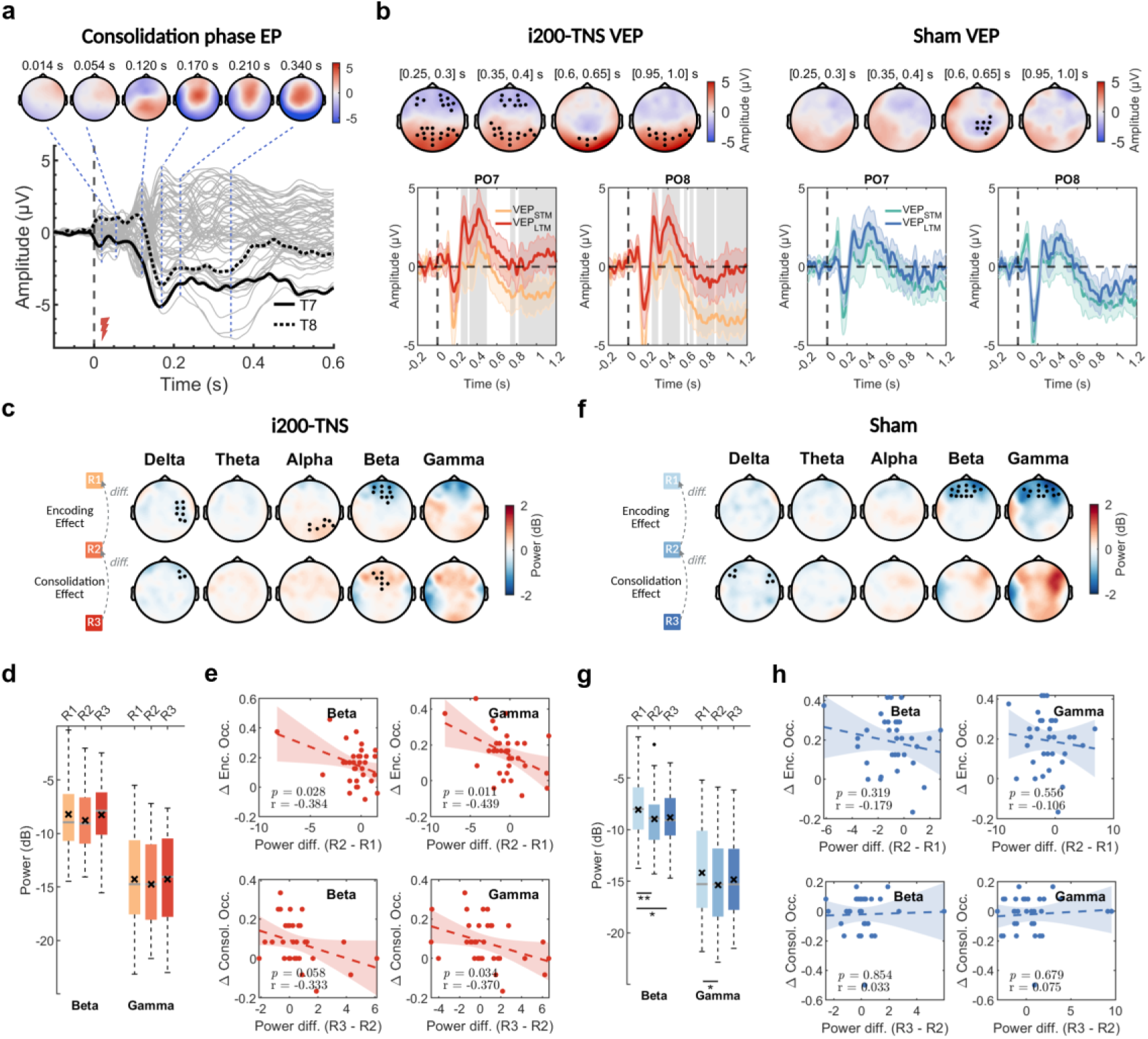
EEG correlates of i200-TNS stimulation and their relationship to memory performance. **a.** Evoked potentials recorded during the consolidation phase following i200-TNS. Bottom: grand-averaged evoked responses across all electrodes, with representative waveforms from bilateral temporal electrodes T7 and T8 highlighted. Stimulation onset occurred at 0 s and lasted for 1 s. Top: scalp topographies showing the temporal evolution of low-frequency evoked activity at representative post-stimulation time points. **b.** Visual evoked potentials during short-term memory retrieval before stimulation and long-term memory retrieval after stimulation in the i200-TNS group and sham group. Top: scalp topographies of VEP differences across representative time windows. Significant electrodes identified by cluster-based permutation testing are marked in black. Bottom: VEP time courses from representative parieto-occipital electrodes PO7 and PO8. Shaded regions indicate significant STM–LTM differences. **c.** Resting-state spectral power changes in the i200-TNS group across delta (0.4-4 Hz), theta (4-8 Hz), alpha (8-13 Hz), beta (13–30 Hz) and gamma (30–90 Hz) bands. The upper row shows power differences from R1 to R2, reflecting encoding effect, whereas the lower row shows power differences from R2 to R3, reflecting consolidation effect. Significant electrodes are marked in black. **d.** Average beta and gamma power across significant left-frontal electrodes during R1, R2, and R3 in the i200-TNS group. **e.** Correlations between left-frontal beta/gamma power changes and individual memory performance gains in the i200-TNS group. Dashed lines indicate linear regression fits, and shaded areas indicate 95% confidence intervals. **f-h.** Corresponding resting-state spectral power analyses for the sham group. Asterisks indicate statistical significance after correction for multiple comparisons: **p < 0.01, *p < 0.05.

We next examined whether i200-TNS altered neural responses during memory retrieval by comparing VEPs between the short-term memory test before 20-min stimulation and the long-term memory test after 20-min stimulation. In the i200-TNS group, cluster-based permutation tests revealed significant differences between STM and LTM VEPs over occipital and frontal electrodes (Fig. 6b, left). These differences emerged approximately 200 ms after face onset and extended into later time windows, particularly over parieto-occipital regions. In contrast, the sham group showed weaker and less spatially extensive STM–LTM differences (Fig. 6b, right), suggesting that the retrieval-related cortical modulation was more pronounced following active i200-TNS.

Resting-state EEG further revealed stimulation-related changes in spectral power. In the i200-TNS group, beta and gamma power showed moderate reductions from baseline to the stimulation period, with effects primarily distributed over left-frontal electrodes (Fig. 6c, d).

These reductions were generally smaller than those observed in the sham group, where beta and gamma power decreased more prominently across the same resting stages (Fig. 6f, g). This pattern suggests that i200-TNS may attenuate the decline in higher-frequency frontal activity during the consolidation interval.

Finally, we tested whether individual differences in spectral modulation were associated with behavioral memory gains. In the i200-TNS group, smaller reductions in left-frontal beta and gamma power during the stimulation phase were associated with larger encoding-related memory benefits, as indicated by negative correlations between power changes from R2 to R1 and memory improvement (beta: r = –0.384, p = 0.028; gamma: r = –0.439, p = 0.011; Fig. 6e). Similar negative associations were also observed for consolidation-related gains from R3 to R2, particularly in the gamma band (gamma: r = –0.370, p = 0.034). In contrast, no significant correlations were found in the sham group (Fig. 6h), indicating that the relationship between frontal high-frequency power preservation and memory enhancement was specific to active stimulation.

Across the EEG analyses, active stimulation elicited robust cortical responses during consolidation, enhanced late retrieval-related VEP components after stimulation, and attenuated beta/gamma power reductions over left-frontal regions. Importantly, the preservation of frontal beta and gamma activity was associated with larger memory gains, supporting a link between i200-TNS-induced cortical modulation and improved associative memory performance.

## 3. Discussion

This study shows that temporal patterning is a key determinant of the hippocampal response to trigeminal nerve stimulation across species. In anaesthetized rats, i200-TNS produced sustained CA1 synaptic potentiation and increased neuronal firing. LC inhibition prevented these sustained responses, whereas local D1/D5 receptor blockade altered both synaptic and spiking activity, with partial recovery of neuronal firing after stimulation. Translation of this optimized protocol to humans revealed robust hippocampal and thalamic responses, increased hippocampal theta–gamma phase–amplitude coupling and brain-state-dependent modulation of ripple-related activity. In healthy adults, i200-TNS selectively improved long-term occupation recall and altered retrieval-related and resting-state EEG measures. These findings establish i200-TNS as a mechanistically informed neuromodulation approach with cross-species evidence supporting its translational potential for engaging hippocampal memory networks.

### i200-TNS facilitates hippocampal plasticity

i200-TNS induced a lasting increase of hippocampal spiking activity and fEPSP slope that surpassed the canonical >20% criterion for LTP (Fig.1c). However, c100-TNS failed to elicit sustained post-synaptic potentiation (Fig.1e). Our finding challenges the efficiency of conventional tonic pulse trains in cognitive modulation and showed that the hippocampus not only serves as a promising downstream target for trigeminal nerve afferents but also benefits from temporally structured extracranial stimulation through enhanced synaptic plasticity. Notably, i200-TNS produced these effects while delivering substantially less cumulative charge than c100-TNS. The difference therefore cannot be explained by greater delivered charge alone. The intermittent burst design may better engage neurotransmitter release that gate plasticity. Prior work in our team has shown rodent^19,20^ and human evidence that TNS^37^ could activate locus coeruleus (LC), which supply noradrenaline (NE) and dopamine (DA) to the hippocampus. Both NE and DA are indispensable for LTP persistence, enhancing NMDA receptor–mediated calcium influx and downstream signaling cascades^38–40^. LC neurons operate through both tonic and phasic firing modes, with phasic activity producing temporally concentrated catecholamine release that can promote hippocampal plasticity ^41^. The pauses between successive bursts may permit transmitter clearance and vesicle replenishment, thereby preserving NE and DA signalling across repeated activations^42,43^. Accordingly, the intermittent structure of i200-TNS may be better suited than continuous stimulation to coordinate LC-dependent neuromodulation with afferent input to CA1. Our pharmacological experiments provide direct support for this mechanism: local inhibition of the LC prevented the sustained increase in CA1 fEPSP slope and the prolonged elevation in hippocampal firing normally observed after i200-TNS. By contrast, local CA1 D1/D5 receptor blockade altered both synaptic and spiking responses less completely, with partial recovery of neuronal firing after stimulation. This difference suggests that hippocampal D1/D5 signalling contributes to the i200-TNS response but does not fully account for the broader effect of LC activity, which is likely to involve coordinated dopaminergic and noradrenergic modulation.

This interpretation is consistent with a broader principle in neuromodulation: patterned stimulation (e.g., intermittent theta-burst rTMS and tACS) often produces stronger and more durable plasticity than tonic stimulation^44–46^. In our experiments, i200-TNS induced persistent synaptic potentiation and neuronal firing despite delivering approximately 15-fold less cumulative absolute charge than c100-TNS (≈0.0008 C versus ≈0.012 C). Continuous 100-Hz stimulation may instead impose a more tonic drive that is less favourable for durable potentiation and more susceptible to synaptic fatigue,^47–49^. Moreover, LC-dependent catecholaminergic modulation commonly follows an inverted-U relationship^50–52^, such that moderate, temporally structured recruitment may be more effective than sustained high-intensity drive.^53^. These findings indicate that the efficacy of i200-TNS is determined not simply by delivered charge, but by a temporal pattern that recruits LC-dependent neuromodulatory mechanisms conducive to hippocampal plasticity.

### Brain-state-dependent ripple modulation and ripple-locked cortical activity

Hippocampal sharp-wave ripples are highly state-dependent population events whose occurrence and functional context vary with vigilance and behavioural engagement ^6,7^. In urethane-anaesthetized rats, i200-TNS transiently increased sharp-wave (Fig.4b), sharp-wave– ripple (Fig.4c) and ripple rates (Fig.4d) during stimulation, whereas mean ripple duration was unchanged (Fig.4e). Urethane anaesthesia preserves spontaneous alternations between activated and deactivated sleep-like forebrain states^54^, and hippocampal ripples remain embedded within coordinated hippocampal–neocortical activity under this preparation^55^. The transient increase in event rate therefore indicates that i200-TNS recruited synchronous hippocampal population events in a quiescent, anaesthetized state. However, because neither replay content nor subsequent memory was measured in these animals, increased ripple occurrence alone should not be interpreted as evidence of enhanced replay or consolidation.

In awake patients, i200-TNS produced a different response: hippocampal ripple rate was modestly reduced during stimulation, whereas mean ripple duration did not change significantly. The difference between rats and humans is plausible given the strong dependence of ripple occurrence on brain state. Recent human intracranial evidence suggests that ripples are more prevalent during low-arousal states and become less frequent as arousal increases^18^, providing one possible explanation for the reduction observed during sensory stimulation. However, awake ripples can also be recruited in behaviourally salient or emotionally arousing contexts, indicating that ripple rate is not a monotonic index of either arousal or memory processing^56^. Although pupillary and other arousal measures were not acquired in the sEEG cohort, the reduction in human ripple rate is compatible with a stimulation-related shift in arousal or network state. This interpretation is supported by our previous^19,20^ and present animal data, which identify LC recruitment as a key component of the hippocampal response to TNS and provide a plausible mechanism through which stimulation may engage ascending arousal systems.

A complementary network-level finding was that hippocampal ripples were followed by stronger theta- and alpha/sigma-band responses in the mPFC and MTC during i200-TNS, particularly within the first 200 ms after the ripple peak (Fig.4j-q), and these changes did not persist into the post-stimulation period. Previous intracranial and multisite recordings have shown that hippocampal ripples are temporally coordinated with association cortex and can organize widespread cortical activity during both sleep and wakefulness^57,58^. Our findings are therefore consistent with i200-TNS altering the temporal alignment of hippocampal events with cortical low-frequency responses. Because these recordings were obtained during wakefulness, the 4–13-Hz activity should not be interpreted as a sleep spindle, and ripple-triggered power does not establish the direction of communication or the transfer of memory content. Ripple-triggered cortical power may therefore serve as a physiological readout of hippocampal–cortical network engagement during stimulation^58^.

### Human associative-memory enhancement and convergent electrophysiological signatures

The behavioral experiments provide direct evidence that i200-TNS can drive functional memory gains in humans. In a controlled face–name-occupation associative memory task, i200-TNS during the consolidation phase improved recall accuracy (Fig.5d), whereas i200-TNS during encoding had no reliable effect on recall accuracy gain after stimulation (Fig.5c). This phase specificity strongly supports the mechanistic interpretation that i200-TNS acts on post-encoding stabilization processes, consistent with the electrophysiological signatures of enhanced LTP and sharp-wave ripple rate found in rodent. It is widely accepted that memory formation is based on activity-dependent increases in synaptic strength, as predicted by Cajal and formalized by Hebb, with LTP serving as the principal experimental model linking such synaptic potentiation to information storage^3^. We found that i200-TNS selectively enhanced occupation recall during retrieval tasks, with no measurable effect on name recall (Fig.5d). The selective benefit for occupation recall may reflect differential semantic demands: occupations are common nouns with richer associative structure^59^, whereas proper names depend on more fragile, specialized retrieval pathways^60^. This difference likely makes occupation information more amenable to consolidation across hippocampal–neocortical networks^61^.

The sEEG recordings provide direct evidence that i200-TNS engages the human hippocampus. Individual i200-TNS bursts evoked prominent responses in the hippocampus and thalamus, whereas neighboring peri-hippocampal white-matter contacts showed no consistent stimulus-locked response. Beyond this evoked response, hippocampal theta–gamma phase–amplitude coupling (PAC) increased after burst offset (Fig.3h,3j). Theta–gamma PAC provides a candidate temporal mechanism through which local gamma-band activity is organized by slower hippocampal rhythms^31,62^. Previous rodent^32^ and human studies^33,63^ have linked this organization to associative learning, successful episodic encoding and the temporal ordering of mnemonic representations. The post-burst PAC increase extended beyond the immediate evoked response and was followed by a transient change in hippocampal network timing. PAC and behavioral performance were measured in separate cohorts, so we could not test whether the PAC change mediated the memory effect. Even so, the PAC result provides independent physiological evidence that i200-TNS engages hippocampal network activity relevant to memory.

In the healthy cohort, scalp EEG showed concurrent changes in distributed cortical activity. During the consolidation interval, i200-TNS elicited robust stimulation-locked potentials with prominent temporal and broader scalp distributions (Fig.6a). During subsequent retrieval, the active condition showed more pronounced and spatially extensive differences between short- and long-term visual evoked responses than the sham condition, particularly over occipital and frontal regions in later post-stimulus windows (Fig.6b). These late responses occurred within a time range associated with attentional allocation, mnemonic updating and retrieval-related evaluation ^64,65^. Additionally, in the sham condition we observed decreases in resting-state beta and gamma power (Fig.6g), which could be explained with rising sleepiness/fatigue over the session^66,67^ and with time-dependent event-related desynchronization linked to heightened cognitive control demands^68,69^. However, i200-TNS stabilized frontal beta and gamma power (Fig.6d). These frequencies have been linked to active maintenance and top-down control^70^, and their preservation was associated with individual memory gains^71^. The attenuated decline in beta under i200-TNS could also be explained with a more alert brain state driven by sensory afferent pathways of TNS, in line with our human evidence with decreased ripple rate and prior TNS studies^37,72^. The human recordings therefore show concurrent hippocampal and cortical effects of i200-TNS during the memory experiment. The present data do not establish a directional influence from hippocampus to cortex.

### Limitations and future directions

We acknowledge several limitations that point to clear avenues for future work. First, while we established LTP-like plasticity in rodents and memory benefits in humans, we did not directly access memory behavior in awake animals, leaving a gap between physiological and behavioral endpoints. The cross-species design partly addresses this gap, but it does not replace behavioral testing in awake animals. Second, our human behavior experiments focused on short-term (1-2 hour) consolidation with single TNS session, whether i200-TNS promotes longer-term memory retention over days or weeks remains to be determined. Future studies should extend longer stimulation protocols and follow-up tests to evaluate memory maintenance. Third, stimulation parameters were chosen to mimic invasive classical LTP induction, but systematic variation of frequency, duty cycle, and duration could identify protocols with greater efficacy or specificity. Finally, the human intracranial data were obtained in epilepsy patients, whose brain networks may differ from healthy populations. However, the convergence with scalp EEG and behavioral findings in healthy participants suggests that the observed effects are not disease specific.

## Conclusion

In this work we offer extensive evidence that temporally structured TNS may promote hippocampal plasticity and enhance memory consolidation, using rodent electrophysiology, human brain recordings, and behavioral experiments. Unlike conventional protocols, i200-TNS is designed around fundamental principles of classic synaptic plasticity induction, enabling peripheral stimulation to access central memory circuits in a non-invasive way. These results clarify how patterned TNS can engage hippocampal circuits. Further studies are needed to determine whether the approach can produce durable benefits in ageing or memory disorders.

## 4. Methods

### 4.1 Animal experiment

#### 4.1.1 Ethics approval

Animal experiments were conducted using a total of 24 adult male Sprague-Dawley rats (250–400 g; Charles River Laboratories). All procedures adhered to protocols approved by the KU Leuven Animal Ethics Committee (protocol number P072/2020).

#### 4.1.2 Electrophysiological recording

To record field excitatory postsynaptic potentials (fEPSPs), adult male rats were anesthetized with an intraperitoneal injection of urethane at 1500mg/kg rat weight (Sigma-Aldrich, USA) and fixed in a stereotaxic frame (Narishige SR-6). A midline scalp incision was made to expose the skull, followed by a craniotomy with two burr holes drilled: one for the recording electrode at hippocampal CA1 coordinates AP = −3.6 mm and ML = 1.5 mm (inserted at a 52.5° angle), and another for the bipolar stimulating electrode at hippocampal CA3 Schaffer-collateral pathway coordinates AP = −4.2 mm and ML = −3.8 mm, inserted vertically (see Fig.1A). Both coordinates were relative to Bregma.

Monosynaptic fEPSPs were recorded using a single-shank, 32-channel silicon probe (E32+R-50-S1M-L20 NT, Atlas Neuro, Leuven, Belgium; vertical span: 1550 μm). The probe was lowered to a depth of 2.8–3.4 mm from the dura, targeting the CA1 region. Signals were amplified (×192), bandpass filtered between 0.1 Hz and 7.9 kHz, and digitized at 30 kHz with 16-bit resolution using an RHD32 Intan headstage (Intan Technologies, USA) connected to the Open Ephys acquisition system (www.openephys.org). Real-time feedback allowed for identification of hippocampal CA1 spike bursts and CA3 sharp-wave events (see Fig.2A).

The bipolar stimulating electrode (Invilog Research Ltd., Finland), connected to a battery-powered, low-noise stimulus isolation unit, was inserted to a depth of 3.0–3.8 mm (DV from skull surface) and adjusted until a reliable “sharped U-shaped” fEPSP waveform was observed in the CA1 region (see Fig.1B). Biphasic square-wave pulses (pulse width: 200 μs) were delivered every 30 seconds for fEPSP measurement. Input–output (I/O) curves were constructed by progressively increasing stimulation intensity until the fEPSP amplitude plateaued. The stimulation strength was then set to evoke 50% of the maximal fEPSP amplitude and kept consistent throughout the experiment.

Following a stable 30-minute baseline recording, a 5-minute TNS intervention was administered, after which fEPSPs were continuously recorded for 90 minutes.

#### 4.1.3 TNS protocol in animal experiment

In the animal experiments, i200-TNS and c100-TNS were delivered via two disposable subdermal needle electrodes (0.4 mm diameter; Technomed, USA) connected to a second stimulus isolation unit (same model as described above). To ensure stable subcutaneous placement, the electrodes were manually bent into a fishhook shape. One electrode was positioned to hook around the distal branch of the mandibular division of the trigeminal nerve (stimulation site 1, SS1), while the second electrode was inserted proximally along the same nerve branch, near the trigeminal ganglion (stimulation site 2, SS2). SS1 was connected to the anodal output (inward current flow relative to the skin surface), and SS2 to the cathodal output (outward current flow relative to the underlying grey matter).

Stimulation parameters were programmed in MATLAB and delivered through the stimulus isolation unit. Two stimulation protocols were tested: 1) i200-TNS condition: Trains of biphasic square pulses (200 Hz, 200 μs pulse width) at 1 mA were delivered in 1-second bursts, followed by a 29-second inter-train interval. The stimulation lasted for 5 minutes, comprising a total of 10 trains. 2) c100-TNS condition: A continuous 5-minute train of biphasic square pulses at 1 mA was applied at 100 Hz (pulse width: 200 μs).

#### 4.1.4 Pharmacological blockade experiment procedure

Pharmacological blockade experiments were performed in the same anesthetized in vivo CA1 recording preparation described above to test whether LC activity and hippocampal D1/D5 receptor signaling contributed to i200-TNS-induced fEPSP potentiation. Separate cohorts of rats received a local infusion of either clonidine into the locus coeruleus (LC) or the D1/D5 receptor antagonist (+)-SCH 23390 into dorsal hippocampal CA1 before i200-TNS. Drugs were dissolved in artificial cerebrospinal fluid (aCSF; EcoCyte Bioscience, USA) and prepared on the day of recording.

For LC blockade, clonidine was used to suppress LC activity through activation of presynaptic alpha-2 adrenergic receptors, following the local LC blockade procedure described previously^20^. A burr hole was drilled above the LC target at AP = -4.2 mm from lambda and ML = 1.2 mm, and a Hamilton microsyringe (1750RN, Hamilton Co., USA) was inserted at a 15° posterior angle to a depth of DV = 5.5 mm from the dura. A total dose of 4 μg clonidine was infused slowly over 10 min. For hippocampal D1/D5 receptor blockade, (+)-SCH 23390 was infused into dorsal CA1 at AP = -3.6 mm, ML = 2.3 mm and DV = 2.2 mm from the dura, with coordinates defined relative to Bregma. SCH 23390 was prepared at 5 μM, consistent with prior CA1 LTP experiments using D1/D5 receptor blockade^73^.

After electrode placement and input-output curve calibration, fEPSPs were evoked every 30 s as described above. A stable 30-min baseline was recorded before pharmacological manipulation. After infusion, recordings were continued for a 30-min post-infusion blockade period. Rats then received the same 5-min i200-TNS protocol described in section 4.1.3, consisting of 1-s trains of 200-Hz biphasic pulses delivered every 30 s for 10 trains. Post-stimulation fEPSPs and spontaneous spiking activity were recorded for 90 min.

Each animal was assigned to one pharmacological manipulation. Analyses were aligned to the following experimental epochs: 30-min baseline, 30-min post-infusion blockade period, 5-min i200-TNS and 90-min post-stimulation recording. fEPSP slope and spike-rate analyses were performed using the same preprocessing, normalization and statistical procedures described below.

#### 4.1.5 Electrophysiologic signal processing, analysis and statistics

##### 4.1.5.1 fEPSP data analysis and statistics

fEPSPs were analyzed offline using customized MATLAB scripts. Local field potential (LFP) recordings were preprocessed to exclude noisy or non-functional channels, apply a 50 Hz notch filter, and remove stimulation artifacts. Signals were then bandpass filtered between 0.1 and 300 Hz. Stimulus-locked epochs were extracted from 0 to 30 ms following each stimulation pulse. The fEPSP amplitude was defined as the minimum (trough) voltage within the 0–30 ms post-stimulation window. The fEPSP slope was estimated as the steepest negative deflection (measured as its maximum absolute slope) within the 5% to 95% descending phase of the trough ^74^, which is in line with the typical timing of Schaffer collateral-evoked responses. For each animal, the channel showing the largest average fEPSP amplitude during baseline was selected for further analysis. Within that channel, evoked responses were averaged in non-overlapping 10-minute bins, each consisting of 20 responses recorded at 30-second intervals. The average fEPSP slope during the first 10 minutes of recording was used as the baseline reference and defined as 100%. All subsequent slopes were normalized to this value to visualize changes in synaptic strength over time.

To evaluate stimulation-induced changes, we used linear mixed effect (LME) model to compare the normalized slope of each 10-minute bin to the baseline, with rat number as random effect and time epoch as fixed-effect (MATLAB, fitlme.m). We also used an LME model to compare the original fEPSP slopes between the 30-min baseline and 30-min post-stimulation periods within each experimental group, with time epoch (baseline versus post-stimulation) as a fixed effect and rat identity as a random intercept. Group-specific sample sizes are reported in the corresponding Results and figure legends. Model syntax using the Wilkinson notation was as such: fEPSPslope∼ 1 + timeEpoch+ (1 | ratNum). ANOVA was performed on the model output to test the significance of the fixed-effects and interactions. ANOVA F-statistics and p-values for fixed-effects and interactions are reported in Results. If the main effect of time epoch was significant, 11 planned post hoc linear contrasts compared each of the remaining time epochs with the first baseline epoch (−30 to −20 min). P values were Bonferroni-adjusted across the 11 contrasts.

##### 4.1.5.2 Spike rate analysis and statistics

Spike sorting was performed on the raw data obtained from the multi-channel probe using SpykingCircus, followed by manual curation in Phy (https://github.com/cortex-lab/phy) to guarantee the selection of well-isolated single units. We extracted spike timings and calculated spike rates from these curated units for the 12 time epochs (3 for baseline and 9 for post-stimulation) independently. We used LME model to compare the spike rate of each 10-minute bin to the first baseline, with neuron number and rat number as random effect and time epoch as fixed-effect (MATLAB, fitlme.m). Model syntax in Wilkinson notation was: spikeRate ∼ 1 + timeEpoch + (1 | ratNum) + (1 | unitID). The overall effect of time epoch was evaluated using ANOVA on the fitted model. When this effect was significant, 11 planned linear contrasts compared each remaining epoch with the first baseline epoch (−30 to −20 min), with P values Bonferroni-adjusted across the 11 contrasts within each stimulation condition.

##### 4.1.5.3 Sharp-wave ripple detection and analysis

Hippocampal sharp-wave ripple (SWR) events were detected from 32-channel silicon-probe recordings using a custom MATLAB pipeline adapted from established protocols (https://github.com/buzsakilab/buzcode/tree/master/analysis/SharpWaveRipples) for ripple analysis in rodents ^6^. Continuous Open Ephys recordings were sampled at 30 kHz. Recordings were analysed separately during the pre-stimulation, stimulation and post-stimulation periods, corresponding to 10 min, 5 min and 10 min epochs, respectively. Stimulation pulse artefacts were removed before event detection using the epoch-specific artifact-removal mode implemented in the pipeline. Common-average referencing was not applied in this analysis.

The ripple channel was defined by manual screening using the Open Ephys GUI RippleDetector plugin (https://github.com/bmsousa91/RippleDetector) ^75^, with the same detection strategy. Contacts showing clear negative-going hippocampal sharp-wave deflections accompanied by ripple-band activity were retained for the present analysis. This manual screening step was used only for contact selection. Event detection and quantification were subsequently performed automatically using the predefined criteria described below.

For each selected channel and epoch, the signal was filtered using a second-order zero-phase Butterworth band-pass filter. Sharp waves were detected in the 2–50 Hz band. The normalized sharp-wave signal was z-scored using the mean and standard deviation of the corresponding channel and epoch. Candidate sharp-wave events were defined as contiguous periods exceeding 1.5 standard deviations. Events were retained when their peak z-score exceeded 2.5 standard deviations and their duration was between 20 and 250 ms. The sharp-wave peak was defined as the maximum of the polarity-corrected z-scored sharp-wave signal. Sharp-wave amplitude was extracted from the signed 2–50 Hz filtered signal at this peak.

Ripples were detected from the same channel using a 100–250 Hz second-order zero-phase Butterworth filter. The filtered signal was squared and smoothed using a 10-ms moving-average window. The resulting ripple-band power was z-scored using the mean and standard deviation of the corresponding channel and epoch. Candidate ripple events were defined as contiguous periods exceeding 2 standard deviations. Candidate segments separated by less than 30 ms were merged. Events were retained only when their normalized ripple-power peak exceeded 3.5 standard deviations and their duration was between 20 and 200 ms. The ripple peak was defined as the time of maximum normalized ripple-band power within the retained event. Ripple onset and offset were defined by the boundaries of the low-threshold candidate segment.

Sharp-wave-associated ripple events were identified by coupling each ripple to the temporally nearest sharp-wave peak. A ripple was considered coupled to a sharp wave when its peak occurred within the sharp-wave interval or within 100 ms of the sharp-wave peak. The coupling lag was calculated as ripple peak time minus sharp-wave peak time, with positive values indicating that the ripple peak followed the sharp-wave peak. Each channel and epoch produced event counts, event rates and event-level feature summaries. Sharp-wave rate, ripple rate and SWR rate were calculated as the corresponding event count divided by epoch duration and expressed as events per minute. SWR rate was defined as the number of ripples coupled to a sharp wave per minute. Ripple-coupled fraction was calculated as the number of sharp-wave-coupled ripples divided by the total number of ripples. Sharp-wave-coupled fraction was calculated analogously as the number of sharp waves associated with at least one ripple divided by the total number of sharp waves.

Additional event-level metrics included mean sharp-wave duration, mean ripple duration, mean sharp-wave peak z-score, mean signed sharp-wave amplitude, mean ripple peak z-score and mean ripple-to-sharp-wave peak lag. Event-conditional metrics were calculated as the mean across valid events within each channel and epoch. When no events were detected, event rates were assigned a value of zero, whereas event-conditional means were treated as missing and excluded from subsequent model fitting.

For visualization, channel-level values were retained as paired observations across the three epochs. Rat-level values were obtained by first averaging all selected channels within each rat and epoch, followed by averaging across rats. The inferential analyses reported here focused on four primary metrics: sharp-wave rate, sharp-wave–ripple (SWR) rate, ripple rate and mean ripple duration. Sharp-wave-coupled fraction was retained as an additional descriptive metric.

For inferential analysis, channel-by-epoch observations were assembled into a long-format table. Each metric was analysed using a linear mixed-effects model fitted by maximum likelihood in MATLAB. The model was:

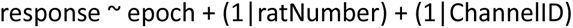

Here, epoch was treated as a categorical fixed effect with Pre, During and Post levels. ratNumber and the unique channel identifier were included as random intercepts, with channels therefore treated as repeated measurements nested within rats. Marginal ANOVA assessed the overall effect of epoch. Planned contrasts compared During versus Pre, Post versus Pre and During versus Post. Contrast estimates, standard errors, test statistics, denominator degrees of freedom, 95% confidence intervals and uncorrected P values were exported. P values were Bonferroni-corrected across the three planned epoch contrasts within each metric. A model-standardized effect size was also calculated by dividing each contrast estimate by the model residual standard deviation.

### 4.2 Human experiments

#### 4.2.1 Ethics approval

A total of 37 healthy adults were recruited from the general population. One participant withdrew before completing the second experimental session and was excluded from the behavioral analysis, leaving 36 participants with complete behavioral data. This study was approved by the Ethics Committee Research UZ/KU Leuven (approval number S63709) and registered on ClinicalTrials.gov (NCT04577677). For the intracranial sEEG recordings, 9 patients with pharmacoresistant epilepsy undergoing clinical implantation of stereotactic electrodes were recruited at The Second Affiliated Hospital, Zhejiang University School of Medicine (SAHZU). This study was approved by the SAHZU Human Research Ethics Committee (approval number IH2020001047). All participants provided written informed consent prior to participation and received monetary compensation. All procedures involving human subjects were conducted in accordance with the Declaration of Helsinki (1964) and Good Clinical Practice guidelines.

#### 4.2.2 sEEG experiment

##### 4.2.2.1 sEEG data acquisition, processing

Stereotactic EEG (sEEG) recordings were acquired at The Second Affiliated Hospital of Zhejiang University School of Medicine using a Nihon Kohden EEG-1200C system, sampled at 2000 Hz. Preoperative high-resolution T1-weighted MRI and postoperative CT scans were obtained as part of standard clinical care and anonymized prior to analysis. Electrode contact positions were manually identified in native space by two independent neurologists based on individual scans. For anatomical reconstruction, MRI data were segmented using Freesurfer (v7.4.0) according to the Desikan–Killiany atlas, and co-registered with the corresponding CT using affine transformation. Electrode coordinates were extracted from the CT using the Brainstorm toolbox, and for visualization purposes, aligned electrode positions were normalized to MNI space and projected onto a template brain using BrainNet Viewer. All electrode localization and data analyses were performed in native space, and MNI-based projections were used exclusively for group-level visualization.

sEEG preprocessing was performed using a custom pipeline implemented in Python, incorporating functions from MNE-Python and SciPy libraries. Raw electrophysiological signals were acquired at 2000 Hz and segmented into three experimental conditions: pre-stimulation, during stimulation, and post-stimulation. Each condition was stored in separate EDF files.

All signals were notch filtered at 50, 100, and 150 Hz (Q=30) to remove line noise and harmonics. A fourth-order Butterworth low-pass filter with a 190 Hz cutoff was applied to reduce high-frequency artifacts. To identify stimulation events, a high-pass filter (cutoff = 100 Hz) was applied to the trigger channel, and events were detected based on threshold crossings. Stimulation onsets were detected from the trigger channel in the during-stimulation recording only. For stimulation-locked analyses, the during-stimulation data were epoched from −15 to +15 s relative to each detected burst onset. To ensure comparable recording durations across conditions, the pre- and post-stimulation data were trimmed to match the duration of the during-stimulation recording. Epochs falling outside the valid signal range were excluded. All valid epochs were downsampled from 2000 Hz to 500 Hz using polyphase decimation to reduce computational load while preserving signal integrity.

Channel selection for analysis was based on a predefined anatomical mapping of electrode contacts, tailored to each participant. Only contacts located in predefined regions of interest (ROIs) were retained for further analysis. After filtering and channel selection, condition-specific epoch data were saved as .mat files for downstream analysis using MATLAB (2024a) or Python-based toolchains.

##### 4.2.2.2 TNS evoked potential analysis

To quantify i200-TNS–evoked responses in stereo-EEG data, we computed group-level event-related potentials (ERPs) for each region of interest (ROI). Analyses were performed on previously preprocessed data, which were epoched from −15 to +15 seconds relative to stimulation onset (0 s) and sampled at 500 Hz. Signals were bandpass filtered (0.1–20 Hz, 2nd-order Butterworth) to isolate low-frequency components, and baseline correction was applied using the −200 to 0 ms pre-stimulus window. For each electrode channel, trial-averaged evoked potentials were computed, followed by subject-level averaging across channels within each ROI. Group-level ERPs were then generated by averaging across subjects. Peak and trough amplitudes and latencies were identified within a post-stimulus window of 0–1 s to capture early evoked components. To estimate variability, 95% confidence intervals (CI) were calculated across subjects. Group-level responses were visualized with shaded CI regions and overlaid with peak/trough markers. ROIs with fewer than three contributing electrodes across subjects were excluded from latency analysis. All computations were performed using custom MATLAB scripts.

##### 4.2.2.3 Human sEEG hippocampal phase amplitude coupling analysis and statistics

Hippocampal phase-amplitude coupling (PAC) was analysed from burst-centred sEEG epochs recorded from ipsilateral hippocampal contacts. For each i200-TNS burst, data were segmented from 15 s before to 15 s after burst onset, with burst onset defined as 0 s. Signals were analysed separately for the pre-stimulation, stimulation and post-stimulation recording epochs.

PAC was quantified using the phase of theta-band activity and the amplitude of gamma-band activity. Theta phase was extracted from the 4-8 Hz band, and gamma amplitude was estimated from 30-100 Hz in 5-Hz steps using 10-Hz-wide amplitude bands. Signals were filtered with zero-phase Butterworth filters, and analytic phase and amplitude were obtained using the Hilbert transform. For each trial, amplitude-frequency bin and 1-s sliding window, advanced in 250-ms steps, PAC was calculated as the absolute mean vector length of gamma amplitude weighted by theta phase, abs(mean(gamma_amp.* exp(1i * theta_phase))). PAC values were converted to PAC_z by normalizing each trial to its own pre-burst baseline windows. Channel-level PAC_z values were then averaged across trials, and subject-level values were obtained by averaging across hippocampal contacts within each subject.

The burst interval from 0 to 1 s was treated as invalid for PAC inference because stimulation artefact removal and band-pass filtering can distort phase and amplitude estimates during and immediately around stimulation. This interval was therefore masked in visualizations and was not used for the primary pre- versus post-burst comparison. The main after-effect analysis focused on the stimulation epoch and compared mean PAC_z in the pre-burst window (-15 to 0 s) with mean PAC_z in the post-burst window (1 to 15 s).

For group visualization, PAC_z heat maps were generated by averaging subject-level PAC_z values across subjects for each time window and gamma-amplitude frequency. The phase-amplitude profile was generated from the peak post-burst PAC_z tile by reloading the corresponding raw hippocampal data, binning theta phase from -π to π, and averaging gamma amplitude within each phase bin. Amplitude values were normalized within each gamma-frequency band for display. The representative single-trial example was selected from the subject-channel and post-burst tile with the highest PAC_z, and the trial with the highest single-trial PAC score was shown for illustration only.

Statistical inference for the primary after-effect was performed at the channel level using a linear mixed-effects model with time period as a fixed effect and random intercepts for subject and subject-channel: PAC_z ∼ period + (1|subject) + (1|subject_channel). Models were fitted in MATLAB using restricted maximum likelihood. Subject-level means and 95% confidence intervals were used for visualization, whereas statistical inference was based on the mixed-effects model to account for the nested structure of channels within subjects.

##### 4.2.2.4 Human sEEG hippocampal ripples analysis and statistics

Following preprocessing-stage exclusion of pathological high-frequency oscillations and artefacts, hippocampal ripple events were detected separately in pre-, during-, and post-stimulation sEEG recordings using a custom MATLAB pipeline. Signals were analysed at the downsampled rate of 500 Hz. Continuous 20-min sEEG recordings were segmented during preprocessing into 40 non-overlapping 30-s subepochs. During stimulation, these segments were aligned to the 40x i200-TNS bursts, whereas the pre- and post-stimulation recordings were divided into temporally matched segments. Because the resulting subepochs were stored as separate data segments, each was filtered and analysed independently to avoid boundary-related filtering artefacts and spurious ripple detections. Nonfinite samples were linearly interpolated when possible, using nearest-value padding at the recording boundaries.

For each valid subepoch, sEEG signals were bandpass-filtered at 80–120 Hz using a second-order zero-phase Butterworth filter. The filtered signal was squared and smoothed using a 10-ms moving-average kernel. Ripple-band power was then normalized to z scores using the mean and standard deviation pooled across valid subepochs within the same channel and time condition. Candidate events were defined as contiguous periods with normalized ripple power exceeding 2 SD. Candidate segments separated by less than 30 ms were merged. Events were retained only if they contained a normalized-power peak exceeding 3.5 SD and had a duration of 20–500 ms. Ripple duration was defined from the low-threshold onset to offset ^6^.Ripple peak time was defined as the maximal negative deflection in the ripple-band-filtered signal.

For each hippocampal channel and time condition, ripple rate was calculated as the total number of retained events divided by the total valid recording duration and expressed as events/min. Mean ripple duration was calculated across retained events and expressed in milliseconds. Channels without detected ripples contributed a zero rate but no duration value.

Primary inference used separate channel-level linear mixed-effects models for ripple rate and mean ripple duration. Models were fitted by restricted maximum likelihood in MATLAB, with time condition as a categorical fixed effect and subject-specific condition effects:

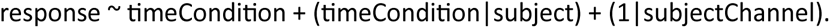

The model therefore included subject-specific random intercepts and time-condition slopes, together with a random intercept for each unique subject-channel combination. Marginal ANOVA tests assessed the overall effect of time condition. Planned contrasts compared During versus Pre, Post versus Pre, and During versus Post. Contrast P values were Bonferroni-corrected within each metric, and 95% confidence intervals were reported.

As complementary analyses, the script calculated paired Wilcoxon signed-rank tests on subject-level channel means, with paired-effect size reported as Cohen’s dz and Bonferroni-corrected P values. Sensitivity analyses included an animal-compatible random-intercept LME for both metrics, response ∼ epoch + (1|subjectID) + (1|ChannelID). For ripple rate, a further sensitivity analysis used a Poisson generalized linear mixed-effects model of subepoch ripple counts with a log-duration offset, subject-specific time-condition slopes, subject-channel and subject-channel-subepoch random intercepts, and an observation-level random effect. Pearson dispersion was calculated as a diagnostic for overdispersion.

##### 4.2.2.5 Human sEEG hippocampal ripples triggered cortical power analysis and statistics

To assess whether hippocampal ripples were associated with time-locked responses in cortical and medial temporal regions, we performed ripple-triggered spectral power analysis on the preprocessed human sEEG data. The analysis used the same pre/during/post session structure and preprocessing framework described above. Hippocampal ripple events were detected in contacts assigned to the ipsilateral hippocampal region (IpsHipChan), and cortical responses were evaluated in ipsilateral medial prefrontal cortex (mPFC), medial temporal cortex (MTC), and superior temporal cortex (STC) contacts (IpsmPFCChan, IpsMTCChan, and IpsSTCChan).

Ripple detection was performed independently for each hippocampal contact and analysis epoch. Signals were band-pass filtered between 80 and 120 Hz using a second-order zero-phase Butterworth filter. The squared filtered signal was smoothed with a 10-ms moving-average kernel and standardized using the mean and standard deviation calculated from all valid samples across epochs within the same channel and session phase. Candidate events were defined as contiguous periods exceeding 2 standard deviations, with gaps shorter than 30 ms merged. Events were retained when their peak amplitude exceeded 3.5 standard deviations and their duration was between 20 and 500 ms. Ripple peak time was defined from the maximum negative deflection in the filtered 80–120-Hz signal. Non-finite samples were linearly interpolated, with nearest-value padding at epoch boundaries.

For each cortical contact, spectral power was estimated separately in five predefined frequency bands: theta (4–8 Hz), alpha/sigma (8–13 Hz), beta (13–30 Hz), gamma (30–80 Hz), and ripple-band activity (80–120 Hz). The 8–13-Hz band was labelled alpha/sigma activity because the recordings were obtained during wakefulness and were not interpreted as sleep spindles. Each band was extracted using a second-order zero-phase Butterworth filter. The analytic amplitude was obtained with the Hilbert transform, converted to instantaneous power by squaring the amplitude, and smoothed with a 50-ms moving-average kernel. Baseline power was defined as the median power across all valid samples and epochs for the corresponding cortical contact, frequency band, and session phase.

For every detected hippocampal ripple, cortical power was extracted from −500 to +500 ms relative to the hippocampal ripple peak. Events for which this window extended beyond the available epoch were discarded. During stimulation sessions, ripple events whose peak occurred between −250 and +1,250 ms relative to the stimulation onset were excluded to avoid contamination by stimulation-related artifacts or immediate stimulation-evoked activity. Event-related power was expressed in decibels as:

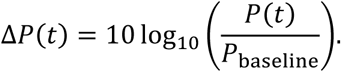

For each hippocampal–cortical contact pair, event-related waveforms were averaged across valid ripple events. The primary response measure was the mean cortical power change from 0 to 200 ms after the hippocampal ripple peak. Peak power and peak latency within this interval were also retained as descriptive measures. Each contact-pair and session-phase combination contributed one observation to the primary statistical analysis. Individual ripple events were not treated as independent observations. Contact pairs without valid ripple-triggered events for a given phase and frequency band were omitted from the corresponding model.

Statistical analyses were performed in MATLAB using linear mixed-effects models (fitlme). Separate models were fitted for each cortical ROI and frequency band. The primary contact-pair-level model was:

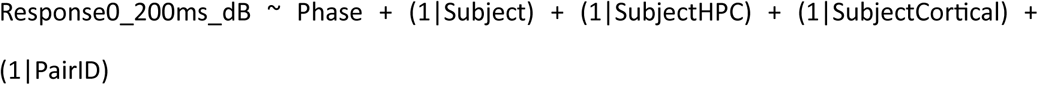

where Phase was a categorical factor with pre-stimulation as the reference level. SubjectHPC represented the subject-by-hippocampal-contact grouping factor, SubjectCortical represented the subject-by-cortical-contact grouping factor, and PairID represented the subject-by-hippocampal-contact-by-cortical-contact pair. These random intercepts accounted for repeated measurements across session phases and the hierarchical dependence of observations arising from subjects, contacts, and hippocampal–cortical pairs.

The overall effect of session phase was evaluated using the fixed-effect ANOVA from the fitted mixed-effects model. Planned two-sided linear contrasts tested during versus pre, post versus pre, and during versus post. Contrast estimates, standard errors, test statistics, model-derived degrees of freedom, and p values were obtained from the fitted model using the fixed-effect covariance matrix and coefTest. A model-derived standardized effect size was additionally calculated as 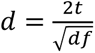, where *t* is the contrast test statistic and *df* is the corresponding model-derived denominator degrees of freedom. Bonferroni-adjusted p values across the three planned contrasts were reported within each ROI-by-frequency-band model.

As a sensitivity analysis, contact-pair responses were first summarized by taking the median across available contact pairs within each subject, phase, ROI, and frequency band. These subject-level summaries were analysed using a reduced model containing Phase and a subject-level random intercept:

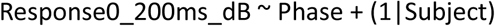

For visualization, ripple-triggered time courses were summarized at the subject level, whereas the 0–200-ms response plots displayed the fixed-effect marginal means and 95% confidence intervals estimated from the primary contact-pair-level model.

#### 4.2.3 Human memory behavior experiment and scalp EEG measurement

##### 4.2.3.1 Participant recruitment and experimental design

A total of 37 right-handed participants (aged 18–45 years) were recruited for this study, following a randomized crossover design. Each participant was scheduled for two experimental sessions: one with active trigeminal nerve stimulation (eTNS) and one with sham stimulation, separated by at least 2 days. In both sessions, two patch electrodes were placed on the skin over the mandibular branch of the trigeminal nerve. Stimulation intensity was individually determined before each session using 1-s, 200-Hz test bursts. Current was increased in 0.5 mA increments until participants reported a clearly perceptible but comfortable sensation without pain; This intensity was then maintained throughout the session. In this experiment two stimulation conditions were tested: 1) i200-TNS condition: This protocol was designed to match the stimulation paradigm used in the animal experiments. Trains of biphasic square pulses (200 Hz, 200 μs pulse width) were delivered in 1-second bursts, each followed by a 29-second inter-train interval. Stimulation was applied for a total of 20 minutes, comprising 40 trains. This duration was chosen to better align with common clinical neuromodulation protocols. 2) Sham condition: Electrodes were placed identically and stimulation duration same as another stimulation condition, but no electrical stimulation was delivered.

##### 4.2.3.2 The FACE-NAME-OCCUPATION task

The face–name-occupation memory task was designed to evaluate associative memory over short- and long-term intervals, comprising three phases: two learning trials, a short-term recall test, and a delayed long-term memory evaluation. As shown in Fig. 5b, twelve unfamiliar faces were consecutively displayed on the screen, each associated with a distinct name and occupation. Participants were directed to memorize the associations between the face and its corresponding name–occupation pair over two successive encoding trials. During each learning trial, participants were instructed to read aloud the name and occupation displayed beneath each face and to actively attempt memorization. Immediately following each learning trial, a reinforced recall phase was performed during which each face was reintroduced without labels. Participants were instructed to vocally recall their associated name (RN1/RN2) and occupation (RO1/RO2). Short-term memory (STM) was evaluated two minutes after the second learning trial. Participants were shown each face separately and instructed to recall and report its associated name (RSN) and occupation (RSO). Long-term memory (LTM) was tested 20 minutes after the short-term recall task and consisted of three components:1) Face Recognition (Q1): For each trial, participants were shown three faces and asked to identify the one previously learned. 2) Free Recall (Q2): Once the correct face was selected, it was displayed on the screen, and participants were asked to recall the associated name and occupation. 3) Cued Recognition (Q3): A previously learned face was presented alongside three name–occupation pair options, and participants were asked to select the pair that correctly matched the face.

The sequence of face presentations was randomized for each test. For all recall tasks (reinforced learning, short-term memory, and long-term memory), if participants were unable to remember either the name, the occupation, or both, they were instructed to respond with “I don’t know.”

The FACE-NAME-OCCUPATION task was implemented using Psychophysics Toolbox Version 3 (PTB-3) in MATLAB (version 2024a). The face pictures were selected from the Chicago FACE Database ^76^, comprising a total of twelve faces per set: six male and six female. Each set included individuals from four ethnic backgrounds—Asian (n = 1), Black (n = 1), Latinx (n = 1), and White (n = 3)—to ensure diversity and demographic balance. Names were drawn from the most common names over the past century, as reported by the U.S. Social Security Administration ^77^. Occupation labels were selected to represent a board spectrum of socioeconomic roles and were curated from publicly available lists on Wikipedia (https://en.wikipedia.org/wiki/Lists_of_occupations).

##### 4.2.3.3 Behavior evaluation and statistical analysis

Behavioral performance in the face–name memory task was assessed by quantifying associative recall accuracy across three task phases: (1) the encoding phase, based on the combined recall accuracy during the two reinforced learning trials (RN(1+2) for names; RO(1+2) for occupations); (2) the short-term memory phase, measured by recall accuracy 2mins following the encoding phase (RSN for names; RSO for occupations); and (3) the long-term memory phase, assessed 20 minutes after short-term memory phase (RLN for names; RLO for occupations).

To quantify the effects of stimulation on different memory stages, Recall accuracy gains were computed as follows: (1) Encoding enhancement: ΔEnco_Name = RSN − RN(1+2); ΔEnco_Occupation = RSO − RO(1+2). (2) Consolidation enhancement: ΔConso_Name = RLN − RSN; ΔConso_Occupation = RLO – RSO. These delta scores reflect changes in recall accuracy attributable to stimulation during either the encoding or consolidation phase.

Behavioral outcomes were analysed in the 36 participants who completed both sessions. Four prespecified within-participant comparisons were performed: encoding-related name and occupation recall gains and consolidation-related name and occupation recall gains. For each outcome, normality of the paired sham-minus-i200-TNS differences was assessed using the Shapiro–Wilk test. Normally distributed differences were analysed using two-sided paired *t*-tests; otherwise, two-sided Wilcoxon signed-rank tests were used. All four paired-difference distributions met the normality assumption, and paired *t*-tests were therefore used. *P* values were Bonferroni-adjusted by multiplication by four, with adjusted values capped at 1. Effect sizes were reported as Cohen’s *d_z_*, calculated as the mean paired difference divided by its standard deviation. Statistical significance was defined as an adjusted *P* < 0.05.

#### 4.2.4 EEG Experiment

##### 4.2.4.1 EEG recording

The experiment was conducted in a sound-attenuated and electrically shielded recording room. Participants were seated comfortably approximately 1.5 m from an LCD screen on which the face–name associative memory task was presented. Continuous electroencephalographic (EEG) activity was recorded using a 64-channel BioSemi ActiveTwo system (BioSemi, The Netherlands). Electrodes were positioned according to the international 10–20 system. Data were acquired at a sampling rate of 4 kHz using the default Common Mode Sense (CMS) and Driven Right Leg (DRL) electrodes as the online reference. Event markers indicating trial and stimulus onsets were generated by the stimulation computer and synchronized with the EEG data stream using a StimTracker device (Cedrus, USA).

##### 4.2.4.2 EEG data processing and analysis

###### General EEG preprocessing

EEG preprocessing was performed using custom MATLAB scripts in combination with functions from the FieldTrip toolbox ^78^. The same preprocessing framework was applied across analyses, with analysis-specific filtering, epoching, and artifact handling described below.

For each participant, continuous 64-channel EEG data were first downsampled from 4 kHz to 500 Hz to reduce computational load while preserving the frequency range of interest. Power-line noise was attenuated using a third-order Butterworth notch filter at 50 Hz and its harmonics. The data were then band-pass filtered between 0.1 and 140 Hz using a third-order Butterworth filter. Channels showing persistent noise, flat signals, or abnormal activity were identified by visual inspection and removed from subsequent preprocessing. Independent component analysis (ICA) was then performed using the *ft_componentanalysis* function (FieldTrip, method ‘runica’) to identify and remove components (*ft_rejectcomponent* function) associated with eye movements and other stereotyped non-neural artifacts. The cleaned continuous data were subsequently re-referenced to the common average reference.

After continuous preprocessing, data were segmented into epochs according to the event markers relevant to each analysis. A final epoch-level quality-control procedure was then applied. Within each epoch, channels with abnormal statistical properties, defined as kurtosis, mean, or variance exceeding three standard deviations from the across-channel distribution, were marked as noisy and repaired by spatial interpolation using the *ft_channelrepair* function (FieldTrip). Epochs were rejected if more than one-third of channels exceeded a peak amplitude of ±100 µV, indicating excessive residual noise. The resulting artifact-cleaned epochs were used for the evoked-potential, visual-evoked-potential, and resting-state spectral analyses described below.

###### Evoked potentials elicited by i200-TNS

Stimulation-related evoked potentials were analyzed in the i200-TNS group. Data from 34 participants were included. Two participants were excluded because of incomplete EEG markers during the consolidation stage. For each participant, EEG data from the 20-min consolidation period were extracted.

Because trigeminal nerve stimulation introduced short stimulation-locked artifacts, an additional artifact-correction step was applied before standard preprocessing. Power-line noise was first removed as described above, and the data were high-pass filtered at 0.1 Hz. Stimulation artifacts were detected using a threshold-based algorithm applied to the first-order temporal derivative of the EEG signal. For each detected artifact, a 2.5-ms segment centred on the artifact, corresponding to 10 samples at the original 4-kHz sampling rate, was replaced by linear interpolation using the *linspace* function (MATLAB). The artifact-corrected data were then low-pass filtered at 140 Hz and processed using the general preprocessing pipeline, including downsampling to 500 Hz, visual channel inspection, ICA-based artifact removal, and common average re-referencing.

For evoked-potential analysis, the preprocessed consolidation data were band-pass filtered between 0.2 and 30 Hz using a third-order Butterworth filter and segmented into epochs time-locked to the stimulation markers. This procedure yielded an expected 40 stimulation-locked epochs per participant. Epochs were subjected to the artifact-rejection procedure described above. Valid epochs were baseline-corrected using the 200-ms pre-stimulus interval. Individual evoked potentials were obtained by averaging all valid epochs within each participant. Grand-average evoked potentials were then computed across participants for visualization.

###### Visual-evoked potentials

Visual-evoked potentials elicited by face stimuli were analyzed to assess the effects of i200-TNS on memory-related neural responses. VEPs were examined during the short-term memory and long-term memory test stages and compared between the sham group and the i200-TNS group.

For each participant, preprocessed EEG data were band-pass filtered between 0.2 and 30 Hz using a third-order Butterworth filter. Data were then segmented into epochs time-locked to face-stimulus markers during the short-term and long-term memory test stages. Epochs were screened using the same artifact-rejection procedure described above. Valid epochs were baseline-corrected using the 200-ms pre-stimulus interval. Subject-level VEPs were computed separately for the short-term and long-term memory stages by averaging valid epochs within each stage. Grand-average VEPs were then calculated across participants within each group for visualization and statistical comparison.

###### Resting-stage power spectral density

Resting-state power spectral density was analyzed to assess changes in spontaneous neural activity across the three 2-min resting stages of the face–name associative memory task. Preprocessed EEG data were band-pass filtered between 0.2 and 90 Hz using a third-order Butterworth filter. Data corresponding to the three resting stages, denoted R1, R2, and R3, were extracted according to event markers and segmented into non-overlapping 10-s epochs. The general epoch-level artifact-rejection procedure was applied to all resting-state epochs.

Power spectral density was estimated for each EEG channel and epoch using the short-time Fourier transform implemented with the MATLAB *spectrogram* function. Power values were converted to decibel units. For visualization, grand-average spectra were computed across epochs and participants for each group and resting stage. The final analysis included 33 participants in the sham group and 33 participants in the i200-TNS group. To quantify stimulation-related changes in spectral power, PSD differences were calculated between R2 and R1 and between R3 and R2. These changes were evaluated in five frequency bands: delta, 0.5–4 Hz; theta, 4–8 Hz; alpha, 8–13 Hz; beta, 13–30 Hz; and gamma, 30–90 Hz.

##### 4.2.4.3 Statistical analysis

Statistical analyses were performed using FieldTrip and MATLAB. VEP differences between the short-term and long-term memory stages were assessed using cluster-based permutation tests implemented with *ft_timelockstatistics* (FieldTrip). Dependent-samples t-tests were used at the sample level. Clusters were formed using a cluster-forming threshold of *p* < 0.05, with a minimum of two neighbouring channels required to define a cluster. Cluster-level significance was assessed using the maximum-sum statistic with 1,000 random permutations. Two-tailed cluster-level tests were considered significant at *p* < 0.05.

Resting-state power changes between R2 and R1 and between R3 and R2 were analyzed using cluster-based permutation tests with the same statistical framework. For these analyses, power values were averaged within each frequency band of interest before statistical testing. In addition, mean power across seven left-frontal electrodes, Fp1, AF7, AF3, F7, F5, F3 and F1, was analyzed using linear mixed-effects models with resting stage as a fixed effect. Post-hoc comparisons were performed using coefTest with Bonferroni correction. The significance threshold was set to α = 0.05 for all statistical analyses.

## Data Availability

The data supporting the findings of this study are available from the corresponding authors upon reasonable request. De-identified human data, including intracranial sEEG recordings, may be subject to institutional ethics and privacy restrictions. Summary data supporting the main findings are provided in the manuscript and Supplementary Information.

## Acknowledgements

This work was supported by FWO Funding G0B4520N. We thank Dr. Yuhong Sun (University of Manchester) for constructive technical advice on the in vivo rodent fEPSP experiments.

## Contributions

**Liyi Chen:** Writing – original draft, Visualization of Results2.1-2.5, Validation of Results2.1-2.6, Software of Results2.1-2.4, Methodology of Results2.1-2.6, Investigation of Results2.1-2.6, Data curation of Results2.1-2.6, Conceptualization, Project administration. **Qiang Sun:** Writing – original draft, Visualization of Results 2.6, Validation of Results 2.6, Software of Results2.5-2.6. **Xinxia Guo:** Review & editing, Investigation Results2.3-2.4, Data curation Results2.3-2.4. **Hemmings Wu:** Review & editing, Methodology of Results2.3-2.4, Supervision of Results2.3-2.4, Funding acquisition of Results2.3-2.4. **Boateng Asamoah:** Methodology of Results2.1. **Wentai Ye:** Review & editing, Data curation Results2.3. **Nina Seminck:** Review & editing, Methodology of Results2.6. **Haorun Huang:** Review & editing, Data curation Results2.5. **Myles Mc Laughlin:** Writing – Review & editing, Visualization, Validation, Supervision, Project administration, Methodology, Funding acquisition, Data curation, Conceptualization.

