## Supplementary for "Temporal patterning of trigeminal nerve stimulation gates hippocampal plasticity across species"

### Supplementary Figures

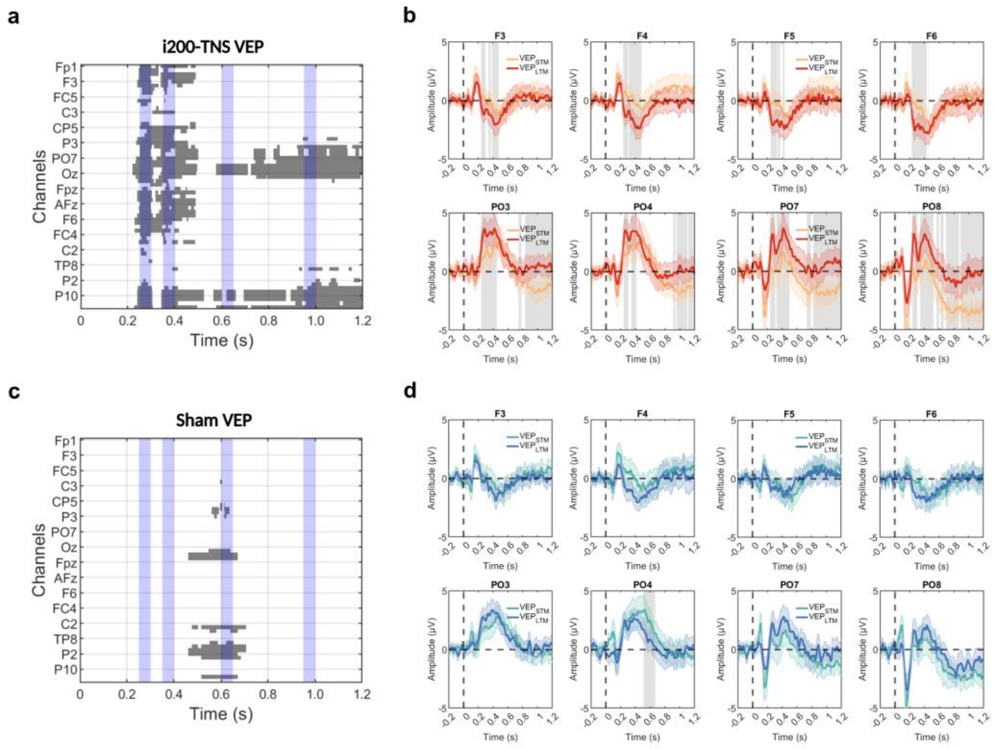

**Figure S1. Visual evoked potentials (VEPs) during the short- (STM) and long-term (LTM) memory test.**

**a, c** Permutation test results from the i200-TNS and Sham group, respectively. Channel-time clusters (in grey) showed significant difference between the VEP amplitude of STM and LTM. Parietal-occipital regions demonstrated longer lasting significant difference compared to frontal regions. Four representative time windows were selected for visualization in the main text. **b, d** Comparison between STM and LTM VEPs from frontal and parietal-occipital channels for the i200-TNS and Sham group, respectively. Shaded regions indicate there are significant differences

### Supplementary Tables

#### Table1.LME model and statistical details of i200-TNS on fEPSP slope (Fig.1c)

**Reference: Fig.1c**

**Linear mixed-effects model fit by ML**

| **Model information:** | | | **Model fit statistics:** | | | |
|---|---|---|---|---|---|---|
| Number of observations | 92 | | AIC | BIC | ogLikelihoo | Deviance |
| Fixed effects coefficients | 12 | | 710.23 | 745.54 | -341.11 | 682.23 |
| Random effects coefficients | 8 | | | | | |
| Covariance parameters | 2 | | | | | |

Formula:

**fEPSPslope~ 1 + timeCond + (1 | ratNum)**

**Fixed effects coefficients (95% CIs):**

| Name | Estimate | SE | tStat | DF | pValue | Lower | Upper |
|---|---|---|---|---|---|---|---|
| (Intercept) | 100 | 4.749644 | 21.05421 | 80 | 2.28E-34 | 90.54791 | 109.452092 |
| timeCond_Seg2 | 3.579142348 | 4.363669 | 0.820214 | 80 | 4.15E-01 | -5.10484 | 12.26312017 |
| timeCond_Seg3 | 7.852080663 | 4.363669 | 1.799422 | 80 | 7.57E-02 | -0.8319 | 16.53605849 |
| timeCond_Seg4 | 21.7224387 | 4.363669 | 4.978022 | 80 | 3.62E-06 | 13.03846 | 30.40641652 |
| timeCond_Seg5 | 20.09596604 | 4.363669 | 4.605291 | 80 | 1.53E-05 | 11.41199 | 28.77994386 |
| timeCond_Seg6 | 21.54770256 | 4.363669 | 4.937978 | 80 | 4.24E-06 | 12.86372 | 30.23168039 |
| timeCond_Seg7 | 23.35766349 | 4.363669 | 5.352758 | 80 | 8.07E-07 | 14.67369 | 32.04164131 |
| timeCond_Seg8 | 26.11272393 | 4.363669 | 5.984121 | 80 | 5.84E-08 | 17.42875 | 34.79670176 |
| timeCond_Seg9 | 32.97707146 | 4.363669 | 7.557189 | 80 | 5.93E-11 | 24.29309 | 41.66104928 |
| timeCond_Seg10 | 23.7834614 | 4.532274 | 5.247579 | 80 | 1.24E-06 | 14.76395 | 32.80297318 |
| timeCond_Seg11 | 22.65039288 | 4.532274 | 4.997579 | 80 | 3.35E-06 | 13.63088 | 31.66990466 |
| timeCond_Seg12 | 26.90837845 | 4.744427 | 5.671576 | 80 | 2.17E-07 | 17.46667 | 36.35008943 |

**Random effects covariance parameters (95% CIs):**

Group: ratNum (8 Levels)

| Name1 | Name2 | Type | Estimate | Lower | Upper |
|---|---|---|---|---|---|
| (Intercept) | (Intercept) | std' | 10.213 | 6.0644 | 17.2 |

Group: Error

| Name | Estimate | Lower | Upper |
|---|---|---|---|
| 'Res Std' | 8.7273 | 7.5026 | 10.152 |

**ANOVA marginal tests: DFMethod = 'Residual'**

| Term | FStat | DF1 | DF2 | pValue |
|---|---|---|---|---|
| (Intercept) | 443.2798142 | 1 | 80 | 2.28E-34 |
| timeCond | 10.29876086 | 11 | 80 | 1.93E-11 |

Linear contrasts (Segment 1 vs Seg2–12, Bonferroni corrected):

| Segment | Time period(minute) | Estimate | SE | t_value | df | p_value | p_Bonferroni | F_value | df1 | Cohen_d |
|---|---|---|---|---|---|---|---|---|---|---|
| 1 | [-30 -20] | / | / | / | / | / | / | / | / | / |
| 2 | [-20 -10] | 3.579142 | 4.363669 | 0.820214 | 80 | 0.414531 | 1 | 0.672751 | 1 | 0.183405 |
| 3 | [-10 0] | 7.852081 | 4.363669 | 1.799422 | 80 | 0.075723 | 0.832948409 | 3.237919 | 1 | 0.402363 |
| 4 | [ 5 15] | 21.72244 | 4.363669 | 4.978022 | 80 | 3.62E-06 | 3.98251E-05 | 24.7807 | 1 | 1.11312 |
| 5 | [ 15 25] | 20.09597 | 4.363669 | 4.605291 | 80 | 1.53E-05 | 0.00016848 | 21.20871 | 1 | 1.029774 |
| 6 | [ 25 35] | 21.5477 | 4.363669 | 4.937978 | 80 | 4.24E-06 | 4.66187E-05 | 24.38363 | 1 | 1.104166 |
| 7 | [ 35 45] | 23.35766 | 4.363669 | 5.352758 | 80 | 8.07E-07 | 8.87635E-06 | 28.65202 | 1 | 1.196913 |
| 8 | [ 45 55] | 26.11272 | 4.363669 | 5.984121 | 80 | 5.84E-08 | 6.42642E-07 | 35.80971 | 1 | 1.33809 |
| 9 | [ 55 65] | 32.97707 | 4.363669 | 7.557189 | 80 | 5.93E-11 | 6.51879E-10 | 57.11111 | 1 | 1.689839 |
| 10 | [ 65 75] | 23.78346 | 4.532274 | 5.247579 | 80 | 1.24E-06 | 1.35918E-05 | 27.53708 | 1 | 1.173394 |
| 11 | [ 75 85] | 22.65039 | 4.532274 | 4.997579 | 80 | 3.35E-06 | 3.68687E-05 | 24.97579 | 1 | 1.117493 |
| 12 | [ 85 95] | 26.90838 | 4.744427 | 5.671576 | 80 | 2.17E-07 | 2.38986E-06 | 32.16677 | 1 | 1.268203 |

#### Table2.LME model and statistical details of i200-TNS on fEPSP slope (Fig.1d)

**Reference: Fig.1d**

**Linear mixed-effects model fit by ML**

| **Model information:** | | | **Model fit statistics:** | | | |
|---|---|---|---|---|---|---|
| Number of observations | 953 | | AIC | BIC | LogLikelihood | Deviance |
| Fixed effects coefficients | 2 | | 9006.6 | 9026.1 | -4499.3 | 8998.6 |
| Random effects coefficients | 8 | | | | | |
| Covariance parameters | 2 | | | | | |

Formula:

**fEPSPslope~ 1 + timeCond + (1 | ratNum)**

**Fixed effects coefficients (95% CIs):**

| Name | Estimate | SE | tStat | DF | pValue | Lower | Upper |
|---|---|---|---|---|---|---|---|
| (Intercept) | 103.854 | 2.823964 | 36.77596 | 951 | 6.92E-185 | 98.31208 | 109.3959 |
| timeCond_Post30 | 17.2255 | 1.744015 | 9.876924 | 951 | 5.73E-22 | 13.80294 | 20.64806 |

**Random effects covariance parameters (95% CIs):**

Group: ratNum (8 Levels)

| Name1 | Name2 | Type | Estimate | Lower | Upper |
|---|---|---|---|---|---|
| (Intercept) | (Intercept) | std' | 7.1864 | 4.1557 | 12.427 |

Group: Error

| Name | Estimate | Lower | Upper |
|---|---|---|---|
| 'Res Std' | 26.919 | 25.732 | 28.161 |

**ANOVA marginal tests: DFMethod = 'Residual'**

| Term | FStat | DF1 | DF2 | pValue |
|---|---|---|---|---|
| (Intercept) | 1352.471 | 1 | 951 | 6.92E-185 |
| timeCond | 97.55362 | 1 | 951 | 5.73E-22 |

| Comparison | Estimate | SE | t_value | df | p_value | F_value | df1 | Cohen_d |
|---|---|---|---|---|---|---|---|---|
| Post30_vs_Baseline | 17.2255 | 1.744015 | 9.876924 | 951 | 5.72875E-22 | 97.55362 | 1 | 0.640562 |

### Table3.LME model and statistical details of c100-TNS on fEPSP slope (Fig.1e)

**Reference: Fig.1e**

**Linear mixed-effects model fit by ML**

| **Model information:** | | | **Model fit statistics:** | | | |
|---|---|---|---|---|---|---|
| Number of observations | 96 | | AIC | BIC | ogLikelihoo | Deviance |
| Fixed effects coefficients | 12 | | 752.53 | 788.43 | -362.26 | 724.53 |
| Random effects coefficients | 8 | | | | | |
| Covariance parameters | 2 | | | | | |

Formula:

**fEPSPslope~ 1 + timeCond + (1 | ratNum)**

**Fixed effects coefficients (95% CIs):**

| Name | Estimate | SE | tStat | DF | pValue | Lower | Upper |
|---|---|---|---|---|---|---|---|
| (Intercept) | 100 | 4.064536 | 24.60306 | 84 | 3.74E-40 | 91.91723 | 108.0827747 |
| timeCond_Seg2 | -1.77002 | 4.907291 | -0.36069 | 84 | 7.19E-01 | -11.52871 | 7.988665217 |
| timeCond_Seg3 | 0.247519 | 4.907291 | 0.050439 | 84 | 9.60E-01 | -9.511168 | 10.0062064 |
| timeCond_Seg4 | 1.384528 | 4.907291 | 0.282137 | 84 | 0.778532 | -8.374159 | 11.14321522 |
| timeCond_Seg5 | 1.197641 | 4.907291 | 0.244053 | 84 | 0.807785 | -8.561047 | 10.95632767 |
| timeCond_Seg6 | 0.026814 | 4.907291 | 0.005464 | 84 | 0.995653 | -9.731873 | 9.785501475 |
| timeCond_Seg7 | -2.45783 | 4.907291 | -0.50085 | 84 | 0.617784 | -12.21652 | 7.300853506 |
| timeCond_Seg8 | 1.07242 | 4.907291 | 0.218536 | 84 | 0.827542 | -8.686267 | 10.83110729 |
| timeCond_Seg9 | -3.57269 | 4.907291 | -0.72804 | 84 | 0.468615 | -13.33138 | 6.185995691 |
| timeCond_Seg10 | 2.131613 | 4.907291 | 0.434377 | 84 | 0.665129 | -7.627075 | 11.89029965 |
| timeCond_Seg11 | 2.708846 | 4.907291 | 0.552004 | 84 | 0.582411 | -7.049841 | 12.46753351 |
| timeCond_Seg12 | -10.2008 | 4.907291 | -2.07871 | 84 | 0.040695 | -19.95953 | -0.442152877 |

**Random effects covariance parameters (95% CIs):**

Group: ratNum (8 Levels)

| Name1 | Name2 | Type | Estimate | Lower | Upper |
|---|---|---|---|---|---|
| (Intercept) | (Intercept) | std' | 5.9864 | 3.2833 | 10.915 |

Group: Error

| Name | Estimate | Lower | Upper |
|---|---|---|---|
| 'Res Std' | 9.8146 | 8.4666 | 11.377 |

**ANOVA marginal tests: DFMethod = 'Residual'**

| Term | FStat | DF1 | DF2 | pValue |
|---|---|---|---|---|
| (Intercept) | 605.3104 | 1 | 84 | 3.74E-40 |
| timeCond | 1.021815 | 11 | 84 | 4.35E-01 |

Linear contrasts (Segment 1 vs Seg2–12, Bonferroni corrected):

| Segment | period(mir | Estimate | SE | t_value | df | p_value | p_Bonferroni | F_value | df1 | Cohen_d |
|---|---|---|---|---|---|---|---|---|---|---|
| 1 | [-30 -20] | / | / | / | / | / | / | / | / | / |
| 2 | [-20 -10] | -1.77002 | 4.907291 | -0.36069223 | 84 | 0.719235 | 1 | 0.130099 | 1 | -0.07871 |
| 3 | [-10 0] | 0.247519 | 4.907291 | 0.050439091 | 84 | 0.959892 | 1 | 0.002544 | 1 | 0.011007 |
| 4 | [ 5 15] | 1.384528 | 4.907291 | 0.28213693 | 84 | 0.778532 | 1 | 0.079601 | 1 | 0.061567 |
| 5 | [ 15 25] | 1.197641 | 4.907291 | 0.244053285 | 84 | 0.807785 | 1 | 0.059562 | 1 | 0.053257 |
| 6 | [ 25 35] | 0.026814 | 4.907291 | 0.005464193 | 84 | 0.995653 | 1 | 2.99E-05 | 1 | 0.001192 |
| 7 | [ 35 45] | -2.45783 | 4.907291 | -0.5008534 | 84 | 0.617784 | 1 | 0.250854 | 1 | -0.1093 |
| 8 | [ 45 55] | 1.07242 | 4.907291 | 0.218536075 | 84 | 0.827542 | 1 | 0.047758 | 1 | 0.047688 |
| 9 | [ 55 65] | -3.57269 | 4.907291 | -0.72803735 | 84 | 0.468615 | 1 | 0.530038 | 1 | -0.15887 |
| 10 | [ 65 75] | 2.131613 | 4.907291 | 0.434376603 | 84 | 0.665129 | 1 | 0.188683 | 1 | 0.094789 |
| 11 | [ 75 85] | 2.708846 | 4.907291 | 0.552004397 | 84 | 0.582411 | 1 | 0.304709 | 1 | 0.120457 |
| 12 | [ 85 95] | -10.2008 | 4.907291 | -2.07871087 | 84 | 0.040695 | 0.44764538 | 4.321039 | 1 | -0.45361 |

#### Table4.LME model and statistical details of c100-TNS on fEPSP slope (Fig.1f)

**Reference: Fig.1f**

**Linear mixed-effects model fit by ML**

| **Model information:** | | **Model fit statistics:** | | | |
|---|---|---|---|---|---|
| Number of observations | 941 | AIC | BIC | ogLikelihoo | Deviance |
| Fixed effects coefficients | 2 | 9263.9 | 9283.3 | -4628 | 9255.9 |
| Random effects coefficients | 8 | | | | |
| Covariance parameters | 2 | | | | |

Formula:

**fEPSPslope~ 1 + timeCond + (1 | ratNum)**

**Fixed effects coefficients (95% CIs):**

| Name | Estimate | SE | tStat | DF | pValue | Lower | Upper |
|---|---|---|---|---|---|---|---|
| (Intercept) | 99.54419 | 1.526257 | 65.22111 | 939 | 0.00E+00 | 96.54892 | 102.5395 |
| timeCond_Post30 | 1.418442 | 2.157308 | 0.657506 | 939 | 5.11E-01 | -2.81526 | 5.652145 |

**Random effects covariance parameters (95% CIs):**

Group: ratNum (8 Levels)

| Name1 | Name2 | Type | Estimate | Lower | Upper |
|---|---|---|---|---|---|
| (Intercept) | (Intercept) | std' | 0 | NaN | NaN |

Group: Error

| Name | Estimate | Lower | Upper |
|---|---|---|---|
| 'Res Std' | 33.088 | 31.627 | 34.618 |

**ANOVA marginal tests: DFMethod = 'Residual'**

| Term | FStat | DF1 | DF2 | pValue |
|---|---|---|---|---|
| (Intercept) | 4253.793 | 1 | 939 | 0.00E+00 |
| timeCond | 0.432314 | 1 | 939 | 5.11E-01 |

| Comparison | Estimate | SE | t_value | df | p_value | F_value | df1 | Cohen_d |
|---|---|---|---|---|---|---|---|---|
| Post30_vs_Baseline | 1.418442 | 2.157308 | 0.657506 | 939 | 0.511017 | 0.432314 | 1 | 0.042914 |

### Table5.LME model and statistical details of i200-TNS on spike rate (Fig.1h)

**Reference: Fig.1h**

**Linear mixed-effects model fit by ML**

| **Model information:** | | | **Model fit statistics:** | | | |
|---|---|---|---|---|---|---|
| Number of observations | 2269 | | AIC | BIC | LogLikelihood | Deviance |
| Fixed effects coefficients | 12 | | 13509 | 13595 | -6739.7 | 13479 |
| Random effects coefficients | 206 | | | | | |
| Covariance parameters | 3 | | | | | |

Formula:

**SpikeRateMean ~ 1 + timeCond + (1 | ratNum) + (1 | unitID)**

**Fixed effects coefficients (95% CIs):**

| Name | Estimate | SE | tStat | DF | pValue | Lower | Upper |
|---|---|---|---|---|---|---|---|
| (Intercept) | 5.406927 | 0.816858 | 6.619173 | 2257 | 4.50E-11 | 3.805055 | 7.008799278 |
| timeCond_Seg2 | 0.189015 | 0.408689 | 0.462492 | 2257 | 6.44E-01 | -0.61243 | 0.990459774 |
| timeCond_Seg3 | 0.3057 | 0.408689 | 0.748003 | 2257 | 4.55E-01 | -0.49574 | 1.107144959 |
| timeCond_Seg4 | 2.022552 | 0.408689 | 4.948884 | 2257 | 8.01546E-07 | 1.221108 | 2.823996811 |
| timeCond_Seg5 | 2.975992 | 0.408689 | 7.281808 | 2257 | 4.52756E-13 | 2.174547 | 3.777436205 |
| timeCond_Seg6 | 3.991173 | 0.408689 | 9.765807 | 2257 | 4.29327E-22 | 3.189729 | 4.792618023 |
| timeCond_Seg7 | 2.790658 | 0.408689 | 6.828325 | 2257 | 1.10056E-11 | 1.989214 | 3.592102871 |
| timeCond_Seg8 | 3.599764 | 0.408689 | 8.808087 | 2257 | 2.48428E-18 | 2.79832 | 4.401208932 |
| timeCond_Seg9 | 4.493965 | 0.408689 | 10.99606 | 2257 | 1.9571E-27 | 3.69252 | 5.295409269 |
| timeCond_Seg10 | 4.252432 | 0.423322 | 10.04538 | 2257 | 2.94242E-23 | 3.42229 | 5.082573699 |
| timeCond_Seg11 | 4.507238 | 0.423322 | 10.64729 | 2257 | 7.29379E-26 | 3.677096 | 5.337379414 |
| timeCond_Seg12 | 4.714338 | 0.455814 | 10.34268 | 2257 | 1.57952E-24 | 3.82048 | 5.608196048 |

**Random effects covariance parameters (95% CIs):**

Group: ratNum (8 Levels)

| Name1 | Name2 | Type | Estimate | Lower | Upper |
|---|---|---|---|---|---|
| (Intercept) | (Intercept) | std' | 1.7217 | 0.71538 | 4.1435 |

Group: unitID (198 Levels)

| Name1 | Name2 | Type | Estimate | Lower | Upper |
|---|---|---|---|---|---|
| (Intercept) | (Intercept) | std' | 6.3667 | 5.7336 | 7.0697 |

Group: Error

| Name | Estimate | Lower | Upper |
|---|---|---|---|
| 'Res Std' | 4.0664 | 3.9444 | 4.1921 |

**ANOVA marginal tests: DFMethod = 'Residual'**

| Term | FStat | DF1 | DF2 | pValue |
|---|---|---|---|---|
| (Intercept) | 43.81345 | 1 | 2257 | 4.50E-11 |
| timeCond | 35.9986 | 11 | 2257 | 1.23E-71 |

Linear contrasts (Segment 1 vs Seg2–12, Bonferroni corrected):

| Segment | period(mi | Estimate | SE | t_value | df | p_value | p_Bonferroni | F_value | df1 | Cohen_d |
|---|---|---|---|---|---|---|---|---|---|---|
| 1 | [-30 -20] | / | / | / | / | / | / | / | / | / |
| 2 | [-20 -10] | 0.189015 | 0.408689 | 0.462492 | 2257 | 0.643773 | 1 | 0.213899 | 1 | 0.01947 |
| 3 | [-10 0] | 0.3057 | 0.408689 | 0.748003 | 2257 | 0.454536 | 1 | 0.559509 | 1 | 0.03149 |
| 4 | [ 5 15] | 2.022552 | 0.408689 | 4.948884 | 2257 | 8.02E-07 | 8.81701E-06 | 24.49145 | 1 | 0.208339 |
| 5 | [ 15 25] | 2.975992 | 0.408689 | 7.281808 | 2257 | 4.53E-13 | 4.98031E-12 | 53.02473 | 1 | 0.306552 |
| 6 | [ 25 35] | 3.991173 | 0.408689 | 9.765807 | 2257 | 4.29E-22 | 4.7226E-21 | 95.37098 | 1 | 0.411124 |
| 7 | [ 35 45] | 2.790658 | 0.408689 | 6.828325 | 2257 | 1.1E-11 | 1.21062E-10 | 46.62602 | 1 | 0.287461 |
| 8 | [ 45 55] | 3.599764 | 0.408689 | 8.808087 | 2257 | 2.48E-18 | 2.73271E-17 | 77.5824 | 1 | 0.370805 |
| 9 | [ 55 65] | 4.493965 | 0.408689 | 10.99606 | 2257 | 1.96E-27 | 2.15281E-26 | 120.9134 | 1 | 0.462915 |
| 10 | [ 65 75] | 4.252432 | 0.423322 | 10.04538 | 2257 | 2.94E-23 | 3.23666E-22 | 100.9096 | 1 | 0.422893 |
| 11 | [ 75 85] | 4.507238 | 0.423322 | 10.64729 | 2257 | 7.29E-26 | 8.02317E-25 | 113.3649 | 1 | 0.448233 |
| 12 | [ 85 95] | 4.714338 | 0.455814 | 10.34268 | 2257 | 1.58E-24 | 1.73747E-23 | 106.9711 | 1 | 0.435409 |

### Table6.LME model and statistical details of c100-TNS on spike rate (Fig.1i)

**Reference: Fig.1i**

**Linear mixed-effects model fit by ML**

| **Model information:** | | | **Model fit statistics:** | | | |
|---|---|---|---|---|---|---|
| Number of observations | 2520 | | AIC | BIC | LogLikelihood | Deviance |
| Fixed effects coefficients | 12 | | 15982 | 16069 | -7976 | 15952 |
| Random effects coefficients | 218 | | | | | |
| Covariance parameters | 3 | | | | | |

Formula:

**SpikeRateMean ~ 1 + timeCond + (1 | ratNum) + (1 | unitID)**

**Fixed effects coefficients (95% CIs):**

| Name | Estimate | SE | tStat | DF | pValue | Lower | Upper |
|---|---|---|---|---|---|---|---|
| (Intercept) | 8.067234259 | 0.944147 | 8.544465 | 2508 | 2.21E-17 | 6.215846 | 9.91862261 |
| timeCond_Seg2 | -1.327822222 | 0.489387 | -2.71324 | 2508 | 6.71E-03 | -2.28747 | -0.368178222 |
| timeCond_Seg3 | -0.435911111 | 0.489387 | -0.89073 | 2508 | 3.73E-01 | -1.39556 | 0.523732889 |
| timeCond_Seg4 | 2.959171429 | 0.489387 | 6.04669 | 2508 | 1.69922E-09 | 1.999527 | 3.918815428 |
| timeCond_Seg5 | 4.105915873 | 0.489387 | 8.389916 | 2508 | 8.00965E-17 | 3.146272 | 5.065559873 |
| timeCond_Seg6 | 3.616230159 | 0.489387 | 7.389306 | 2508 | 1.99959E-13 | 2.656586 | 4.575874159 |
| timeCond_Seg7 | 2.951877778 | 0.489387 | 6.031786 | 2508 | 1.86116E-09 | 1.992234 | 3.911521778 |
| timeCond_Seg8 | 2.163393651 | 0.489387 | 4.420619 | 2508 | 1.02607E-05 | 1.20375 | 3.123037651 |
| timeCond_Seg9 | 0.809704762 | 0.489387 | 1.654529 | 2508 | 0.098145291 | -0.14994 | 1.769348762 |
| timeCond_Seg10 | -2.054271429 | 0.489387 | -4.19764 | 2508 | 2.79102E-05 | -3.01392 | -1.094627429 |
| timeCond_Seg11 | -1.290993651 | 0.489387 | -2.63798 | 2508 | 0.008391661 | -2.25064 | -0.331349651 |
| timeCond_Seg12 | -0.491436508 | 0.489387 | -1.00419 | 2508 | 0.315384914 | -1.45108 | 0.468207492 |

**Random effects covariance parameters (95% CIs):**

Group: ratNum (8 Levels)

| Name1 | Name2 | Type | Estimate | Lower | Upper |
|---|---|---|---|---|---|
| (Intercept) | (Intercept) | std' | 2.0581 | 1.032 | 4.1044 |

Group: unitID (210 Levels)

| Name1 | Name2 | Type | Estimate | Lower | Upper |
|---|---|---|---|---|---|
| (Intercept) | (Intercept) | std' | 6.8987 | 6.232 | 7.6367 |

Group: Error

| Name | Estimate | Lower | Upper |
|---|---|---|---|
| 'Res Std' | 5.0147 | 4.8722 | 5.1614 |

**ANOVA marginal tests: DFMethod = 'Residual'**

| Term | FStat | DF1 | DF2 | pValue |
|---|---|---|---|---|
| (Intercept) | 73.00788126 | 1 | 2508 | 2.21E-17 |
| timeCond | 38.51142544 | 11 | 2508 | 2.88E-77 |

Linear contrasts (Segment 1 vs Seg2–12, Bonferroni corrected):

| Segment | Time period(minute) | Estimate | SE | t_value | df | p_value | p_Bonferroni | F_value | df1 | Cohen_d |
|---|---|---|---|---|---|---|---|---|---|---|
| 1 | [-30 -20] | / | / | / | / | / | / | / | / | / |
| 2 | [-20 -10] | -1.32782 | 0.489387 | -2.71324 | 2508 | 0.006709 | 0.073793676 | 7.361648 | 1 | -0.10836 |
| 3 | [-10 0] | -0.43591 | 0.489387 | -0.89073 | 2508 | 0.37316 | 1 | 0.793398 | 1 | -0.03557 |
| 4 | [ 5 15] | 2.959171 | 0.489387 | 6.04669 | 2508 | 1.7E-09 | 1.86914E-08 | 36.56246 | 1 | 0.241482 |
| 5 | [ 15 25] | 4.105916 | 0.489387 | 8.389916 | 2508 | 8.01E-17 | 8.81061E-16 | 70.39069 | 1 | 0.335061 |
| 6 | [ 25 35] | 3.61623 | 0.489387 | 7.389306 | 2508 | 2E-13 | 2.19954E-12 | 54.60184 | 1 | 0.2951 |
| 7 | [ 35 45] | 2.951878 | 0.489387 | 6.031786 | 2508 | 1.86E-09 | 2.04727E-08 | 36.38245 | 1 | 0.240886 |
| 8 | [ 45 55] | 2.163394 | 0.489387 | 4.420619 | 2508 | 1.03E-05 | 0.000112868 | 19.54188 | 1 | 0.176543 |
| 9 | [ 55 65] | 0.809705 | 0.489387 | 1.654529 | 2508 | 0.098145 | 1 | 2.737465 | 1 | 0.066076 |
| 10 | [ 65 75] | -2.05427 | 0.489387 | -4.19764 | 2508 | 2.79E-05 | 0.000307012 | 17.6202 | 1 | -0.16764 |
| 11 | [ 75 85] | -1.29099 | 0.489387 | -2.63798 | 2508 | 0.008392 | 0.092308276 | 6.958944 | 1 | -0.10535 |
| 12 | [ 85 95] | -0.49144 | 0.489387 | -1.00419 | 2508 | 0.315385 | 1 | 1.008393 | 1 | -0.0401 |

### Table7.LME model and statistical details of i200-TNS on fEPSP slope with clonidine blockade (Fig.2c)

**Reference: Fig.2c**

**Linear mixed-effects model fit by ML**

| **Model information:** | | | **Model fit statistics:** | | | |
|---|---|---|---|---|---|---|
| Number of observations | 60 | | AIC | BIC | LogLikelihood | Deviance |
| Fixed effects coefficients | 15 | | 462.7 | 498.3 | -214.35 | 428.7 |
| Random effects coefficients | 4 | | | | | |
| Covariance parameters | 2 | | | | | |

Formula:

**fEPSPslope~ 1 + timeCond + (1 | ratNum)**

**Fixed effects coefficients (95% CIs):**

| Name | Estimate | SE | tStat | DF | pValue | Lower | Upper |
|---|---|---|---|---|---|---|---|
| (Intercept) | 100 | 7.0274889 | 14.229834 | 45 | 2.86E-18 | 85.845911 | 114.1540892 |
| timeCond_Seg2 | 0.185226107 | 5.4023581 | 0.0342862 | 45 | 9.73E-01 | -10.69568 | 11.0661338 |
| timeCond_Seg3 | -3.354761144 | 5.4023581 | -0.620981 | 45 | 5.38E-01 | -14.23567 | 7.526146547 |
| timeCond_Seg4 | -16.13191308 | 5.4023581 | -2.986087 | 45 | 0.004559181 | -27.01282 | -5.251005392 |
| timeCond_Seg5 | -31.53752763 | 5.4023581 | -5.837734 | 45 | 5.43403E-07 | -42.41844 | -20.65661994 |
| timeCond_Seg6 | -33.47187205 | 5.4023581 | -6.195789 | 45 | 1.59808E-07 | -44.35278 | -22.59096436 |
| timeCond_Seg7 | -37.25945876 | 5.4023581 | -6.896888 | 45 | 1.44927E-08 | -48.14037 | -26.37855107 |
| timeCond_Seg8 | -33.60394426 | 5.4023581 | -6.220236 | 45 | 1.46977E-07 | -44.48485 | -22.72303657 |
| timeCond_Seg9 | -33.13141714 | 5.4023581 | -6.13277 | 45 | 1.98279E-07 | -44.01232 | -22.25050945 |
| timeCond_Seg10 | -33.32825412 | 5.4023581 | -6.169205 | 45 | 1.75033E-07 | -44.20916 | -22.44734643 |
| timeCond_Seg11 | -32.00421547 | 5.4023581 | -5.92412 | 45 | 4.04647E-07 | -42.88512 | -21.12330778 |
| timeCond_Seg12 | -30.84218688 | 5.4023581 | -5.709023 | 45 | 8.42529E-07 | -41.72309 | -19.96127919 |
| timeCond_Seg13 | -37.26582588 | 5.4023581 | -6.898067 | 45 | 1.44345E-08 | -48.14673 | -26.38491819 |
| timeCond_Seg14 | -29.45248148 | 5.4023581 | -5.451783 | 45 | 2.01724E-06 | -40.33339 | -18.57157379 |
| timeCond_Seg15 | -34.8632691 | 5.4023581 | -6.453343 | 45 | 6.61458E-08 | -45.74418 | -23.98236141 |

**Random effects covariance parameters (95% CIs):**

Group: ratNum (4 Levels)

| Name1 | Name2 | Type | Estimate | Lower | Upper |
|---|---|---|---|---|---|
| (Intercept) | (Intercept) | std' | 11.797 | 5.7864 | 24.052 |

Group: Error

| Name | Estimate | Lower | Upper |
|---|---|---|---|
| 'Res Std' | 7.6401 | 6.3484 | 9.1945 |

**ANOVA marginal tests: DFMethod = 'Residual'**

| Term | FStat | DF1 | DF2 | pValue |
|---|---|---|---|---|
| (Intercept) | 202.4881732 | 1 | 45 | 2.86E-18 |
| timeCond | 12.84083269 | 14 | 45 | 2.27E-11 |

Linear contrasts (Segment 1 vs Seg2–15, Bonferroni corrected):

| Segment | Time period(minute) | Estimate | SE | t_value | df | p_value | p_Bonferroni | F_value | df1 | Cohen_d |
|---|---|---|---|---|---|---|---|---|---|---|
| 1 | [-60 -50] | / | / | / | / | / | / | / | / | / |
| 2 | [-50 -40] | 0.1852261 | 5.4023581 | 0.0342862 | 45 | 0.9728006 | 1 | 0.0011755 | 1 | 0.0102222 |
| 3 | [-40 -30] | -3.354761 | 5.4023581 | -0.620981 | 45 | 0.5377451 | 1 | 0.3856173 | 1 | -0.185141 |
| 4 | [-30 -20] | -16.13191 | 5.4023581 | -2.986087 | 45 | 0.0045592 | 0.06382853 | 8.9167177 | 1 | -0.890279 |
| 5 | [-20 -10] | -31.53753 | 5.4023581 | -5.837734 | 45 | 5.434E-07 | 7.60764E-06 | 34.079135 | 1 | -1.740476 |
| 6 | [-10 0] | -33.47187 | 5.4023581 | -6.195789 | 45 | 1.598E-07 | 2.23731E-06 | 38.387804 | 1 | -1.847227 |
| 7 | [ 5 15] | -37.25946 | 5.4023581 | -6.896888 | 45 | 1.449E-08 | 2.02898E-07 | 47.567065 | 1 | -2.056255 |
| 8 | [ 15 25] | -33.60394 | 5.4023581 | -6.220236 | 45 | 1.47E-07 | 2.05767E-06 | 38.691341 | 1 | -1.854516 |
| 9 | [ 25 35] | -33.13142 | 5.4023581 | -6.13277 | 45 | 1.983E-07 | 2.7759E-06 | 37.610863 | 1 | -1.828439 |
| 10 | [ 35 45] | -33.32825 | 5.4023581 | -6.169205 | 45 | 1.75E-07 | 2.45046E-06 | 38.05909 | 1 | -1.839302 |
| 11 | [ 45 55] | -32.00422 | 5.4023581 | -5.92412 | 45 | 4.046E-07 | 5.66506E-06 | 35.095193 | 1 | -1.766231 |
| 12 | [ 55 65] | -30.84219 | 5.4023581 | -5.709023 | 45 | 8.425E-07 | 1.17954E-05 | 32.592945 | 1 | -1.702102 |
| 13 | [ 65 75] | -37.26583 | 5.4023581 | -6.898067 | 45 | 1.443E-08 | 2.02082E-07 | 47.583323 | 1 | -2.056606 |
| 14 | [ 75 85] | -29.45248 | 5.4023581 | -5.451783 | 45 | 2.017E-06 | 2.82413E-05 | 29.721933 | 1 | -1.625408 |
| 15 | [ 85 95] | -34.86327 | 5.4023581 | -6.453343 | 45 | 6.615E-08 | 9.26042E-07 | 41.645635 | 1 | -1.924015 |

### Table8.LME model and statistical details of i200-TNS on fEPSP slope with SCH-23390 blockade (Fig.2d)

**Reference: Fig.2d**

**Linear mixed-effects model fit by ML**

| **Model information:** | | | **Model fit statistics:** | | | |
|---|---|---|---|---|---|---|
| Number of observations | 60 | | AIC | BIC | LogLikelihood | Deviance |
| Fixed effects coefficients | 15 | | 493.4 | 529.01 | -229.7 | 459.4 |
| Random effects coefficients | 4 | | | | | |
| Covariance parameters | 2 | | | | | |

Formula:

**fEPSPslope~ 1 + timeCond + (1 | ratNum)**

**Fixed effects coefficients (95% CIs):**

| Name | Estimate | SE | tStat | DF | pValue | Lower | Upper |
|---|---|---|---|---|---|---|---|
| (Intercept) | 100 | 5.5637473 | 17.973498 | 45 | 3.67E-22 | 88.794038 | 111.2059623 |
| timeCond_Seg2 | 2.205367204 | 7.8683269 | 0.2802841 | 45 | 7.81E-01 | -13.64226 | 18.05299103 |
| timeCond_Seg3 | -0.23437415 | 7.8683269 | -0.029787 | 45 | 9.76E-01 | -16.082 | 15.61324967 |
| timeCond_Seg4 | -46.81028772 | 7.8683269 | -5.949205 | 45 | 3.71421E-07 | -62.65791 | -30.9626639 |
| timeCond_Seg5 | -39.59281332 | 7.8683269 | -5.031923 | 45 | 8.27084E-06 | -55.44044 | -23.74518949 |
| timeCond_Seg6 | -23.02228677 | 7.8683269 | -2.925944 | 45 | 0.005365489 | -38.86991 | -7.174662941 |
| timeCond_Seg7 | -29.72609985 | 7.8683269 | -3.777944 | 45 | 0.00046198 | -45.57372 | -13.87847602 |
| timeCond_Seg8 | -28.33428967 | 7.8683269 | -3.601057 | 45 | 0.000786954 | -44.18191 | -12.48666585 |
| timeCond_Seg9 | -28.8843559 | 7.8683269 | -3.670965 | 45 | 0.000638401 | -44.73198 | -13.03673208 |
| timeCond_Seg10 | -31.38381479 | 7.8683269 | -3.988626 | 45 | 0.000241668 | -47.23144 | -15.53619097 |
| timeCond_Seg11 | -21.31769632 | 7.8683269 | -2.709305 | 45 | 0.00950421 | -37.16532 | -5.470072494 |
| timeCond_Seg12 | -18.447195 | 7.8683269 | -2.344488 | 45 | 0.023526123 | -34.29482 | -2.599571178 |
| timeCond_Seg13 | 1.532662701 | 7.8683269 | 0.1947889 | 45 | 0.846434999 | -14.31496 | 17.38028653 |
| timeCond_Seg14 | -24.3332408 | 7.8683269 | -3.092556 | 45 | 0.003403079 | -40.18086 | -8.485616976 |
| timeCond_Seg15 | -13.04119071 | 7.8683269 | -1.657429 | 45 | 0.104389375 | -28.88881 | 2.806433114 |

**Random effects covariance parameters (95% CIs):**

Group: ratNum (4 Levels)

| Name1 | Name2 | Type | Estimate | Lower | Upper |
|---|---|---|---|---|---|
| (Intercept) | (Intercept) | std' | 1.24E-15 | NaN | NaN |

Group: Error

| Name | Estimate | Lower | Upper |
|---|---|---|---|
| 'Res Std' | 11.127 | 9.3045 | 13.308 |

**ANOVA marginal tests: DFMethod = 'Residual'**

| Term | FStat | DF1 | DF2 | pValue |
|---|---|---|---|---|
| (Intercept) | 323.0466252 | 1 | 45 | 3.67E-22 |
| timeCond | 7.62163708 | 14 | 45 | 7.64E-08 |

Linear contrasts (Segment 1 vs Seg2–12, Bonferroni corrected):

| Segment | Time period(minute) | Estimate | SE | t_value | df | p_value | p_Bonferroni | F_value | df1 | Cohen_d |
|---|---|---|---|---|---|---|---|---|---|---|
| 1 | [-60 -50] | / | / | / | / | / | / | / | / | / |
| 2 | [-50 -40] | 2.2053672 | 7.8683269 | 0.2802841 | 45 | 0.780544 | 1 | 0.0785592 | 1 | 0.0835646 |
| 3 | [-40 -30] | -0.234374 | 7.8683269 | -0.029787 | 45 | 0.9763686 | 1 | 0.0008873 | 1 | -0.008881 |
| 4 | [-30 -20] | -46.81029 | 7.8683269 | -5.949205 | 45 | 3.714E-07 | 5.19989E-06 | 35.393037 | 1 | -1.77371 |
| 5 | [-20 -10] | -39.59281 | 7.8683269 | -5.031923 | 45 | 8.271E-06 | 0.000115792 | 25.320247 | 1 | -1.50023 |
| 6 | [-10 0] | -23.02229 | 7.8683269 | -2.925944 | 45 | 0.0053655 | 0.075116853 | 8.5611505 | 1 | -0.872348 |
| 7 | [ 5 15] | -29.7261 | 7.8683269 | -3.777944 | 45 | 0.000462 | 0.006467714 | 14.272862 | 1 | -1.126365 |
| 8 | [ 15 25] | -28.33429 | 7.8683269 | -3.601057 | 45 | 0.000787 | 0.011017349 | 12.967608 | 1 | -1.073628 |
| 9 | [ 25 35] | -28.88436 | 7.8683269 | -3.670965 | 45 | 0.0006384 | 0.008937615 | 13.475987 | 1 | -1.09447 |
| 10 | [ 35 45] | -31.38381 | 7.8683269 | -3.988626 | 45 | 0.0002417 | 0.003383357 | 15.909139 | 1 | -1.189179 |
| 11 | [ 45 55] | -21.3177 | 7.8683269 | -2.709305 | 45 | 0.0095042 | 0.133058937 | 7.3403329 | 1 | -0.807759 |
| 12 | [ 55 65] | -18.4472 | 7.8683269 | -2.344488 | 45 | 0.0235261 | 0.329365729 | 5.4966222 | 1 | -0.698991 |
| 13 | [ 65 75] | 1.5326627 | 7.8683269 | 0.1947889 | 45 | 0.846435 | 1 | 0.0379427 | 1 | 0.0580748 |
| 14 | [ 75 85] | -24.33324 | 7.8683269 | -3.092556 | 45 | 0.0034031 | 0.047643109 | 9.5639021 | 1 | -0.922022 |
| 15 | [ 85 95] | -13.04119 | 7.8683269 | -1.657429 | 45 | 0.1043894 | 1 | 2.7470699 | 1 | -0.49415 |

#### Table9.LME model and statistical details of i200-TNS on fEPSP slope with clonidine blockade (Fig.2e)

| | | | | | | |
|---|---|---|---|---|---|---|
| **Reference: Fig.2e** | | | | | | |
| **Linear mixed-effects model fit by ML** | | | | | | |
| **Model information:** | | | **Model fit statistics:** | | | |
| Number of observations | 718 | | AIC | BIC | ogLikelihoo | Deviance |
| Fixed effects coefficients | 3 | | 6454.1 | 6477 | -3222 | 6444.1 |
| Random effects coefficients | 4 | | | | | |
| Covariance parameters | 2 | | | | | |
| Formula: | | | | | | |
| **fEPSPslope~ 1 + timeCond + (1 \| ratNum)** | | | | | | |

**Fixed effects coefficients (95% CIs):**

| Name | Estimate | SE | tStat | DF | pValue | Lower | Upper |
|---|---|---|---|---|---|---|---|
| (Intercept) | 98.98401 | 5.4979882 | 18.003678 | 715 | 4.80E-60 | 88.18987939 | 109.77814 |
| timeCond_Blockade | -26.03111 | 1.9449492 | -13.38396 | 715 | 1.29E-36 | -29.84960867 | -22.21262 |
| timeCond_Post30 | -33.63894 | 1.9469922 | -17.27739 | 715 | 3.71E-56 | -37.4614465 | -29.81644 |

**Random effects covariance parameters (95% CIs):**

Group: ratNum (4 Levels)

| Name1 | Name2 | Type | Estimate | Lower | Upper |
|---|---|---|---|---|---|
| (Intercept) | (Intercept) | std' | 10.646 | 5.2424 | 21.618 |

Group: Error

| Name | Estimate | Lower | Upper |
|---|---|---|---|
| 'Res Std' | 21.284 | 20.208 | 22.417 |

**ANOVA marginal tests: DFMethod = 'Residual'**

| Term | FStat | DF1 | DF2 | pValue |
|---|---|---|---|---|
| (Intercept) | 324.13 | 1 | 715 | 4.80E-60 |
| timeCond | 164.22 | 2 | 715 | 2.05E-59 |

| Comparison | Estimate | SE | t_value | df | p_value | p_Bonferroni | F_value | df1 | Cohen_d |
|---|---|---|---|---|---|---|---|---|---|
| Blockade_vs_Baseline | -26.03111 | 1.9449492 | -13.38396 | 715 | 1.288E-36 | 3.86465E-36 | 179.13028 | 1 | -1.001063 |
| Post30_vs_Baseline | -33.63894 | 1.9469922 | -17.27739 | 715 | 3.714E-56 | 1.1141E-55 | 298.50815 | 1 | -1.292275 |
| Post30_vs_Blockade | -7.607827 | 1.9449492 | -3.911581 | 715 | 0.0001004 | 0.000301308 | 15.300468 | 1 | -0.29257 |

#### Table10.LME model and statistical details of i200-TNS on fEPSP slope with SCH-23390 blockade (Fig.2f)

| | | | | | | | | | |
|---|---|---|---|---|---|---|---|---|---|
| **Reference: Fig.2f** | | | | | | | | | |
| **Linear mixed-effects model fit by ML** | | | | | | | | | |
| **Model information:** | | | **Model fit statistics:** | | | | | | |
| Number of observations | 679 | | AIC | BIC | ogLikelihoo | Deviance | | | |
| Fixed effects coefficients | 3 | | 6557.4 | 6580 | -3273.7 | 6547.4 | | | |
| Random effects coefficients | 4 | | | | | | | | |
| Covariance parameters | 2 | | | | | | | | |
| Formula: | | | | | | | | | |
| **fEPSPslope~ 1 + timeCond + (1 \| ratNum)** | | | | | | | | | |
| **Fixed effects coefficients (95% CIs):** | | | | | | | | | |
| **Name** | **Estimate** | **SE** | **tStat** | **DF** | **pValue** | **Lower** | **Upper** | | |
| (Intercept) | 100.59901 | 1.9470348 | 51.667805 | 676 | 6.12E-237 | 96.77605193 | 104.42198 | | |
| timeCond_Blockade | -35.21244 | 2.8227653 | -12.47445 | 676 | 2.73E-32 | -40.75487732 | -29.66999 | | |
| timeCond_Post30 | -28.89279 | 2.7930129 | -10.34467 | 676 | 2.20E-23 | -34.37680963 | -23.40876 | | |
| **Random effects covariance parameters (95% CIs):** | | | | | | | | | |
| Group: ratNum (4 Levels) | | | | | | | | | |
| **Name1** | **Name2** | **Type** | **Estimate** | **Lower** | **Upper** | | | | |
| (Intercept) | (Intercept) | std' | 6.67E-15 | NaN | NaN | | | | |
| Group: Error | | | | | | | | | |
| **Name** | **Estimate** | **Lower** | **Upper** | | | | | | |
| 'Res Std' | 30.037 | 28.482 | 31.678 | | | | | | |
| **ANOVA marginal tests: DFMethod = 'Residual'** | | | | | | | | | |
| **Term** | **FStat** | **DF1** | **DF2** | **pValue** | | | | | |
| (Intercept) | 2669.5621 | 1 | 676 | 6.12E-237 | | | | | |
| timeCond | 90.092536 | 2 | 676 | 2.06E-35 | | | | | |
| **Comparison** | **Estimate** | **SE** | **t_value** | **df** | **p_value** | **p_Bonferroni** | **F_value** | **df1** | **Cohen_d** |
| Blockade_vs_Baseline | -35.21244 | 2.8227653 | -12.47445 | 676 | 2.734E-32 | 8.20215E-32 | 155.61183 | 1 | -0.959573 |
| Post30_vs_Baseline | -28.89279 | 2.7930129 | -10.34467 | 676 | 2.204E-23 | 6.61264E-23 | 107.01211 | 1 | -0.795744 |
| Post30_vs_Blockade | 6.3196496 | 2.8612997 | 2.208664 | 676 | 0.0275328 | 0.082598309 | 4.8781966 | 1 | 0.1698972 |

### Table11.LME model and statistical details of i200-TNS on spike rate with clonidine blockade (Fig.2g)

**Reference: Fig.2g**

**Linear mixed-effects model fit by ML**

| **Model information:** | | | **Model fit statistics:** | | | |
|---|---|---|---|---|---|---|
| Number of observations | 1500 | | AIC | BIC | LogLikelihood | Deviance |
| Fixed effects coefficients | 15 | | 8394.8 | 8490.5 | -4179.4 | 8358.8 |
| Random effects coefficients | 104 | | | | | |
| Covariance parameters | 3 | | | | | |

Formula:

**SpikeRateMean ~ 1 + timeCond + (1 | ratNum) + (1 | unitID)**

**Fixed effects coefficients (95% CIs):**

| Name | Estimate | SE | tStat | DF | pValue | Lower | Upper |
|---|---|---|---|---|---|---|---|
| (Intercept) | 9.57100257 | 1.1300503 | 8.4695368 | 1485 | 5.84E-17 | 7.354338 | 11.78766714 |
| timeCond_Seg2 | -0.8897 | 0.5083328 | -1.750231 | 1485 | 8.03E-02 | -1.886827 | 0.10742663 |
| timeCond_Seg3 | -1.253366667 | 0.5083328 | -2.465642 | 1485 | 1.38E-02 | -2.250493 | -0.256240036 |
| timeCond_Seg4 | -5.0624 | 0.5083328 | -9.958831 | 1485 | 1.1636E-22 | -6.059527 | -4.06527337 |
| timeCond_Seg5 | -6.5729 | 0.5083328 | -12.93031 | 1485 | 2.55654E-36 | -7.570027 | -5.57577337 |
| timeCond_Seg6 | -7.210166667 | 0.5083328 | -14.18395 | 1485 | 6.38853E-43 | -8.207293 | -6.213040036 |
| timeCond_Seg7 | -6.7703 | 0.5083328 | -13.31864 | 1485 | 2.58793E-38 | -7.767427 | -5.77317337 |
| timeCond_Seg8 | -6.601616667 | 0.5083328 | -12.9868 | 1485 | 1.31931E-36 | -7.598743 | -5.604490036 |
| timeCond_Seg9 | -6.504016667 | 0.5083328 | -12.7948 | 1485 | 1.23823E-35 | -7.501143 | -5.506890036 |
| timeCond_Seg10 | -6.291883333 | 0.5083328 | -12.37749 | 1485 | 1.46819E-33 | -7.28901 | -5.294756703 |
| timeCond_Seg11 | -4.857966667 | 0.5083328 | -9.556666 | 1485 | 4.82849E-21 | -5.855093 | -3.860840036 |
| timeCond_Seg12 | -4.07745 | 0.5083328 | -8.021222 | 1485 | 2.10514E-15 | -5.074577 | -3.08032337 |
| timeCond_Seg13 | -3.983133333 | 0.5083328 | -7.835681 | 1485 | 8.82377E-15 | -4.98026 | -2.986006703 |
| timeCond_Seg14 | -3.7702 | 0.5083328 | -7.416795 | 1485 | 2.00862E-13 | -4.767327 | -2.77307337 |
| timeCond_Seg15 | -3.585816667 | 0.5083328 | -7.054073 | 1485 | 2.65533E-12 | -4.582943 | -2.588690036 |

**Random effects covariance parameters (95% CIs):**

Group: ratNum (4 Levels)

| Name1 | Name2 | Type | Estimate | Lower | Upper |
|---|---|---|---|---|---|
| (Intercept) | (Intercept) | std' | 2.0411 | 0.95007 | 4.3849 |

Group: unitID (100 Levels)

| Name1 | Name2 | Type | Estimate | Lower | Upper |
|---|---|---|---|---|---|
| (Intercept) | (Intercept) | std' | 3.1809 | 2.7283 | 3.7086 |

Group: Error

| Name | Estimate | Lower | Upper |
|---|---|---|---|
| 'Res Std' | 3.5945 | 3.4638 | 3.7301 |

**ANOVA marginal tests: DFMethod = 'Residual'**

| Term | FStat | DF1 | DF2 | pValue |
|---|---|---|---|---|
| (Intercept) | 71.73305346 | 1 | 1485 | 5.84E-17 |
| timeCond | 41.19986379 | 14 | 1485 | 1.77E-95 |

Linear contrasts (Segment 1 vs Seg2–12, Bonferroni corrected):

| Segment | Time period(minute) | Estimate | SE | t_value | df | p_value | p_Bonferroni | F_value | df1 | Cohen_d |
|---|---|---|---|---|---|---|---|---|---|---|
| 1 | [-60 -50] | / | / | / | / | / | / | / | / | / |
| 2 | [-50 -40] | -0.8897 | 0.5083328 | -1.750231445 | 1485 | 0.0802849 | 1 | 3.0633101 | 1 | -0.090837 |
| 3 | [-40 -30] | -1.253367 | 0.5083328 | -2.465642072 | 1485 | 0.0137892 | 0.193048138 | 6.0793908 | 1 | -0.127967 |
| 4 | [-30 -20] | -5.0624 | 0.5083328 | -9.95883069 | 1485 | 1.164E-22 | 1.62904E-21 | 99.178309 | 1 | -0.516863 |
| 5 | [-20 -10] | -6.5729 | 0.5083328 | -12.93030939 | 1485 | 2.557E-36 | 3.57915E-35 | 167.1929 | 1 | -0.671082 |
| 6 | [-10 0] | -7.210167 | 0.5083328 | -14.18395012 | 1485 | 6.389E-43 | 8.94394E-42 | 201.18444 | 1 | -0.736146 |
| 7 | [ 5 15] | -6.7703 | 0.5083328 | -13.31863769 | 1485 | 2.588E-38 | 3.62311E-37 | 177.38611 | 1 | -0.691236 |
| 8 | [ 15 25] | -6.601617 | 0.5083328 | -12.98680125 | 1485 | 1.319E-36 | 1.84703E-35 | 168.65701 | 1 | -0.674014 |
| 9 | [ 25 35] | -6.504017 | 0.5083328 | -12.79480104 | 1485 | 1.238E-35 | 1.73352E-34 | 163.70693 | 1 | -0.664049 |
| 10 | [ 35 45] | -6.291883 | 0.5083328 | -12.3774891 | 1485 | 1.468E-33 | 2.05546E-32 | 153.20224 | 1 | -0.642391 |
| 11 | [ 45 55] | -4.857967 | 0.5083328 | -9.556666311 | 1485 | 4.828E-21 | 6.75988E-20 | 91.329871 | 1 | -0.49599 |
| 12 | [ 55 65] | -4.07745 | 0.5083328 | -8.021221989 | 1485 | 2.105E-15 | 2.94719E-14 | 64.340002 | 1 | -0.416301 |
| 13 | [ 65 75] | -3.983133 | 0.5083328 | -7.8356808 | 1485 | 8.824E-15 | 1.23533E-13 | 61.397894 | 1 | -0.406671 |
| 14 | [ 75 85] | -3.7702 | 0.5083328 | -7.416795091 | 1485 | 2.009E-13 | 2.81206E-12 | 55.008849 | 1 | -0.384931 |
| 15 | [ 85 95] | -3.585817 | 0.5083328 | -7.054073378 | 1485 | 2.655E-12 | 3.71746E-11 | 49.759951 | 1 | -0.366106 |

#### Table12.LME model and statistical details of i200-TNS on spike rate with SCH-23390 blockade (Fig.2h)

**Reference: Fig.2h**

**Linear mixed-effects model fit by ML**

| **Model information:** | | | **Model fit statistics:** | | | |
|---|---|---|---|---|---|---|
| Number of observations | 1230 | | AIC | BIC | LogLikelihood | Deviance |
| Fixed effects coefficients | 15 | | 6551.8 | 6643.9 | -3257.9 | 6515.8 |
| Random effects coefficients | 86 | | | | | |
| Covariance parameters | 3 | | | | | |

Formula:

**SpikeRateMean ~ 1 + timeCond + (1 | ratNum) + (1 | unitID)**

**Fixed effects coefficients (95% CIs):**

| Name | Estimate | SE | tStat | DF | pValue | Lower | Upper |
|---|---|---|---|---|---|---|---|
| (Intercept) | 3.608211382 | 0.4015441 | 8.9858401 | 1215 | 9.58E-19 | 2.8204146 | 4.396008202 |
| timeCond_Seg2 | 0.57449187 | 0.5068763 | 1.1333966 | 1215 | 2.57E-01 | -0.419958 | 1.56894177 |
| timeCond_Seg3 | 0.889776423 | 0.5068763 | 1.7554114 | 1215 | 7.94E-02 | -0.104673 | 1.884226323 |
| timeCond_Seg4 | -3.090914634 | 0.5068763 | -6.097967 | 1215 | 1.44106E-09 | -4.085365 | -2.096464734 |
| timeCond_Seg5 | -3.019756098 | 0.5068763 | -5.95758 | 1215 | 3.34846E-09 | -4.014206 | -2.025306198 |
| timeCond_Seg6 | -2.801117886 | 0.5068763 | -5.526236 | 1215 | 4.0001E-08 | -3.795568 | -1.806667986 |
| timeCond_Seg7 | -2.53300813 | 0.5068763 | -4.997291 | 1215 | 6.66521E-07 | -3.527458 | -1.53855823 |
| timeCond_Seg8 | -2.588760163 | 0.5068763 | -5.107282 | 1215 | 3.79144E-07 | -3.58321 | -1.594310263 |
| timeCond_Seg9 | -2.335 | 0.5068763 | -4.606647 | 1215 | 4.523E-06 | -3.32945 | -1.3405501 |
| timeCond_Seg10 | -1.889756098 | 0.5068763 | -3.728239 | 1215 | 0.0002017 | -2.884206 | -0.895306198 |
| timeCond_Seg11 | -1.868638211 | 0.5068763 | -3.686577 | 1215 | 0.000237316 | -2.863088 | -0.874188311 |
| timeCond_Seg12 | -0.712052846 | 0.5068763 | -1.404786 | 1215 | 0.160340443 | -1.706503 | 0.282397054 |
| timeCond_Seg13 | 0.219308943 | 0.5068763 | 0.4326676 | 1215 | 0.66533306 | -0.775141 | 1.213758843 |
| timeCond_Seg14 | 0.509634146 | 0.5068763 | 1.0054409 | 1215 | 0.314884787 | -0.484816 | 1.504084046 |
| timeCond_Seg15 | -0.048455285 | 0.5068763 | -0.095596 | 1215 | 0.923857301 | -1.042905 | 0.945994615 |

**Random effects covariance parameters (95% CIs):**

Group: ratNum (4 Levels)

| Name1 | Name2 | Type | Estimate | Lower | Upper |
|---|---|---|---|---|---|
| (Intercept) | (Intercept) | std' | 1.41E-09 | NaN | NaN |

Group: unitID (82 Levels)

| Name1 | Name2 | Type | Estimate | Lower | Upper |
|---|---|---|---|---|---|
| (Intercept) | (Intercept) | std' | 1.6394 | 1.3512 | 1.9891 |

Group: Error

| Name | Estimate | Lower | Upper |
|---|---|---|---|
| 'Res Std' | 3.2456 | 3.1155 | 3.3811 |

**ANOVA marginal tests: DFMethod = 'Residual'**

| Term | FStat | DF1 | DF2 | pValue |
|---|---|---|---|---|
| (Intercept) | 80.74532216 | 1 | 1215 | 9.58E-19 |
| timeCond | 17.12158409 | 14 | 1215 | 4.76E-39 |

Linear contrasts (Segment 1 vs Seg2–12, Bonferroni corrected):

| Segment | Time period(minute) | Estimate | SE | t_value | df | p_value | p_Bonferroni | F_value | df1 | Cohen_d |
|---|---|---|---|---|---|---|---|---|---|---|
| 1 | [-60 -50] | / | / | / | / | / | / | / | / | / |
| 2 | [-50 -40] | 0.5744919 | 0.5068763 | 1.1333966 | 1215 | 0.2572713 | 1 | 1.2845879 | 1 | 0.0650315 |
| 3 | [-40 -30] | 0.8897764 | 0.5068763 | 1.7554114 | 1215 | 0.0794409 | 1 | 3.0814693 | 1 | 0.1007212 |
| 4 | [-30 -20] | -3.090915 | 0.5068763 | -6.097967 | 1215 | 1.441E-09 | 2.01748E-08 | 37.185196 | 1 | -0.349886 |
| 5 | [-20 -10] | -3.019756 | 0.5068763 | -5.95758 | 1215 | 3.348E-09 | 4.68785E-08 | 35.492761 | 1 | -0.341831 |
| 6 | [-10 0] | -2.801118 | 0.5068763 | -5.526236 | 1215 | 4E-08 | 5.60014E-07 | 30.539283 | 1 | -0.317082 |
| 7 | [ 5 15] | -2.533008 | 0.5068763 | -4.997291 | 1215 | 6.665E-07 | 9.33129E-06 | 24.972914 | 1 | -0.286732 |
| 8 | [ 15 25] | -2.58876 | 0.5068763 | -5.107282 | 1215 | 3.791E-07 | 5.30802E-06 | 26.08433 | 1 | -0.293043 |
| 9 | [ 25 35] | -2.335 | 0.5068763 | -4.606647 | 1215 | 4.523E-06 | 6.3322E-05 | 21.221195 | 1 | -0.264318 |
| 10 | [ 35 45] | -1.889756 | 0.5068763 | -3.728239 | 1215 | 0.0002017 | 0.002823798 | 13.899769 | 1 | -0.213917 |
| 11 | [ 45 55] | -1.868638 | 0.5068763 | -3.686577 | 1215 | 0.0002373 | 0.00332242 | 13.590847 | 1 | -0.211527 |
| 12 | [ 55 65] | -0.712053 | 0.5068763 | -1.404786 | 1215 | 0.1603404 | 1 | 1.9734245 | 1 | -0.080603 |
| 13 | [ 65 75] | 0.2193089 | 0.5068763 | 0.4326676 | 1215 | 0.6653331 | 1 | 0.1872012 | 1 | 0.0248254 |
| 14 | [ 75 85] | 0.5096341 | 0.5068763 | 1.0054409 | 1215 | 0.3148848 | 1 | 1.0109114 | 1 | 0.0576897 |
| 15 | [ 85 95] | -0.048455 | 0.5068763 | -0.095596 | 1215 | 0.9238573 | 1 | 0.0091386 | 1 | -0.005485 |

#### Table13.LME model and statistical details of PACz after-effect in human(Fig.3j)

**Reference: Fig.3j**

**Linear mixed-effects model fit by REML**

| **Model information:** | | | **Model fit statistics:** | | | |
|---|---|---|---|---|---|---|
| Number of observations | 44 | | AIC | BIC | LogLikelihood | Deviance |
| Fixed effects coefficients | 2 | | -138.039 | -129.351 | 74.01945553 | -148.039 |
| Random effects coefficients | 31 | | | | | |
| Covariance parameters | 3 | | | | | |
| Number of subjects | 9 | | | | | |
| Number of subject-channels | 22 | | | | | |

Formula:

**pac_z ~ period + (1|subject) + (1|subject_channel)**

**Fixed effects coefficients (95% CIs):**

| Name | Estimate | SE | tStat | DF | pValue | Lower | Upper |
|---|---|---|---|---|---|---|---|
| (Intercept) | -0.001942832 | 0.0107629 | -0.180511 | 42 | 0.857619373 | -0.023663 | 0.0197777 |
| period_post_burst | 0.067152663 | 0.010487 | 6.4033912 | 42 | 1.04348E-07 | 0.0459889 | 0.0883164 |

**Random effects covariance parameters (95% CIs):**

Group: SubjectNum (9 Levels)

| Name1 | Name2 | Type | Estimate | Lower | Upper |
|---|---|---|---|---|---|
| (Intercept) | (Intercept) | std | 0.0227874 | 0.0111421 | 0.046604101 |

Group: ChannelNum (22 Levels)

| Name1 | Name2 | Type | Estimate | Lower | Upper |
|---|---|---|---|---|---|
| (Intercept) | (Intercept) | std | 1.708E-10 | NA | NA |

Group: Error

| Name | Estimate | Lower | Upper |
|---|---|---|---|
| 'Res Std' | 0.034781599 | 0.0275064 | 0.0439811 |

**ANOVA marginal tests: DFMethod = 'Residual'**

| Term | FStat | DF1 | DF2 | pValue |
|---|---|---|---|---|
| (Intercept) | 0.032584337 | 1 | 14.042616 | 8.59E-01 |
| period | 41.00341877 | 1 | 34.877802 | 2.32E-07 |

| Comparison | Estimate_Difference | SE | DF1 | DF2 | F_value | t_value | CI95_Lower | CI95_Upper | Cohen_d | p |
|---|---|---|---|---|---|---|---|---|---|---|
| post_burst - pre_burst | 0.067152663 | 0.010487 | 1 | 34.878 | 41.003 | 6.4033912 | 0.045989 | 0.088316 | 1.12 | 2.32E-07 |

#### Table14.LME model and statistical details of i200-TNS effect on Sharp-wave rate in rat (Fig.4b)

**Reference: Fig.4b**

**Linear mixed-effects model fit by ML**

| **Model information:** | | | **Model fit statistics:** | | | |
|---|---|---|---|---|---|---|
| Number of observations | 114 | | AIC | BIC | LogLikelihood | Deviance |
| Fixed effects coefficients | 3 | | 873.16507 | 889.58226 | -430.5825364 | 861.16507 |
| Random effects coefficients | 46 | | | | | |
| Covariance parameters | 3 | | | | | |
| Number of rats | 8 | | | | | |
| Number of channels | 38 | | | | | |

Formula:

**SharpWaveRate ~ epoch + (1|ratNumber) + (1|ChannelID)**

**Fixed effects coefficients (95% CIs):**

| Name | Estimate | SE | tStat | DF | pValue | Lower | Upper |
|---|---|---|---|---|---|---|---|
| (Intercept) | 52.279629 | 5.7351995 | 9.1155729 | 111 | 3.87546E-15 | 40.914948 | 63.64430942 |
| epoch_During | 7.9657895 | 2.0874862 | 3.8159723 | 111 | 0.000223582 | 3.8292962 | 12.10228275 |
| epoch_Post | 1.7473684 | 2.0874862 | 0.8370682 | 111 | 0.404352799 | -2.389125 | 5.883861697 |

**Random effects covariance parameters (95% CIs):**

Group: ratNum (8 Levels)

| Name1 | Name2 | Type | Estimate | Lower | Upper |
|---|---|---|---|---|---|
| (Intercept) | (Intercept) | std | 15.552885 | 9.3788241 | 25.7913182 |

Group: ChannelNum (38 Levels)

| Name1 | Name2 | Type | Estimate | Lower | Upper |
|---|---|---|---|---|---|
| (Intercept) | (Intercept) | std | 2.2575318 | 0.3604788 | 14.13799928 |

Group: Error

| Name | Estimate | Lower | Upper |
|---|---|---|---|
| 'Res Std' | 9.0991414 | 7.7617361 | 10.666992 |

**ANOVA marginal tests: DFMethod = 'Residual'**

| Term | FStat | DF1 | DF2 | pValue |
|---|---|---|---|---|
| (Intercept) | 83.093669 | 1 | 111 | 3.88E-15 |
| epoch | 8.0453989 | 2 | 111 | 5.46E-04 |

| Comparison | nate_Differ | SE | DF1 | DF2 | F_value | t_value | CI95_Lower | CI95_Upper | Cohen_d | p_uncorrected | p_Bonferroni |
|---|---|---|---|---|---|---|---|---|---|---|---|
| Pre vs During | 7.9657895 | 2.0874862 | 1 | 111 | 14.56164433 | 3.8159723 | 3.829296198 | 12.1022827 | 0.875444078 | 0.000223582 | 0.000670746 |
| Pre vs Post | 1.7473684 | 2.0874862 | 1 | 111 | 0.70068325 | 0.8370682 | -2.389124855 | 5.8838617 | 0.192036626 | 0.404352799 | 1 |
| During vs Post | -6.218421 | 2.0874862 | 1 | 111 | 8.873869148 | -2.978904 | -10.35491433 | -2.0819278 | -0.683407451 | 0.003553657 | 0.010660971 |

### Table15.LME model and statistical details of i200-TNS effect on SWR rate in rat (Fig.4c)

| | | | | | | |
|---|---|---|---|---|---|---|
| **Reference: Fig.4c** | | | | | | |
| **Linear mixed-effects model fit by ML** | | | | | | |
| **Model information:** | | | **Model fit statistics:** | | | |
| Number of observations | 114 | | AIC | BIC | ogLikelihoo | Deviance |
| Fixed effects coefficients | 3 | | 584.23889 | 600.65608 | -286.1194 | 572.23889 |
| Random effects coefficients | 46 | | | | | |
| Covariance parameters | 3 | | | | | |
| Number of rats | 8 | | | | | |
| Number of channels | 38 | | | | | |

Formula:

**SWR_Rate ~ epoch + (1|ratNumber) + (1|ChannelID)**

**Fixed effects coefficients (95% CIs):**

| Name | Estimate | SE | tStat | DF | pValue | Lower | Upper |
|---|---|---|---|---|---|---|---|
| (Intercept) | 7.179571596 | 1.5757462 | 4.5562995 | 111 | 1.344E-05 | 4.0571253 | 10.302018 |
| epoch_During | 2.181578947 | 0.6033679 | 3.6156693 | 111 | 0.000452 | 0.9859651 | 3.3771928 |
| epoch_Post | -0.626315789 | 0.6033679 | -1.038033 | 111 | 0.3015107 | -1.82193 | 0.5692981 |

**Random effects covariance parameters (95% CIs):**

Group: ratNum (8 Levels)

| Name1 | Name2 | Type | Estimate | Lower | Upper |
|---|---|---|---|---|---|
| (Intercept) | (Intercept) | std | 4.2676561 | 2.5630189 | 7.1060296 |

Group: ChannelNum (38 Levels)

| Name1 | Name2 | Type | Estimate | Lower | Upper |
|---|---|---|---|---|---|
| (Intercept) | (Intercept) | std | 1.709E-06 | NaN | NaN |

Group: Error

| Name | Estimate | Lower | Upper |
|---|---|---|---|
| 'Res Std' | 2.630019905 | 2.298713 | 3.0090771 |

**ANOVA marginal tests: DFMethod = 'Residual'**

| Term | FStat | DF1 | DF2 | pValue |
|---|---|---|---|---|
| (Intercept) | 20.75986478 | 1 | 111 | 1.34E-05 |
| epoch | 11.93584026 | 2 | 111 | 2.02E-05 |

| Comparison | Estimate_Difference | SE | DF1 | DF2 | F_value | t_value | CI95_Lower | CI95_Upper | Cohen_d | p_uncorrected | p_Bonferroni |
|---|---|---|---|---|---|---|---|---|---|---|---|
| Pre vs During | 2.181578947 | 0.6033679 | 1 | 111 | 13.073064 | 3.6156693 | 0.9859651 | 3.3771928 | 0.8294914 | 0.000452047 | 0.001356141 |
| Pre vs Post | -0.626315789 | 0.6033679 | 1 | 111 | 1.0775123 | -1.038033 | -1.82193 | 0.5692981 | -0.238141 | 0.301510679 | 0.904532037 |
| During vs Post | -2.807894737 | 0.6033679 | 1 | 111 | 21.656944 | -4.653702 | -4.003509 | -1.612281 | -1.067633 | 9.07614E-06 | 2.72284E-05 |

### Table16.LME model and statistical details of i200-TNS effect on ripple rate (Fig.4d)

| | | | | | | |
|---|---|---|---|---|---|---|
| **Reference: Fig.4d** | | | | | | |
| **Linear mixed-effects model fit by ML** | | | | | | |
| **Model information:** | | | **Model fit statistics:** | | | |
| Number of observations | 114 | | AIC | BIC | LogLikelihood | Deviance |
| Fixed effects coefficients | 3 | | 732.15655 | 748.57374 | -360.0782746 | 720.15655 |
| Random effects coefficients | 46 | | | | | |
| Covariance parameters | 3 | | | | | |
| Number of rats | 8 | | | | | |
| Number of channels | 38 | | | | | |

Formula:

**Ripple_Rate ~ epoch + (1|ratNumber) + (1|ChannelID)**

**Fixed effects coefficients (95% CIs):**

| Name | Estimate | SE | tStat | DF | pValue | Lower | Upper |
|---|---|---|---|---|---|---|---|
| (Intercept) | 20.18131185 | 3.9797876 | 5.0709519 | 111 | 1.59794E-06 | 12.295097 | 28.067527 |
| epoch_During | 5.189473684 | 1.1294394 | 4.5947342 | 111 | 1.15191E-05 | 2.9514142 | 7.4275332 |
| epoch_Post | -2.518421053 | 1.1294394 | -2.229797 | 111 | 0.027773379 | -4.756481 | -0.280362 |

**Random effects covariance parameters (95% CIs):**

Group: ratNum (8 Levels)

| Name1 | Name2 | Type | Estimate | Lower | Upper |
|---|---|---|---|---|---|
| (Intercept) | (Intercept) | std | 10.995224 | 6.6658917 | 18.13635048 |

Group: ChannelNum (38 Levels)

| Name1 | Name2 | Type | Estimate | Lower | Upper |
|---|---|---|---|---|---|
| (Intercept) | (Intercept) | std | 2.379E-08 | NaN | NaN |

Group: Error

| Name | Estimate | Lower | Upper |
|---|---|---|---|
| 'Res Std' | 4.923112021 | 4.3030162 | 5.6325682 |

**ANOVA marginal tests: DFMethod = 'Residual'**

| Term | FStat | DF1 | DF2 | pValue |
|---|---|---|---|---|
| (Intercept) | 25.71455355 | 1 | 111 | 1.60E-06 |
| epoch | 24.219271 | 2 | 111 | 1.87E-09 |

| Comparison | Estimate_Difference | SE | DF1 | DF2 | F_value | t_value | CI95_Lower | CI95_Upper | Cohen_d | p_uncorrected | p_Bonferroni |
|---|---|---|---|---|---|---|---|---|---|---|---|
| Pre vs During | 5.189473684 | 1.1294394 | 1 | 111 | 21.11158272 | 4.5947342 | 2.9514142 | 7.4275332 | 1.0541043 | 1.15191E-05 | 3.45573E-05 |
| Pre vs Post | -2.518421053 | 1.1294394 | 1 | 111 | 4.971996881 | -2.229797 | -4.756481 | -0.2803616 | -0.511551 | 0.027773379 | 0.083320138 |
| During vs Post | -7.707894737 | 1.1294394 | 1 | 111 | 46.57423341 | -6.824532 | -9.945954 | -5.4698353 | -1.565655 | 4.88921E-10 | 1.46676E-09 |

### Table17.LME model and statistical details of i200-TNS effect on ripple duration in rat (Fig.4e)

**Reference: Fig.4e**

**Linear mixed-effects model fit by ML**

| **Model information:** | | **Model fit statistics:** | | | |
|---|---|---|---|---|---|
| Number of observations | 114 | AIC | BIC | LogLikelihood | Deviance |
| Fixed effects coefficients | 3 | 720.84156 | 737.25875 | -354.4207777 | 708.84156 |
| Random effects coefficients | 46 | | | | |
| Covariance parameters | 3 | | | | |
| Number of rats | 8 | | | | |
| Number of channels | 38 | | | | |

Formula:

**Ripple_Duration ~ epoch + (1|ratNumber) + (1|ChannelID)**

**Fixed effects coefficients (95% CIs):**

| Name | Estimate | SE | tStat | DF | pValue | Lower | Upper |
|---|---|---|---|---|---|---|---|
| (Intercept) | 38.77721166 | 1.277594633 | 30.351733 | 111 | 1.42373E-55 | 36.245573 | 41.308851 |
| epoch_During | -0.066898799 | 1.179418742 | -0.056722 | 111 | 0.954868756 | -2.403996 | 2.2701982 |
| epoch_Post | 2.098932197 | 1.179418742 | 1.7796327 | 111 | 0.077872697 | -0.238165 | 4.4360292 |

**Random effects covariance parameters (95% CIs):**

Group: ratNum (8 Levels)

| Name1 | Name2 | Type | Estimate | Lower | Upper |
|---|---|---|---|---|---|
| (Intercept) | (Intercept) | std | 2.6582654 | 2.3235944 | 3.041139579 |

Group: ChannelNum (38 Levels)

| Name1 | Name2 | Type | Estimate | Lower | Upper |
|---|---|---|---|---|---|
| (Intercept) | (Intercept) | std | 3.116E-06 | NaN | NaN |

Group: Error

| Name | Estimate | Lower | Upper |
|---|---|---|---|
| 'Res Std' | 5.14096711 | 4.493728404 | 5.8814286 |

**ANOVA marginal tests: DFMethod = 'Residual'**

| Term | FStat | DF1 | DF2 | pValue |
|---|---|---|---|---|
| (Intercept) | 921.2277181 | 1 | 111 | 1.42E-55 |
| epoch | 2.180836042 | 2 | 111 | 1.18E-01 |

| Comparison | Estimate_Difference | SE | DF1 | DF2 | F_value | t_value | CI95_Lower | CI95_Upper | Cohen_d | p_uncorrected | p_Bonferroni |
|---|---|---|---|---|---|---|---|---|---|---|---|
| Pre vs During | -0.066898799 | 1.179418742 | 1 | 111 | 0.003217367 | -0.056722 | -2.403996 | 2.2701982 | -0.013013 | 0.954868756 | 1 |
| Pre vs Post | 2.098932197 | 1.179418742 | 1 | 111 | 3.167092656 | 1.7796327 | -0.238165 | 4.4360292 | 0.4082757 | 0.077872697 | 0.233618092 |
| During vs Post | 2.165830997 | 1.179418742 | 1 | 111 | 3.372198104 | 1.8363546 | -0.171266 | 4.502928 | 0.4212886 | 0.068981301 | 0.206943903 |

### Table18.LME model and statistical details of i200-TNS effect on ripple rate in human (Fig.4g)

**Reference: Fig.4g**

**Linear mixed-effects model fit by REML**

| **Model information:** | | **Model fit statistics:** | | | |
|---|---|---|---|---|---|
| Number of observations | 66 | AIC | BIC | LogLikelihood | Deviance |
| Fixed effects coefficients | 3 | 522.99 | 546.56 | -250.49 | 500.99 |
| Random effects coefficients | 49 | | | | |
| Covariance parameters | 3 | | | | |
| Number of subjects | 9 | | | | |
| Number of subject-channels | 22 | | | | |

Formula:

**rippleRate ~ timeCondition + (timeCondition|subject) + (1|subjectChannel)**

**Fixed effects coefficients (95% CIs):**

| Name | Estimate | SE | tStat | DF | pValue | Lower | Upper |
|---|---|---|---|---|---|---|---|
| (Intercept) | 26.235475 | 4.972348 | 5.276274 | 63 | 0.000002 | 16.299029 | 36.17192 |
| epoch_During | -7.213947 | 2.672266 | -2.699562 | 63 | 0.008904 | -12.55404 | -1.87385 |
| epoch_Post | -5.272482 | 4.913969 | -1.072958 | 63 | 0.287383 | -15.09227 | 4.547302 |

**Random effects covariance parameters (95% CIs):**

Group: SubjectNum (9 Levels)

| Name1 | Name2 | Type | Estimate | Lower | Upper |
|---|---|---|---|---|---|
| (Intercept) | (Intercept) | std | 13.227703 | 7.091468 | 24.673611 |

Group: ChannelNum (22 Levels)

| Name1 | Name2 | Type | Estimate | Lower | Upper |
|---|---|---|---|---|---|
| (Intercept) | (Intercept) | std | 6.345765 | 3.473587 | 11.592839 |

Group: Error

| Name | Estimate | Lower | Upper |
|---|---|---|---|
| 'Res Std' | 7.983308 | 6.102021 | 10.444607 |

**ANOVA marginal tests: DFMethod = 'Residual'**

| Term | FStat | DF1 | DF2 | pValue |
|---|---|---|---|---|
| (Intercept) | 27.839072 | 1 | 63 | 2.00E-06 |
| epoch | 3.645763 | 2 | 63 | 3.18E-02 |

| Comparison | nate_Differ | SE | DF1 | DF2 | F_value | t_value | CI95_Lower | CI95_Upper | Cohen_d | p_uncorrected | p_Bonferroni |
|---|---|---|---|---|---|---|---|---|---|---|---|
| Pre vs During | -7.213947 | 2.6722658 | 1 | 63 | 7.28763431 | -2.699562 | -12.5540442 | -1.87384981 | -0.432775 | 0.008903709 | 0.026711126 |
| Pre vs Post | -5.272482 | 4.9139691 | 1 | 63 | 1.151238735 | -1.072958 | -15.0922659 | 4.547301538 | -0.316304 | 0.287382772 | 0.862148316 |
| During vs Post | -1.941465 | 4.6273223 | 1 | 63 | 0.176035215 | -0.419566 | -11.1884305 | 7.305500895 | -0.116471 | 0.676230211 | 1 |

#### Table19.LME model and statistical details of i200-TNS effect on ripple duration in human (Fig.4h)

**Reference: Fig.4h**

**Linear mixed-effects model fit by REML**

| **Model information:** | | | **Model fit statistics:** | | | |
|---|---|---|---|---|---|---|
| Number of observations | 66 | | AIC | BIC | LogLikelihood | Deviance |
| Fixed effects coefficients | 3 | | 480.88867 | 504.46315 | -229.444333 | 458.88867 |
| Random effects coefficients | 49 | | | | | |
| Covariance parameters | 3 | | | | | |
| Number of subjects | 9 | | | | | |
| Number of subject-channels | 22 | | | | | |

Formula:

**rippleDuration ~ timeCondition + (timeCondition|subject) + (1|subjectChannel)**

**Fixed effects coefficients (95% CIs):**

| **Name** | **Estimate** | **SE** | **tStat** | **DF** | **pValue** | **Lower** | **Upper** |
|---|---|---|---|---|---|---|---|
| (Intercept) | 43.958818 | 2.1987947 | 19.992234 | 63 | 5.67399E-29 | 39.564877 | 48.352758 |
| epoch_During | 3.7752507 | 2.8302902 | 1.3338741 | 63 | 0.18704735 | -1.880633 | 9.4311344 |
| epoch_Post | 2.4440498 | 2.7543867 | 0.88733 | 63 | 0.378276306 | -3.060153 | 7.9482524 |

**Random effects covariance parameters (95% CIs):**

Group: SubjectNum (9 Levels)

| **Name1** | **Name2** | **Type** | **Estimate** | **Lower** | **Upper** |
|---|---|---|---|---|---|
| (Intercept) | (Intercept) | std | 1.8901897 | 0.1350255 | 26.46030956 |

Group: ChannelNum (22 Levels)

| **Name1** | **Name2** | **Type** | **Estimate** | **Lower** | **Upper** |
|---|---|---|---|---|---|
| (Intercept) | (Intercept) | std | 7.915011 | 5.4686212 | 11.45579442 |

Group: Error

| **Name** | **Estimate** | **Lower** | **Upper** |
|---|---|---|---|
| 'Res Std' | 5.8736492 | 4.653025 | 7.414478 |

**ANOVA marginal tests: DFMethod = 'Residual'**

| **Term** | **FStat** | **DF1** | **DF2** | **pValue** |
|---|---|---|---|---|
| (Intercept) | 399.68943 | 1 | 63 | 5.67E-29 |
| epoch | 0.9332897 | 2 | 63 | 3.99E-01 |

| **Comparison** | **nate_Differ** | **SE** | **DF1** | **DF2** | **F_value** | **t_value** | **CI95_Lower** | **CI95_Upper** | **Cohen_d** | **p_uncorrected** | **p_Bonferroni** |
|---|---|---|---|---|---|---|---|---|---|---|---|
| Pre vs During | 3.7752507 | 2.8302902 | 1 | 63 | 1.779219996 | 1.3338741 | -1.880633 | 9.4311344 | 0.3589211 | 0.18704735 | 0.561142051 |
| Pre vs Post | 2.4440498 | 2.7543867 | 1 | 63 | 0.787354454 | 0.88733 | -3.060153 | 7.9482524 | 0.232361 | 0.378276306 | 1 |
| During vs Post | 1.3312009 | 1.7736915 | 1 | 63 | 0.563288666 | 0.7505256 | -2.213239 | 4.8756405 | 0.1265601 | 0.455732287 | 1 |

#### Table20.LME model and statistical details of i200-TNS effect on ripple-triggered-mPFC power (theta band) in human (Fig.4l)

**Reference: Fig.4l**

**Linear mixed-effects model fit by ML**

| **Model information:** | | | **Model fit statistics:** | | |
|---|---|---|---|---|---|
| Number of observations | 99 | | AIC | BIC | LogLikelihood |
| Fixed effects coefficients | 3 | | 513.46973 | 534.23068 | -248.7348625 |
| Random effects coefficients | 64 | | | | |
| Covariance parameters | 5 | | | | |
| Number of subjects | 5 | | | | |
| Number of Subject HPC | 14 | | | | |
| Number of Subject Cortical | 12 | | | | |
| Number of pairs | 33 | | | | |

Formula:

**Response0_200ms_dB ~ 1 + epoch + (1 | Subject) + (1 | SubjectHPC) + (1 | SubjectCortical) + (1 | PairID)**

**Fixed effects coefficients (95% CIs):**

| Name | Estimate | SE | tStat | DF | pValue | Lower | Upper |
|---|---|---|---|---|---|---|---|
| (Intercept) | 1.976694171 | 1.4375494 | 1.3750443 | 96 | 0.172318355 | -0.876819 | 4.830207204 |
| epoch_During | 2.31433608 | 0.6496355 | 3.5625147 | 96 | 0.000573954 | 1.0248198 | 3.603852399 |
| epoch_Post | 0.059458809 | 0.6496355 | 0.0915264 | 96 | 0.927265034 | -1.230058 | 1.348975127 |

**Random effects covariance parameters (95% CIs):**

Group: SubjectNum (5 Levels)

| Name1 | Name2 | Type | Estimate | Lower | Upper |
|---|---|---|---|---|---|
| (Intercept) | (Intercept) | std | 2.9355714 | 1.4596215 | 5.903982352 |

Group: SubjectHPC (14 Levels)

| Name1 | Name2 | Type | Estimate | Lower | Upper |
|---|---|---|---|---|---|
| (Intercept) | (Intercept) | std | 1.2692778 | 0.5979727 | 2.69421318 |

Group: SubjectCortical (12 Levels)

| Name1 | Name2 | Type | Estimate | Lower | Upper |
|---|---|---|---|---|---|
| (Intercept) | (Intercept) | std | 1.994E-10 | NA | NA |

Group: PairI (33 Levels)

| Name1 | Name2 | Type | Estimate | Lower | Upper |
|---|---|---|---|---|---|
| (Intercept) | (Intercept) | std | 4.796E-09 | NA | NA |

Group: Error

| Name | Estimate | Lower | Upper |
|---|---|---|---|
| 'Res Std' | 2.638831963 | 2.2721268 | 3.0647207 |

**ANOVA marginal tests: DFMethod = 'Residual'**

| Term | FStat | DF1 | DF2 | pValue |
|---|---|---|---|---|
| (Intercept) | 1.89074691 | 1 | 96 | 1.72E-01 |
| epoch | 8.249215814 | 2 | 96 | 4.94E-04 |

| Comparison | Estimate_Difference | SE | DF1 | DF2 | F_value | t_value | Cohen_d | p_uncorrected | p_Bonferroni |
|---|---|---|---|---|---|---|---|---|---|
| Pre vs During | 2.31433608 | 0.6496355 | 1 | 96 | 12.69151082 | 3.5625147 | 0.727195263 | 0.000573954 | 0.001721861 |
| Pre vs Post | 0.059458809 | 0.6496355 | 1 | 96 | 0.008377084 | 0.0915264 | 0.018682751 | 0.927265034 | 1 |
| During vs Post | 2.254877272 | 0.6496355 | 1 | 96 | 12.04775953 | 3.4709883 | 0.708512513 | 0.000778737 | 0.00233621 |

#### Table21.LME model and statistical details of i200-TNS effect on ripple-triggered-mPFC power (AlphaSigma band) in human (Fig.4m)

**Reference: Fig.4m**

**Linear mixed-effects model fit by ML**

| **Model information:** | | | **Model fit statistics:** | | |
|---|---|---|---|---|---|
| Number of observations | 99 | | AIC | BIC | LogLikelihood |
| Fixed effects coefficients | 3 | | 572.8411 | 593.60206 | -278.4205523 |
| Random effects coefficients | 64 | | | | |
| Covariance parameters | 5 | | | | |
| Number of subjects | 5 | | | | |
| Number of Subject HPC | 14 | | | | |
| Number of Subject Cortical | 12 | | | | |
| Number of pairs | 33 | | | | |

Formula:

**Response0_200ms_dB ~ 1 + epoch + (1 | Subject) + (1 | SubjectHPC) + (1 | SubjectCortical) + (1 | PairID)**

**Fixed effects coefficients (95% CIs):**

| Name | Estimate | SE | tStat | DF | pValue | Lower | Upper |
|---|---|---|---|---|---|---|---|
| (Intercept) | 2.3251122 | 1.7996282 | 1.2919958 | 96 | 0.199460212 | -1.247122 | 5.8973459 |
| epoch_During | 2.8130443 | 0.882284 | 3.188366 | 96 | 0.001932169 | 1.0617244 | 4.5643642 |
| epoch_Post | 0.2339165 | 0.882284 | 0.2651261 | 96 | 0.791480807 | -1.517403 | 1.9852364 |

**Random effects covariance parameters (95% CIs):**

Group: SubjectNum (5 Levels)

| Name1 | Name2 | Type | Estimate | Lower | Upper |
|---|---|---|---|---|---|
| (Intercept) | (Intercept) | std | 3.6198313 | 2.0881386 | 6.275052152 |

Group: SubjectHPC (14 Levels)

| Name1 | Name2 | Type | Estimate | Lower | Upper |
|---|---|---|---|---|---|
| (Intercept) | (Intercept) | std | 1.6657655 | 0.9685385 | 2.864909169 |

Group: SubjectCortical (12 Levels)

| Name1 | Name2 | Type | Estimate | Lower | Upper |
|---|---|---|---|---|---|
| (Intercept) | (Intercept) | std | 1.369E-09 | NA | NA |

Group: PairI (33 Levels)

| Name1 | Name2 | Type | Estimate | Lower | Upper |
|---|---|---|---|---|---|
| (Intercept) | (Intercept) | std | 6.435E-09 | NA | NA |

Group: Error

| Name | Estimate | Lower | Upper |
|---|---|---|---|
| 'Res Std' | 3.5838546 | 3.0986987 | 4.1449701 |

**ANOVA marginal tests: DFMethod = 'Residual'**

| Term | FStat | DF1 | DF2 | pValue |
|---|---|---|---|---|
| (Intercept) | 1.6692532 | 1 | 96 | 1.99E-01 |
| epoch | 6.2604337 | 2 | 96 | 2.78E-03 |

| Comparison | nate_Differe | SE | DF1 | DF2 | F_value | t_value | Cohen_d | p_uncorrected | p_Bonferroni |
|---|---|---|---|---|---|---|---|---|---|
| Pre vs During | 2.8130443 | 0.882284 | 1 | 96 | 10.16567761 | 3.188366 | 0.6508225 | 0.001932169 | 0.005796507 |
| Pre vs Post | 0.2339165 | 0.882284 | 1 | 96 | 0.070291828 | 0.2651261 | 0.0541186 | 0.791480807 | 1 |
| During vs Post | 2.5791278 | 0.882284 | 1 | 96 | 8.54533161 | 2.9232399 | 0.5967038 | 0.004320516 | 0.012961548 |

#### Table22.LME model and statistical details of i200-TNS effect on ripple-triggered-MTC power (theta band) in human (Fig.4p)

| **Reference: Fig.4p** | | | | | |
|---|---|---|---|---|---|
| **Linear mixed-effects model fit by ML** | | | | | |
| **Model information:** | | | **Model fit statistics:** | | |
| Number of observations | 171 | | AIC | BIC | LogLikelihood |
| Fixed effects coefficients | 3 | | 737.92978 | 763.06309 | -360.9648888 |
| Random effects coefficients | 101 | | | | |
| Covariance parameters | 5 | | | | |
| Number of subjects | 7 | | | | |
| Number of Subject HPC | 16 | | | | |
| Number of Subject Cortical | 21 | | | | |
| Number of pairs | 57 | | | | |

Formula:

**Response0_200ms_dB ~ 1 + epoch + (1 | Subject) + (1 | SubjectHPC) + (1 | SubjectCortical) + (1 | PairID)**

**Fixed effects coefficients (95% CIs):**

| Name | Estimate | SE | tStat | DF | pValue | Lower | Upper |
|---|---|---|---|---|---|---|---|
| (Intercept) | 1.9485801 | 0.6452788 | 3.0197493 | 168 | 0.002924895 | 0.6746803 | 3.2224799 |
| epoch_During | 1.5638801 | 0.3255992 | 4.8030829 | 168 | 3.44067E-06 | 0.9210869 | 2.2066733 |
| epoch_Post | -0.781571 | 0.3255992 | -2.400407 | 168 | 0.017469719 | -1.424364 | -0.138778 |

**Random effects covariance parameters (95% CIs):**

Group: SubjectNum (7 Levels)

| Name1 | Name2 | Type | Estimate | Lower | Upper |
|---|---|---|---|---|---|
| (Intercept) | (Intercept) | std | 2.871E-07 | NA | NA |

Group: SubjectHPC (16 Levels)

| Name1 | Name2 | Type | Estimate | Lower | Upper |
|---|---|---|---|---|---|
| (Intercept) | (Intercept) | std | 2.3848019 | 1.6546519 | 3.437146057 |

Group: SubjectCortical (21 Levels)

| Name1 | Name2 | Type | Estimate | Lower | Upper |
|---|---|---|---|---|---|
| (Intercept) | (Intercept) | std | 0.1554768 | 0.1055214 | 0.229081812 |

Group: PairI (57 Levels)

| Name1 | Name2 | Type | Estimate | Lower | Upper |
|---|---|---|---|---|---|
| (Intercept) | (Intercept) | std | 5.413E-09 | NA | NA |

Group: Error

| Name | Estimate | Lower | Upper |
|---|---|---|---|
| 'Res Std' | 1.7382243 | 1.5532917 | 1.9451746 |

**ANOVA marginal tests: DFMethod = 'Residual'**

| Term | FStat | DF1 | DF2 | pValue |
|---|---|---|---|---|
| (Intercept) | 9.1188857 | 1 | 168 | 2.92E-03 |
| epoch | 26.907277 | 2 | 168 | 7.29E-11 |

| Comparison | nate_Differe | SE | DF1 | DF2 | F_value | t_value | Cohen_d | p_uncorrected | p_Bonferroni |
|---|---|---|---|---|---|---|---|---|---|
| Pre vs During | 1.5638801 | 0.3255992 | 1 | 168 | 23.06960543 | 4.8030829 | 0.7411318 | 3.44067E-06 | 1.0322E-05 |
| Pre vs Post | -0.781571 | 0.3255992 | 1 | 168 | 5.761955157 | -2.400407 | -0.370391 | 0.017469719 | 0.052409156 |
| During vs Post | 2.3454509 | 0.3255992 | 1 | 168 | 51.89027104 | 7.2034902 | 1.1115227 | 1.88347E-11 | 5.65042E-11 |

#### Table23.LME model and statistical details of i200-TNS effect on ripple-triggered-MTC power (AlphaSigma band) in human (Fig.4q)

**Reference: Fig.4q**

**Linear mixed-effects model fit by ML**

| **Model information:** | | | **Model fit statistics:** | | |
|---|---|---|---|---|---|
| Number of observations | 171 | | AIC | BIC | LogLikelihood |
| Fixed effects coefficients | 3 | | 790.53362 | 815.66693 | -387.2668119 |
| Random effects coefficients | 101 | | | | |
| Covariance parameters | 5 | | | | |
| Number of subjects | 7 | | | | |
| Number of Subject HPC | 16 | | | | |
| Number of Subject Cortical | 21 | | | | |
| Number of pairs | 57 | | | | |

Formula:

**Response0_200ms_dB ~ 1 + epoch + (1 | Subject) + (1 | SubjectHPC) + (1 | SubjectCortical) + (1 | PairID)**

**Fixed effects coefficients (95% CIs):**

| Name | Estimate | SE | tStat | DF | pValue | Lower | Upper |
|---|---|---|---|---|---|---|---|
| (Intercept) | 1.4888936 | 0.8780724 | 1.6956388 | 168 | 0.091806732 | -0.244584 | 3.2223711 |
| epoch_During | 0.9260093 | 0.3726125 | 2.4851804 | 168 | 0.013927312 | 0.1904032 | 1.6616154 |
| epoch_Post | -0.984434 | 0.3726125 | -2.641977 | 168 | 0.00902091 | -1.72004 | -0.248828 |

**Random effects covariance parameters (95% CIs):**

Group: SubjectNum (7 Levels)

| Name1 | Name2 | Type | Estimate | Lower | Upper |
|---|---|---|---|---|---|
| (Intercept) | (Intercept) | std | 1.4810504 | 0.4675059 | 4.691941056 |

Group: SubjectHPC (16 Levels)

| Name1 | Name2 | Type | Estimate | Lower | Upper |
|---|---|---|---|---|---|
| (Intercept) | (Intercept) | std | 2.2661886 | 1.4271202 | 3.598583251 |

Group: SubjectCortical (21 Levels)

| Name1 | Name2 | Type | Estimate | Lower | Upper |
|---|---|---|---|---|---|
| (Intercept) | (Intercept) | std | 0.6106141 | 0.2708347 | 1.376668337 |

Group: PairI (57 Levels)

| Name1 | Name2 | Type | Estimate | Lower | Upper |
|---|---|---|---|---|---|
| (Intercept) | (Intercept) | std | 6.242E-08 | NA | NA |

Group: Error

| Name | Estimate | Lower | Upper |
|---|---|---|---|
| 'Res Std' | 1.9892065 | 1.7707708 | 2.2345875 |

**ANOVA marginal tests: DFMethod = 'Residual'**

| Term | FStat | DF1 | DF2 | pValue |
|---|---|---|---|---|
| (Intercept) | 2.8751909 | 1 | 168 | 9.18E-02 |
| epoch | 13.147971 | 2 | 168 | 4.95E-06 |

| Comparison | nate_Differ | SE | DF1 | DF2 | F_value | t_value | Cohen_d | p_uncorrected | p_Bonferroni |
|---|---|---|---|---|---|---|---|---|---|
| Pre vs During | 0.9260093 | 0.3726125 | 1 | 168 | 6.17612146 | 2.4851804 | 0.3834717 | 0.013927312 | 0.041781935 |
| Pre vs Post | -0.984434 | 0.3726125 | 1 | 168 | 6.98004472 | -2.641977 | -0.407666 | 0.00902091 | 0.02706273 |
| During vs Post | 1.9104431 | 0.3726125 | 1 | 168 | 26.28774704 | 5.1271578 | 0.7911376 | 8.02633E-07 | 2.4079E-06 |

#### Table24.Paired t-tests statistical details of i200-TNS effect on face-name-occupation memory task in human (Fig.5c,5d)

| Reference: Fig.5c,5d | | | | | | | | |
|---|---|---|---|---|---|---|---|---|
| **Test item** | **shapiro_wilk_p_value** | **test_type** | **test_statistic** | **p_value_raw** | **p_value_corrected** | **confidence_interval** | **Cohens_d** | **degrees_of_freedom** |
| Encoding effect on name recall | 0.617187809 | 'Paired t-test' | -2.265007837 | 0.029808984 | 0.119235935 | [-10.0332898778761,-0.548720688790604] | -0.37750131 | 35 |
| Encoding effect on occupation recall | 0.409563162 | 'Paired t-test' | 2.106092746 | 0.042438759 | 0.169755038 | [0.201629582129099,10.9756191012042] | 0.35101546 | 35 |
| Consolidation effect on name recall | 0.290224054 | 'Paired t-test' | 0.300731239 | 0.765398692 | 1 | [-3.99345291788106,5.38234181788106] | 0.05012187 | 35 |
| Consolidation effect on occupation recall | 0.206989745 | 'Paired t-test' | -3.400875816 | 0.001693779 | **0.006775116** | [-12.9381453838883,-3.26555833277834] | -0.56681264 | 35 |

Note: n=36. A Bonferroni correction was applied across four prespecified comparisons

#### Table25.Cluster-based permutation tests statistical details of i200-TNS effect on visual ERP in human (Fig.6b)

**Reference: Fig.6b**

We relied on the cluster-based permutation tests in FieldTrip to compare ERP changes between LTM and STM. The key settings for the ft_timelockstatistics() function are listed below:

| Parameter | value | note |
|---|---|---|
| latency | [0, 1.5] | |
| method | montecarlo | |
| statistic | depsamplesT | |
| correctm | cluster | |
| clusteralpha | 0.05 | |
| clusterstatistic | maxsum | |
| minnbchan | 2 | |
| tail | 0 | |
| clustertail | 0 | |
| alpha | 0.05 | |
| numrandomization | 1000 | |

#### Table26. LME model and statistical details of i200-TNS effect on EEG PSD power analysis in human (Fig.6d-h)

**Reference: Fig.6d-h**

We relied on the cluster-based permutation tests in FieldTrip to compare resting power changes (R2 vs. R1 and R3 vs. R2) . The key settings for the ft_freqstatistics() function are listed below:

| Parameter | value | note |
|---|---|---|
| method | montecarlo | |
| statistic | depsamplesT | |
| correctm | cluster | |
| clusteralpha | 0.05 | |
| clusterstatistic | maxsum | |
| minnbchan | 2 | |
| tail | 0 | |
| clustertail | 0 | |
| alpha | 0.05 | |
| numrandomization | 1000 | |

Left-frontal channel power between different resting stages was compared using LME models. The statistical tests results are listed below:

**eTNS group, Beta band**

**ANOVA marginal tests: DFMethod: 'Residual'**

| Term | Fstat | DF1 | DF2 | pValue |
|---|---|---|---|---|
| {'(Intercept)'} | 202.76 | 1 | 96 | 2.11E-25 |
| {'Condition' } | 2.6429 | 2 | 96 | 0.076325 |

**eTNS group, Gamma band**

**ANOVA marginal tests: DFMethod: 'Residual'**

| Term | Fstat | DF1 | DF2 | pValue |
|---|---|---|---|---|
| {'(Intercept)'} | 356.19 | 1 | 96 | 4.51E-34 |
| {'Condition' } | 0.89704 | 2 | 96 | 0.41116 |

**Control group, Beta band**

**ANOVA marginal tests: DFMethod: 'Residual'**

| Term | Fstat | DF1 | DF2 | pValue |
|---|---|---|---|---|
| {'(Intercept)'} | 239.86 | 1 | 96 | 7.49E-28 |
| {'Condition' } | 6.5461 | 2 | 96 | 0.0021623 |

**Post-hoc comparisons with Wald F test**

| | R1 vs R2 | R1 vs R3 | R2 vs R3 |
|---|---|---|---|
| Raw pValue | 0.0012 | 0.0051 | 0.6359 |
| Bonferroni pValue | 0.0035 | 0.0152 | 1 |

**Control group, Gamma band**

**ANOVA marginal tests: DFMethod: 'Residual'**

| Term | Fstat | DF1 | DF2 | pValue |
|---|---|---|---|---|
| {'(Intercept)'} | 339.12 | 1 | 96 | 2.87E-33 |
| {'Condition' } | 3.2346 | 2 | 96 | 0.043705 |
